# Experience of the COVID-19 pandemic in rural Odisha, India: knowledge, preventative actions, and impacts on daily life

**DOI:** 10.1101/2020.11.20.20235630

**Authors:** Valerie Bauza, Gloria D. Sclar, Alokananda Bisoyi, Ajilé Owens, Apurva Ghugey, Thomas Clasen

## Abstract

We conducted 131 semi-structured phone interviews with householders in rural Odisha, India to explore participants’ COVID-19 related knowledge, perceptions, and preventative actions, as well as how the pandemic was impacting their daily life, economic and food security, and the village-level response. Interviews were conducted with 73 heads of household, 37 primary caregivers, and 21 members of village water and sanitation committees from 43 rural villages in Ganjam and Gajapati districts in Odisha state. The study took place between May-July 2020 throughout various lockdown restrictions and at a time when many migrant workers were returning to their villages. Most respondents could name at least one correct symptom of COVID-19 (75%), but there was lower knowledge about causes of the disease and high-risk groups, and overall COVID-19 knowledge was lowest among caregivers. Respondents reported high compliance with important preventative measures, including staying home as much as possible (94%), social distancing (91%), washing hands frequently (96%), and wearing a facial mask (95%). Additionally, many respondents reported job loss (31%), financial challenges (93%), challenges related to staying home whether as a preventative measure or due to lockdowns (57%), changes in types and/or amount of food consumed (61%), and adverse emotional effects as a result of the pandemic and lockdown. We also provide detailed summaries of qualitative responses to allow for deeper insights into the lived experience of villagers during this pandemic. Although the research revealed high compliance with preventative measures, the pandemic and associated lockdowns also led to many challenges and hardships faced in daily life particularly around job loss, economic security, food security, and emotional wellbeing. The results underscore the vulnerability of marginalized populations to the pandemic and the need for measures that increase resilience to large-scale shocks.

## Introduction

The World Health Organization (WHO) declared COVID-19, the disease caused by the novel coronavirus virus SARS-CoV-2, as a global pandemic on March 11, 2020 (1). The pandemic quickly resulted in lockdowns around the world. India implemented its own nationwide lockdown on March 24, 2020, limiting the movement of 1.3 billion people (2). The national lockdown in India temporarily shut down portions of the economy and substantially altered daily life, generating fear about economic and food security among the many living in poverty and resulting in a mass exodus of millions of migrants workers from cities to their rural villages. Although initially there were limited services for migrant workers, the government began organizing transportation for returning migrants in May 2020 and quarantine centers were set up in many villages or other centralized locations to temporarily house migrants upon returning from other regions. Additionally, while the strict nationwide lockdown eased on May 1, 2020, it was replaced by a series of lockdowns of varying levels of restriction imposed throughout India by national, state, and district governments over the next several months (3). Government and health authorities in India also undertook massive awareness campaigns to educate the public on COVID-19, emphasizing the need for frequent handwashing, social distancing, staying home as much as possible, avoiding touching one’s face, and good respiratory hygiene, including wearing a face mask (4,5).

The objectives of this research were to understand the COVID-19 related knowledge, perceptions, and preventative actions of rural householders and communities, and how the pandemic was impacting their daily life, economic security, and food security. We conducted semi-structured phone interviews to capture participants experiences. The interviews took place throughout various lockdown restrictions in rural Odisha, India at a time when many migrant workers were returning to their villages. Our aim is to contribute to an understanding of participants’ knowledge, actions, and lived experience during this time in order to determine if the government response, in the form of lockdowns, restrictions, and awareness campaigns, were effective at changing behavior and whether they had unintended negative consequences. These findings can be used to help develop and target interventions that minimize adverse effects and improve overall resilience to external shocks, such as a pandemic, increasingly faced by marginalized populations.

## Methods

### Study site and sampling frame

We conducted semi-structured phone interviews with households in Ganjam and Gajapati districts of rural Odisha, India from May-July 2020. In order to capture a range of perspectives and experiences, the phone interviews were conducted with three types of respondents from each district: head of households (HOH), primary caregivers of young children (<5 years old), and members of Village Water and Sanitation Committees (VWSC). We specifically targeted HOH to understand household-level impacts and decision-making, caregivers to understand childcare practices, and VWSC members to understand village-level response and collective action. In some cases the HOH interview was completed by another household member requested by the HOH (such as the HOH’s wife or son), and in a few cases the caregiver interview was completed by her husband when the caregiver was not fluent in Odia. Details of specific village and respondent selection are provided in the Supplemental Information (SI). Additionally, each of the two districts has distinct geographic and demographic characteristics to capture a wider range of experiences. Ganjam is a coastal district with relatively large villages that are typically closer to cities or markets and have predominately Hindu populations. Comparatively, Gajapati is a hilly and mountainous district with more remote, smaller villages that have predominantly Christian populations belonging to Scheduled Tribes.

All included villages had previously participated in a village-level WASH intervention which included the construction of a community piped water system with household connections, as well as village-level mobilization for the construction of latrines with attached bathing rooms at each household (6). The villages are also a subset of villages that are currently enrolled in a randomized-controlled trial evaluating a child feces management (CFM) intervention (ISRCTN15831099), however intervention delivery had not yet started at the time of this study.

Many governmental restrictions and initiatives were instituted at national, state and district level in response to COVID-19. Ganjam was the first district in Odisha to implement a lockdown on March 16, 2020, which shutdown religious institutions, public transportation, and non-essential shops, limited the time of day that shops selling essential commodities could be open, and prohibited public gatherings. Soon after, similar lockdowns were implemented across Odisha and nationwide, including a prohibition on inter-state travel. Although the nationwide lockdown ended on May 1, 2020, it was replaced by zone-specific restrictions and lockdowns at the district or block level, which changed over time to ease or tighten restrictions based on local case counts. A timeline of these activities in relation to the data collection period is provided in Figure 1. During this time, various monetary and public health initiatives were also taken up by the state and central governments. This included subsidizing monthly wages for some employees and providing additional monthly food grain to the marginalized, offering monetary incentives for returned migrants who completed institutional quarantine, setting up dedicated COVID-19 hospitals, and health screening of residents.

**Figure 1.**
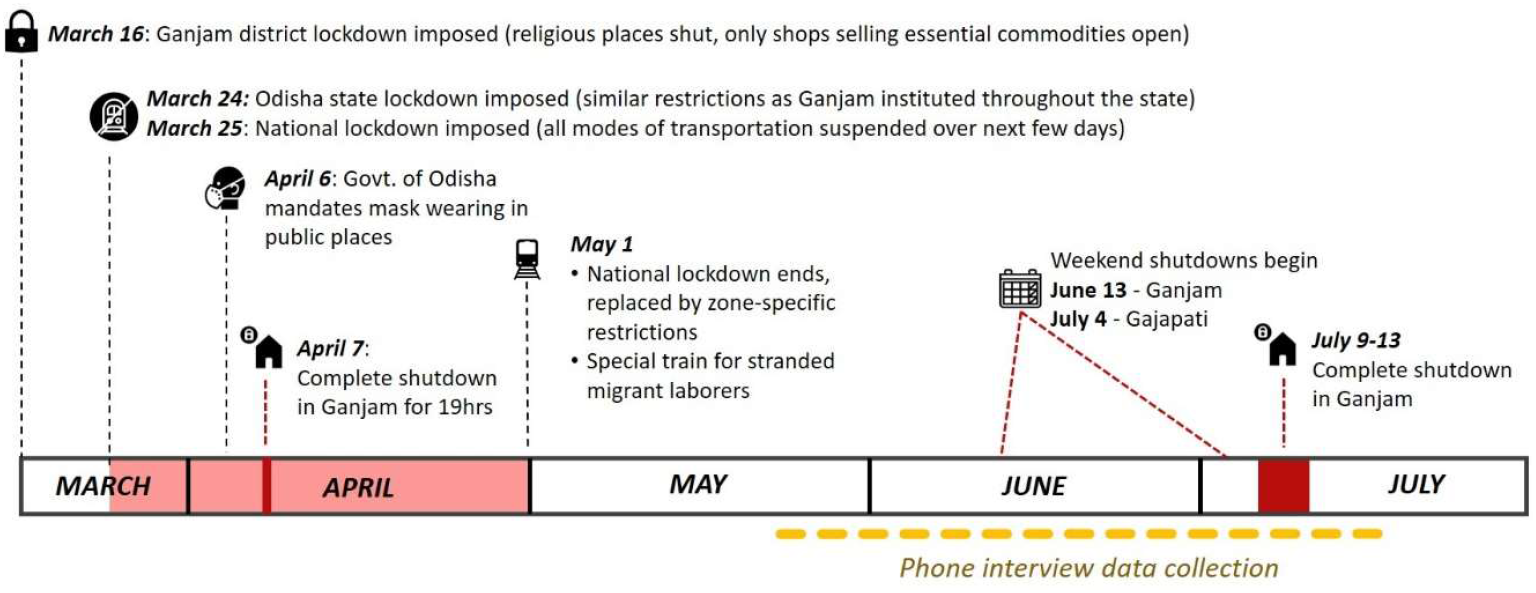
Timeline of local COVID-19 restrictions in relation to the data collection period for the semi-structured phone interviews.

### Interview tool

A semi-structured interview tool was developed for each respondent type with a mix of structured and open-ended questions. All respondents were asked questions about their COVID-19 related knowledge, risk perception, and practices, including any preventative actions taken. All respondents were also asked about how the pandemic or lockdowns had impacted their daily life and social interactions. The HOH and VWSC interview tools also included questions about the impact on finances, food security, and cooking fuel, as well as additional knowledge and actions questions, including challenges related to following recommended preventative actions. The VWSC tool included a specific section on village dynamics and response to the pandemic. Lastly, the caregiver interview tool included a specific section on the impact of the pandemic on childcare practices. The interview tools are provided in the SI. Several of the quantitative questions were modified from the Population Council (7,8) and Y-RISE (9) surveys.

### Data collection and calling procedures

A team of three research assistants, all originally from Ganjam district and fluent in the local language Odia, were trained on how to conduct the phone interviews. The team underwent a multi-day remote training in May 2020, followed by a pilot to revise and finalize the interview tool. Target respondents were randomly ordered using a computer-generated sequence, and research assistants were provided call lists to contact respondents in random order. Details of the specific calling procedure are provided in the SI.

### Data analysis

Quantitative analysis of structured interview questions was done using descriptive statistics and chi-squared tests in Stata 16.1 (StataCorp LLC, College Station, Texas, USA). For qualitative analysis, a modified form of thematic analysis was used to analyze responses to open-ended questions, as well as “please explain” follow-ups to some of the structured questions. For the modified thematic analysis, a research team member read through all responses for a given question and wrote memo notes with reflections, noting common and interesting answers. Then the researcher read through all responses again and categorized responses into initial themes. A third read through was done, as needed, to finalize the themes.

During quality checks after data collection was complete, it was found that one of the research assistants had sometimes skipped select interview questions or fabricated data for some of their interviews. To ensure the accuracy of the data collected, a study supervisor listened to every audio recording of this research assistant’s interviews and corrected the data accordingly prior to analysis. As a result of these skipped questions in some interviews, our sample sizes vary slightly among different questions.

### Ethics

Verbal informed consent was obtained from all participants before starting the interview. At the end of the interview, participants were read a list of helplines they could call if they needed assistance and provided information on preventative measures to protect themselves against COVID-19. This study was approved by the Institutional Review Board (IRB) of Emory University (IRB00115339).

## Results

### Characteristics of study participants

A total of 131 participants were interviewed: 73 HOH, 21 VWSC members, and 37 caregivers. The respondents represented 43 villages (26 in Ganjam and 17 in Gajapati) and two-thirds of all respondents were from Ganjam district. Approximately 8% of respondents (5 HOH, 1 VWSC, 5 caregivers) ended the interview early. Overall, the majority of respondents were male, had at least a primary education, were self-employed, and their household had access to piped water (Table 1).

**Table 1.** Respondent demographics.

|  | **All**<br>**(N=131)** | **HOH**<br>**(N=73)** | **VWSC**<br>**(N=21)** | **Caregiver**<br>**(N=37)** |
| --- | --- | --- | --- | --- |
| Respondent sex: Male [N (%)] | 85 (65%) | 64 (88%) | 18 (86%) | 3 (8%) |
| Respondent age, yr [mean (sd)] | 35.8 (12) | 36.8 (11) | 48.8 (12) | 26.0 (4) |
| Number of household members [mean (sd)] | 5.6 (2) | 5.8 (2) | 4.8 (2) | 5.7 (2) |
| Household has child <5 yr [N (%)] | 69 (53%) | 29 (40%) | 3 (14%) | 37 (100%) |
| Household has functional piped water | 109 (85%) | 60 (82%) | 19 (90%) | 30 (86%) |
| Has Antodaya and/or ration card* [N (%)] | 98 (78%) | 53 (78%) | 14 (70%) | 31 (84%) |
| Completed primary education* [N (%)] | 79 (63%) | 44 (65%) | 17 (85%) | 18 (49%) |
| Occupation*,† [N (%)] |  |  |  |  |
| Employed | 32 (26%) | 23 (34%) | 6 (30%) | 3 (8%) |
| Self-Employed | 71 (57%) | 45 (66%) | 16 (80%) | 10 (27%) |
| Student | 2 (2%) | 2 (3%) | 0 | 0 |
| Unemployed | 31 (25%) | 6 (9%) | 1 (5%) | 24 (65%) |
| Caste/tribe* [N (%)] |  |  |  |  |
| General | 22 (18%) | 13 (19%) | 5 (25%) | 4 (11%) |
| Scheduled caste | 8 (6%) | 4 (6%) | 2 (10%) | 2 (5%) |
| Scheduled tribe | 55 (44%) | 34 (50%) | 8 (40%) | 13 (35%) |
| Other backward caste | 33 (26%) | 16 (24%) | 5 (25%) | 12 (32%) |
| Other | 3 (2%) | 1 (1%) | 0 | 2 (5%) |
| Don't know | 4 (3%) | 0 | 0 | 4 (11%) |
| Religion* [N (%)] |  |  |  |  |
| Hindu | 87 (70%) | 45 (66%) | 14 (70%) | 28 (76%) |
| Christian | 37 (30%) | 22 (32%) | 6 (30%) | 9 (24%) |
| Other | 1 (1%) | 1 (1%) | 0 | 0 |
| District [N (%)] |  |  |  |  |
| Ganjam | 88 (67%) | 47 (64%) | 14 (67%) | 27 (73%) |
| Gajapati | 43 (33%) | 26 (36%) | 7 (33%) | 10 (27%) |

\* Total N=125 (68 HOH, 20 VWSC, 37 Caregiver) due to some HOH/VWSC respondents ending interview early
† Employed was defined as engaged in work outside the home (e.g., labourer, factory, etc.), self-employed as work inside the home (e.g., agriculture, potter, etc.), and unemployed as no work that earns money. Some respondents reported multiple types of occupation.

HOH and VWSC respondents were also asked questions about their household members to assess whether any had characteristics that could put them at higher risk for developing severe illness from COVID-19. Among these respondents, 48% (N=45) had an elderly household member over 60 years old, 17% (N=16) had a household member with a pre-existing condition, with diabetes being the most common (10% of households, N=9), and 4% (N=4) had a household member who was pregnant.

### COVID-19 Knowledge

All respondents confirmed they had “heard of a disease called COVID-19, coronavirus, or corona”.

#### Symptoms

The majority of respondents (75%, N=98) were able to list a correct symptom of the disease (Table S1 in Supplemental Information), although almost half of caregiver respondents (46%, N=17) reported they did not know any symptoms of the disease. Among all symptoms, cough (68%, N=88) and fever (42%, N=55) were most commonly reported. Only one respondent listed ‘loss of taste or smell,’ a unique symptom of COVID-19. Additionally, 23% of respondents (N=30) incorrectly reported sneezing as a symptom.

#### High risk groups

Respondents were asked about the types of people that are at higher risk of becoming very seriously sick if infected with COVID-19. The most common high-risk group mentioned was the elderly (45%, N=58), followed by people with weak immune systems (12%, N=15), children or babies (14%, N=18) and ‘everyone’ (6%, N=8). However, almost a third of respondents (30%, N=39) stated they did not know of any high-risk groups, with caregivers being the most likely to report they did not know any (51%, N=19).

#### Causes

While many respondents said they did not know what causes someone to become sick with COVID-19 (44%, N=58), a similar percentage (44%, N=58) correctly explained that the virus was transmitted from person-to-person contact with a sick person, and/or described behaviors that put a person at risk for infection. Among the few respondents that gave an inaccurate explanation of what causes COVID-19 (12%, N=15), most described perceived causes of illness more broadly, such as eating cold food, not drinking hot water, or coming from/being in a cold area. One respondent mentioned avoiding foods that make the body more susceptible to colds, specifying bananas, pineapple, and lemon – potentially referencing principles from ayurvedic medicine.

#### Sources of information

HOH and VWSC members were first asked what their main source for information about COVID-19 was and then were asked what other sources they received information from, with a list of sources read to them. Most reported that their main source of information related to COVID-19 was the news (66%, N=59) mostly from television (N=55), conversations with a community-level government worker such as an Anganwadi worker (preschool/childcare center worker) or Accredited Social Health Activist (ASHA; community health worker) (31%, N=28), and/or social media/internet (29%, N=26). In general, most respondents reported receiving some information about COVID-19 from a variety of different sources (Table S2). No specific sources were associated with a greater likelihood of a respondent reporting a correct symptom of COVID-19. However, getting their main source of information from social media or the internet was associated with respondents being more likely to know a high risk group (χ^2^=4.47, p=0.04), while getting their main source of information from the news was associated with being more likely to know a correct cause of COVID-19 (χ^2^=9.38, p=0.002). Across the most common main information sources, the majority of respondents reported trusting all of the information received from their main source (73% for news, 70% for Anganwadi/ASHA worker, 77% for social media/internet), with fewer reporting that they trusted some of the information (25% for news, 30% for Anganwadi/ASHA worker, 23% for social media/internet), and only one person who got their main information from the television news and social media/internet reporting that they did not trust any of the information from these sources.

### COVID-19 Perceptions

In regard to self-perceived risk of their chances of contracting COVID-19, the majority of respondents (59%, N=74) felt they had no risk of getting COVID-19, about one-quarter (27%; N=35) said they did not know their risk, and only 13% (N=17) reported feeling either a low, medium, or high level of risk (Table S3). Those who felt no risk explained this was due to them taking precautions, the lack of local cases, or their good health status.

When asked how concerned they would be if they or a family member contracted COVID-19, the majority of respondents (77%, N=68) stated they would be ‘very concerned’, while less answered ‘somewhat concerned’ (13%, N=11), ‘not concerned’ (5%, N=4), or ‘don’t know’ (6%, N=5) (Table S4). For the respondents who said they would be ‘very concerned’, the most common explanations were concern around transmitting the virus to other members of the family, concerns around lethality or severity of the virus, and concern simply because a member of their family contracted the virus. There was no difference in self-perceived risk of their chances of contracting COVID-19 by main source of information, but those who reported getting their main source of information from social media or the internet were less likely to report being very concerned if they or a family member contracted COVID-19 (χ^2^=5.73, p=0.02). Additionally, there was no difference in risk perception or level of concern by district.

### COVID-19 Preventative Actions

#### Preventative measures

Respondents were read a list of specific actions and asked if they or their family members had practiced that action in the past 7 days to protect themselves from COVID-19 (Table 2). Almost all respondents reported practicing key preventative actions: 96% washed hands more often, 95% wore a mask, 94% stayed at home as much as possible, and 91% kept distance from others. There was no difference in proportion of respondents that reported practicing all four of these key preventative measures by district or main source of information. Respondents also commonly reported avoiding public transit and traveling (81%) and hospitals/clinics (74%). Prior to this structured question, HOH and VWSC members were asked the same question in an open-ended format. Responses to the open-ended question were similar to those given in the structured question, and emphasized many of the key preventative measures:

**Table 2.** Preventative measures reported by respondents when asked “In the past 7 days, what actions have you and your family taken to avoid getting coronavirus, if any?” with each action read as a structured yes/no response.

|  | All<br>(N=126)* | HOH<br>(N=69)* | VWSC<br>(N=20) | Caregiver<br>(N=37) |
| --- | --- | --- | --- | --- |
| Stayed at home as much as possible | 118 (94%) | 63 (91%) | 20 (100%) | 35 (95%) |
| Did not attend school or work | 78 (62%) | 40 (58%) | 9 (45%) | 29 (78%) |
| Did not attend social gatherings (e.g. weddings, funerals, church, temple) | 108 (86%) | 58 (84%) | 15 (75%) | 35 (95%) |
| Kept a distance from others | 117 (91%) | 66 (92%) | 18 (90%) | 33 (89%) |
| Washed hands/used hand sanitizer more frequently | 121 (96%) | 65 (94%) | 20 (100%) | 36 (97%) |
| Wore a face mask | 117 (95%) | 63 (95%) | 19 (95%) | 35 (95%) |
| Wore gloves on your hands | 14 (12%) | 6 (9%) | 3 (15%) | 5 (14%) |
| Tried to stop touching face | 59 (49%) | 35 (55%) | 15 (75%) | 9 (24%) |
| Did not shake hands with others | 106 (88%) | 53 (83%) | 18 (90%) | 35 (95%) |
| Covered your mouth with elbow when you sneezed or coughed | 68 (57%) | 34 (54%) | 7 (35%) | 27 (73%) |
| Drank local alcohol | 6 (5%) | 4 (6%) | 0 | 2 (5%) |
| Scrubbed/cleaned surfaces such as door handles, faucets, phone, etc. | 41 (35%) | 27 (44%) | 11 (55%) | 3 (8%) |
| Informed people of illness symptoms | 48 (40%) | 30 (48%) | 15 (75%) | 3 (8%) |
| Contacted a coronavirus helpline | 4 (3%) | 2 (3%) | 2 (10%) | 0 |
| Avoided hospitals/clinics | 88 (74%) | 48 (77%) | 15 (75%) | 25 (68%) |
| Avoided public transit/traveling | 96 (81%) | 48 (79%) | 14 (70%) | 34 (92%) |
| Nothing | 0 | 0 | 0 | 0 |
| Other | 1 (1%) | 1 (1%) | 0 | 0 |

\* These are the total N for respondents that answered about any of the preventative measures. However, the specific N for some actions may be slightly lower as some response options were skipped by a research assistant during a few interviews.

> *“We are washing our hands and feet. We are wearing masks and maintaining distance*.*” Male respondent, 42 years old, Ganjam district (May 2020)*
>
> *“To avoid corona, we are wearing masks while going out and using [waterless hand] sanitizer to clean our hands. When we come home from outside, we must not take the clothes inside. We should keep it outside and change our clothes*.*” Male respondent, 35 years old, Gajapati district (June 2020)*
>
> *“After coming from work, we clean our hands nicely with a good soap. Earlier, we were not doing it because there was no fear for this fever [COVID-19]*.*” Male respondent, 31 years old, Gajapati district (June 2020)*

#### Actions if Exhibiting Symptoms

HOH and VWSC members were asked what actions they would take if they were to begin experiencing symptoms of COVID-19. The majority of respondents (68%, N=60) said they would go to a health clinic or hospital. Some respondents also described calling a coronavirus helpline (22%, N=16), informing a government worker or elected leader, such as an ASHA worker or Village Sarpanch (17%, N=12), or going to get tested for COVID-19 (11%, N=10).

### Challenges related to staying home and social distancing

Many respondents experienced challenges with staying home as much as possible (57%, N=50) and some experienced challenges with social distancing (28%, N=25; Table 3). Many challenges with staying home were related to the lockdown or curfews, including challenges meeting basic needs, problems accessing healthcare, negative impacts on emotional well-being, and difficulty traveling due to police enforcement of lockdown restrictions on travel. Challenges reported related to social distancing included that not everyone was following the guidelines and it was not always possible to maintain distance (see Table 3 for descriptive themes).

**Table 3.** Reported challenges related to following COVID-19 precautions of staying home as much as possible and social distancing.

| Interview Question | Quantitative Results | Qualitative Themes and Descriptions |
| --- | --- | --- |
| What are some of the challenges you and your family face in being able to stay at home at all times except when you are performing essential activities such as working or buying food? | 57% (N=50) described facing challenges, often due to the lockdown or curfews | • <u>Challenge meeting basic needs:</u> The most common challenge respondents faced was being able to meet their basic needs during lockdown or when staying home. Many explained it is a challenge to stay home when one needs to work to have money, repay loans, or feed their family. Others expressed problems getting essential items due to lockdown, curfews, or closed shops, such as food or seeds and fertilizer for their fields. One respondent explained that he needed money but was unable to withdraw it from the bank during the lockdown. Multiple respondents had difficulties selling their fruits/vegetables during the lockdown. Yet another respondent had difficulty getting support for his special needs child because he could not go to the administrative block office or get required paperwork.<br>• <u>Problems accessing healthcare:</u> Multiple respondents explained they would not access the health clinic or hospital if they needed to because they are afraid of catching the virus. One person said that someone had been seriously ill a couple days before our interview, and they had a problem taking the person to the hospital. Another respondent said that he could not get planned treatment for a health condition due to COVID-19.<br>• <u>Negative emotional response:</u> Many respondents expressed emotional challenges with staying home, saying it makes them feel annoyed, irritated, bored, gloomy, or lazy. Others felt it was difficult to stay home because humans are social, and they wanted to meet their friends. A couple of respondents also explained that one needs to move their body to stay healthy and it does not feel good to stay home for a long time.<br>• <u>Difficulty travelling due to police:</u> A few respondents explained that police were not allowing people to travel or go out, even for essential goods. These respondents said a person might get beaten up by the police. One respondent described how the police took pictures of his vehicle when he went to buy medicine and he was worried about getting a fine. |
| What are some of the challenges you and your family face in being able to maintain 2 meters of distance between other non-family members when you are outside of your house? | 28% (N=25) reported challenges with social distancing | • <u>Not everyone is following guidance:</u> Many respondents explained that not all people are listening to this guidance, that some people will come closer to you even if you try to stay away or ask them to maintain distance.<br>• <u>Not always possible to maintain distance:</u> Many respondents explained that it is simply not always possible to maintain distance from others. Some respondents said it was difficult to maintain distance from their friends. Other respondents explained that their job did not allow them to maintain distance (e.g., auto rickshaw driver, farmers who go to the fields in a group, a man who works with another person). Some respondents said that it is not possible to maintain distance in their village at all. |

### Impacts on economic and food security

The pandemic had major impacts on households’ economic and food security (Table 4). Many respondents (31%, N=28) reported they or a family member had lost a job as a result of the pandemic, mainly the loss of work as a daily laborer. Almost all households (93%, N=84) had to take action in the past week to cover their basic needs, with the majority relying on government assistance (74%, N=67) or using savings (69%, N=62). The majority of respondents also reported a change in the type of foods (59%, N=52) and/or a reduction in the quantity of food that their family consumes daily (25%, N=22). Many respondents reported that their family was consuming less meat or vegetables since the pandemic began, often due to reduced income, rising food prices, or lack of availability in markets. There was a range in experiences across respondents in how the pandemic affected their food security. Some respondents reported relatively minor changes in food consumption:

**Table 4.** Reported changes and challenges associated with economic and food security as a result of COVID-19 or lockdowns.

|  | HOH/VWSC reporting<br>'yes'<br>(N=90) |
| --- | --- |
| **Job loss:** Have you or any member of your family lost a job or work due to coronavirus or the lockdown? | 28 (31%) |
| **Finances:** In the past 7 days, did you take any of the following actions to cover your household's basic needs? |  |
| Rely on government assistance | 67 (74%) |
| Use cash savings or bank savings | 62 (69%) |
| Borrow money or food | 36 (40%) |
| Rely on NGO assistance | 5 (6%) |
| Sell assets | 3 (3%) |
| At least one of the above | 84 (93%) |
| **Food security:** Has coronavirus or the lockdown changed the types or quantity of food your family eats daily? |  |
| Change in type of food | 52 (59%) |
| Decrease in quantity of food | 22 (25%) |
| Increase in quantity of food | 2 (2%) |
| No change in type or quantity of food | 34 (39%) |
| **Cooking fuel:** In the past 7 days, have you experienced any difficulties in getting the material you use to cook with (such as wood, gas, etc.) as a result of coronavirus or the lockdown? | 3 (3%) |

> *“I used to go out and eat street food, which is not possible now. And there are some items that we used to have in home, which are not available anymore. Mostly, the quantity has also reduced*.*”*
>
> *-Male respondent, 30 years old, Ganjam district (June 2020)*

However, others described more extreme reductions in food variety. For example, one respondent described how they are now only surviving on rice and water and two respondents similarly described how they used to eat a variety of foods before the pandemic, but since their income has dropped, they are now only surviving on rice, salt, and green chilies:

> *“There is some change in the eating practices. We received rice [ration from the government] which is fine, but the practice of eating has changed because there is no curry. Now, we have to eat green chili and salt. The government is helping but if we get some work, we can live a little more peacefully. We are eating green chili and salt in place of having a curry*.*”*
>
> *-Male respondent, 30 years old, Gajapati district (June 2020)*

Multiple respondents also explained that they had stopped eating meat due to a belief that COVID-19 could be spread through meat consumption:

> *“We have reduced eating fish and meat as there is a problem with fish and meat. These things would cause the [coronavirus] disease. We are eating less vegetables also. We have to eat rice and stir fried potato only*.*”*
>
> *-Male respondent, VWSC member, 61 years old, Ganjam district (June 2020)*

With regard to cooking fuel access, only three respondents reported difficulties in getting wood or gas as a result of the pandemic (3%, N=3). Among the 3 respondents who had difficulties, reasons included lack of finances, inability to travel during the pandemic, and not being given gas assistance (free gas or gas subsidy).

### Impacts on daily life and social interactions

Respondents were asked how their daily life had changed as a result of the pandemic and lockdowns, and many expressed that daily life had become more difficult (Table S5). One of the biggest changes was the experience of job loss or having less work due to the lockdown curfews and travel restrictions. Consequently, many also talked about the financial strain they or their household were facing. Another major change described was that everyone was staying at home more and ‘not going anywhere.’ Some respondents elaborated that this meant they or their family members no longer go to work or school, shop at the market less, do not visit family or friends, do not go to temple, or no longer take part in their normal leisure activities. Several HOH/VWSCs also reported less or restricted travel, such as not visiting other villages or nearby towns, and less ‘roaming around.’ Respondents also described how they were interacting with others less now and maintaining distance or taking precautions while interacting, which was confirmed at a village-wide level by most VWSC members. Finally, it was common for respondents to talk about how COVID-19 and the lockdowns had impacted their mental health, describing the different negative emotions they now felt in their daily life – experiencing a lot of anxiety and fear related to COVID-19, feeling worried and tense over the financial strain, and feeling bored or irritated from staying at home all the time. Some respondents also talked about feeling fearful of interacting with others or expressed how the reduced amount of social interaction was distressing for them. Many respondents touched on a multitude of these themes in their explanations:

> *“When coronavirus came to India from outside, people are facing issues. Whatever works were happening earlier are not happening now. People are also very anxious. Government is also giving some or other things, but will that be enough? There is a lot of expense. People are surviving with pain. The school is closed. Everything is closed*.*”*
>
> *-Male respondent, 40 years old, Ganjam district (June 2020)*
>
> *“Means, before coronavirus, our way of living, mingling with people was good. Earlier there was money also. Now, there is no work or income*.*”*
>
> *-Male respondent, 30 years old, Gajapati district (June 2020)*
>
> *“Earlier[…]we did not have any tension or fear, because any male member from the family could go out and work as laborers and earn money to ensure that the family is managed nicely and safely. Now, there is a challenge because nobody is able to work as a laborer and there is no income*.*”*
>
> *-Female respondent, 20 years old, Gajapati district (July 2020)*

In contrast to HOH and VWSC members who expressed that their daily life felt very different now, about a third of the caregivers explained that there was not much of a change in their own personal lives because they mostly stayed at home even before the lockdowns. While some HOH/VWSCs said they now have more free time, many caregivers noted that their housework and amount of free time remained the same:

> *“We stay at home. We always stay at home. How will life change then? That’s it. Work is also like how it always used to be. I never used to go anywhere*.*”*
>
> *-Female respondent, 30 years old, Ganjam district (July 2020)*

In addition, most caregivers explained that there was no change in their small child’s day-to-day life either, although many caregivers said they were limiting or no longer allowing their child to play outside the home and schools were now closed.

### Village response

In line with responses from individual respondents, the majority of VWSC members (81%, N=17) reported that most villagers took precautions to prevent coronavirus. Many (62%; N=13) reported holding a village-wide meeting as part of village-level actions, primarily to discuss COVID-19 prevention measures. Several VWSC members (33%, N=7) also reported taking precautionary measures against outsiders from the village, including physically barricading the village from outsiders or requiring those returning to stay in quarantine treatment centers before entering the village. Only one VWSC member said their village had taken no action to protect itself from coronavirus.

VWSC members were also asked questions about any outside assistance the village had received in relation to the pandemic. The majority of VWSC members (62%, N=13) said the government had provided guidance around what to do if someone is exposed or might be positive for coronavirus, often from an Anganwadi or ASHA worker. In contrast, when asked if the government or NGOs had provided any materials to the village, most of the respondents (86%, N=18) answered ‘no’, although a few described additional government rations or distribution of masks and soap. Despite the lack of materials provided, most VWSC members (71%, N=15) reported that there were not any resources or needs that their village required in relation to COVID-19 that had not been adequately addressed. However, two expressed issues with food access in their village and four stated their village needed masks and/or soap.

### Returned migrants

VWSC members were asked if there was any place for migrants coming back to the village to quarantine/isolate temporarily upon arrival. About half (43%, N=9) reported that there was a quarantine center in their village, which was typically at the school, while many others said there was a quarantine center nearby instead. Almost all VWSC members (90%, N=19) reported migrants had returned to their village or were currently in a quarantine center outside of the village. When asked how returned migrants were treated, most VWSC members expressed that villagers were fine with migrants returning to the village after staying in quarantine, although a couple VWSC members reported villagers feeling a little afraid.

Although we did not specifically target interviews with returned migrant workers, four of the head of households interviewed were migrant workers who had returned to their village due to the pandemic and stayed in quarantine centers, and some related themes arose naturally during their interviews. Two of the migrant workers faced extreme difficulties in returning home after the initial lockdown and expressed how panicked they felt during the experience. One of the migrant workers hired a bus with several other migrants to return home. However, he could only pay for the bus fare after his family took out a loan, because he was cheated out of all of his money by someone that promised to return him home and did not follow through. Although he was told to go to a police station in Surat to register for a train ride home, he did not do so out of fear. The other migrant worker explained that he started to walk home from Andhra Pradesh on his own and was later able to pay a truck driver to take him home. The migrant workers also described having a lack of money and work opportunities now that they have returned home. Additionally, one migrant worker described how depressing it was for him to face discrimination from his fellow villagers, who were afraid of talking to him now because he returned from a quarantine center.

## Discussion

The COVID-19 pandemic and related lockdowns had a substantial impact on villager’s daily life, social interactions, and economic and food security in rural Odisha, India. In particular, we found many individuals reported job loss, challenges related to lockdowns, changes and reductions in food consumed, and negative impacts on emotional well-being. In many instances, it was not possible to separate the impacts of the pandemic itself from the impacts of the government lockdowns. Additionally, there was high awareness and reported compliance with most important preventative measures, including staying home as much as possible, social distancing, washing hands frequently, and wearing a facial mask, suggesting that government restrictions and awareness campaigns were effective at changing behavior. Furthermore, experiences and challenges were similar between respondents across the two districts with distinct geographic and demographic characteristics, indicating our results may be generalizable to larger populations in Odisha or similar regions in other parts of rural India. The experiences captured by the interviews help document the lived experiences of villagers in rural Odisha during the pandemic, and can be useful for improving response to COVID-19 and future pandemics.

While respondents reported high compliance with preventative measures, we identified notable health knowledge gaps, which could be addressed in future awareness campaigns. Overall, one quarter of respondents were unable to name a correct symptom of COVID-19, one third did not know any high-risk groups, and less than half knew what causes the disease. Health knowledge about COVID-19 was notably lower among caregivers who may be more likely to care for sick or elderly household members. This suggests information campaigns targeted at these less-knowledgeable demographics could be beneficial, particularly door-to-door information campaigns to reach women and young caregivers who experience gendered restrictions on freedom of movement and interactions outside of the home. Those getting their main source of information from social media/internet and the news were also more likely to know a high-risk group and the disease cause, respectively, suggesting that greater education on these aspects from other sources like Anganwadi and ASHA workers could be beneficial. Despite some difficulties with accessing healthcare due to lockdowns and fear of contracting COVID-19, the study population generally reported strong health-seeking behavior, with the majority reporting they would go to a health clinic or hospital if they began experiencing symptoms. However, few said they would call a coronavirus help line set up by the government, indicating greater promotion of this resource may be needed. Our results showing high compliance with preventative measures despite low perceived personal risk of infection are generally in agreement with results from studies in other states of India (10,11). However, we found lower knowledge of COVID-19 symptoms compared to a study conducted in May 2020 in Tamil Nadu, India (11), which may be due in part to caregivers of young children being a specific target respondent in our study.

Many respondents reported struggles related to COVID-19, particularly related to economic and food security and mental health. Almost all respondents reported relying on some form of support to cover basic needs and almost a third reported job loss due to COVID-19. Although loss of jobs for daily wage workers was common in both areas, reported job loss in our study area was lower than a study in Bihar, India, where job loss effected almost two thirds of households (12). This may be because many respondents in our study worked in agriculture and were largely able to continue farming during the pandemic despite some struggles in selling their fruits and vegetables. Although many respondents reported receiving food rations from the government, reductions in dietary diversity and food quantity were commonly reported, which could lead to negative impacts on nutritional status and health (13,14). Additionally, migrant workers have been identified as a particularly vulnerable group in India during the pandemic (15). The experience of the few returned migrant workers we interviewed illustrated this vulnerability, including their extreme difficult returning home after job loss and facing discrimination. Mental health is also a substantial concern related to COVID-19, particularly as a result of job loss, financial strain, lockdowns, and fear of the illness (16–18). Many respondents reported declines in mental health as a result of the pandemic and feeling increased anxiety, fear, boredom, and irritation. Future pandemic response by governments and NGOs should consider these factors to prevent greater impacts on mental health.

This study had some important limitations. As it relied on phone interviews, we could only reach households with a mobile phone and network connection, which could potentially exclude the poorest and most remote households. However, we were able to talk to some respondents who lived in villages without a mobile network by connecting with them when they were outside the village in an area with network. Responses were also self-reported which may result in reporting bias, as respondents may over report practicing desirable hygienic behaviors, like handwashing (19–21). To try to reduce this bias and capture detailed experiences, the interview included several open-ended questions and asked follow-up explanations to closed-ended questions. Further findings related to handwashing and other WASH-related practices will also be reported in detail in a forthcoming paper. Additionally, although quantitative surveys have been used extensively to capture knowledge and impacts of the COVID-19 pandemic for large population samples, our use of open-ended, qualitative questions allowed us to explore the lived experiences of participants during this public health crisis more deeply than what can be captured by structured questions alone.

Overall, the study results help identify gaps in local capacity building for information providers, content of the awareness messaging, and channel of communication for message delivery to ensure accessibility to all. In rural Odisha, ASHA workers, Anganwadi workers, and auxiliary nurse midwifes (ANMs) typically serve as information providers on health matters. However, they are trained in-person periodically as per the demand of situations, which was not possible in this pandemic, and may have contributed to less reliance on these typical change agents as shown by our finding that less than half as many respondents reported getting their main source of information on the virus from Anganwadi or ASHA workers compared to the news. Additionally, the common message delivery mechanism of person-to-person communication was less feasible as people were more isolated during the pandemic, which may have resulted in a change in information channel to other sources for COVID-19 knowledge. There was also a gap in messaging content, which suggests that messaging needs to be designed keeping in perspective the broader aspects of health impacts such as physical health, mental health, and nutrition needs. Additionally, message delivery channels need to be made accessible to all, including women with limited access to the external world. As such, new delivery channels need to be explored for creating awareness other than the village meetings or in-person consultation and digital literacy may be helpful in this context.

Additionally, although the lockdowns and information campaigns appeared to be effective at changing villagers’ behavior to increase preventative measures taken, the lockdowns also had many adverse effects that should be mitigated during future responses. The evidence base created through understanding lived experiences during this pandemic can also help improve planning and preparedness at village and Gram panchayat (GP) administrative levels. The struggles that respondents reported related to difficulties meeting basic needs, accessing healthcare, experiencing negative emotions from staying home more, and police in some areas not allowing people to travel even for essential goods, demonstrate that administrative efficiency in dealing with the deterrent actions against the spread of virus could be improved. In particular, there is a need for greater government response to limit harm to livelihood and mental, social, and nutritional health during any lockdown periods required to reduce disease spread. Although there were already some government initiatives in place targeting economic and food security, our results show that there were still needs remaining for many rural villagers despite these programs and further initiatives may be needed. Overall, we identified high compliance with COVID-19 preventive actions despite shortcomings with messaging, many challenges and hardships faced in daily life due to the pandemic and lockdowns, all of which are insights that could be used to improve messaging and preparedness for future crisis response.

## Supporting information

Interview tools

## Data Availability

All relevant data are included within the manuscript and supplemental information. Further inquiries related to data access can be sent to Valerie Bauza.

## Acknowledgments

The authors thank Varsha Priyadarshini and Dhiren Swain for their help with data collection as well as Fiona Majorin, Sabrina Haque, Sheela Sinharoy, and Miles Kirby for their input related to the interview tool and mobile phone data collection plans. This research was supported in part by grants from the Bill & Melinda Gates Foundation (OPP1125067) and National Institute of Environmental Health Sciences, USA (T32ES012870 to VB).

## Author Roles

Conceptualization: VB, GS, TC

Formal Analysis: VB, GS, AO

Funding Acquisition: VB, GS, TC

Investigation: AB

Methodology: VB, GS, AB, AG

Writing – Original Draft Preparation: VB

Writing – Review & Editing: VB, GS, AB, AO, AG, TC

