## Supplemental Information for "Experience of the COVID-19 pandemic in rural Odisha, India: knowledge, preventative actions, and impacts on daily life"

**Sections:**
Section S1: Methods
Section S2: Results

**Tables:**
Table S1: Symptoms of COVID-19 reported by respondents.
Table S2: Sources for information about coronavirus for HOH and VWSC members.
Table S3: Respondent explanations by self-perceived risk level of contracting COVID-19.
Table S4: Respondent explanations for their level of concern if they or their family members contracted COVID-19.
Table S5. Qualitative themes related to changes in daily life and social interactions.

**Interview tools** (separate attachment)

**Methods** (Section S1)

Village and respondent selection procedures

The villages were a sub-set of villages enrolled in a child feces management (CFM) intervention trial. The trial villages included 74 randomly selected villages in Ganjam and Gajapati districts that met the following criteria: completed MANTRA program, 75% of households in village have access to a latrine, community water tank is functional with at least 75% of households connected to the piped water supply and 2 months or less of water shortage per year, village size between 35 to 250 households, village has its own Anganwadi center, and Gram Vikas has no programming planned in the village during the study period.

These 74 villages were then categorized into intervention and control villages among five distinct geographic-demographic ('geo-demo') groups that were used for village randomization in the CFM intervention trial, for ten distinct groups. For HOH and VWSC interviews, two villages within each of these ten village groups were randomly selected for inclusion for a total of 20 villages. 3 additional villages in Ganjam (1 from each Ganjam geo-demo group) were later added in June when a second strict lockdown went into place to try to capture the impact of that lockdown. We aimed to interview one VWSC member and three HOHs per village, across each of these 23 villages, and to interview 20 caregivers of children $<2$ years old and 20 caregivers of children $\geq 2$ years old. Target respondents in some remote villages were unreachable due to no or poor mobile networks. In some cases the HOH interview was completed by another household member requested by the HOH (such as the HOH's wife or son), and in a few cases the caregiver interview was completed by her husband when the caregiver was not fluent in Odia.

The list of phone numbers for all HOHs and VWSC members in the selected villages was provided by Gram Vikas and came from a household-level survey they had conducted about a year earlier. The list of phone numbers for caregivers came from the CFM intervention trial baseline survey conducted with households with a child <5 years old between December 2019 to February 2020.

Calling procedure

Each enumerator was assigned their own call lists, which included a list of phone numbers for a given respondent type residing in a certain village or set of villages. For example, a call list would include the phone numbers for all HOHs residing in village A. The phone numbers were randomly ordered and the enumerator was required to go down the list, calling each phone number at least twice and at different times of the day (once before and once after 5pm) if they could not connect with a respondent, before moving on to the next phone number. In some villages where cellular connection was poor, enumerators had to make many call attempts over several days before completing the desired number of surveys or having to move on to the next call list.

When a respondent answered the phone, the research assistant introduced themselves and briefly explained the purpose of the call. The research assistant then read a consent form that provided more details, including any risks and benefits to participating. The research assistant also asked the respondent for their consent to audio record the conversation. Once the respondent gave their consent, the research assistant continued with the interview questions. Throughout the phone call, the research assistant recorded answers in a Microsoft Word version of the interview. At the end of the call, the research assistant gave the respondent a list of helplines they could call if they needed assistance and also provided information on preventative measures to protect themselves against COVID-19. After the call, research assistants listened to the audio recording and completed detailed notes.

Target respondents in some villages were unable to be reached due to lack of mobile network in more remote villages. If one of the selected villages did not have adequate network to complete the surveys (based on calling all numbers and asking Gram Vikas about mobile network connectivity in the village), then the next random village was selected. If a VWSC member was not able to be reached in a village after several attempts and reaching out to Gram Vikas for additional contact information, then a VWSC member from a nearby village in the same geo-demo group was contacted.

Additionally, in two of the three additional Ganjam villages that were added after the second lockdown, we had difficulty with response due to poor relations of the village with Gram Vikas. After assessing the limited data collected in these two villages, it appeared that no new themes were emerging due to the second lockdown, so we did not reassign replacement villages for the two villages that we could not fully complete the target surveys in.

Categorizing correct symptoms and causes of COVID-19

We categorized symptoms and causes of COVID-19 given by respondents as correct if they matched COVID-19 information available from any of the following organizations: WHO,[1] CDC,[2] Government of India,[3] Government of Odisha.[4]

**Results** (Section S2)

Knowledge about potential COVID-19 treatment. When HOH and VWSC respondents were asked whether there was a cure or treatment for COVID-19, 46% of respondents (N=42) said yes, 32% (N=29) reported they did not know, and 22% (N=21) said no. However, even for the respondents who said there was no cure, several still mentioned various treatments. In general, most respondents described preventative measures as a treatment such as isolation, proper hygiene, and mask wearing. Four respondents stated the drug 'Chloroquine', commonly used as an anti-malarial, was a treatment. Several respondents (N=8) did not mention a specific treatment but were sure there must be one given the low lethality of individuals infected.

Table S1. Symptoms of COVID-19 reported by respondents (asked as an open-ended question).

| *What are the symptoms of the coronavirus?* | **All (N=125)** | **HOH (N=68)** | **VWSC (N=20)** | **Caregiver (N=37)** |
|---|---|---|---|---|
| Cough* | 88 (68%) | 53 (74%) | 19 (90%) | 16 (43%) |
| Fever* | 55 (42%) | 28 (39%) | 14 (67%) | 13 (35%) |
| Sneezing | 30 (23%) | 20 (28%) | 4 (19%) | 6 (16%) |
| Chills* | 24 (18%) | 13 (18%) | 11 (52%) | 0 |
| Headache* | 14 (11%) | 10 (14%) | 3 (14%) | 1 (3%) |
| Difficulty breathing* | 8 (6%) | 4 (6%) | 3 (14%) | 1 (3%) |
| Sore throat* | 6 (5%) | 3 (4%) | 2 (10%) | 1 (3%) |
| Chest pain* | 5 (4%) | 3 (4%) | 2 (10%) | 0 |
| Tiredness* | 4 (3%) | 1 (1%) | 3 (14%) | 0 |
| Body aches* | 3 (2%) | 2 (3%) | 1 (5%) | 0 |
| Diarrhea* | 3 (2%) | 1 (1%) | 2 (10%) | 0 |
| Loss of taste or smell* | 1 (1%) | 0 | 1 (5%) | 0 |
| Other | 14 (11%) | 6 (8%) | 7 (33%) | 1 (3%) |
| No symptoms | 1 (1%) | 1 (1%) | 0 | 0 |
| Don't know | 29 (22%) | 11 (15%) | 1 (5%) | 17 (46%) |

*Correct symptom of COVID-19 based on guidance from WHO, CDC, Gov. of India, and Gov. of Odisha.

Table S2. Sources for information about coronavirus for HOH and VWSC members.

| *What has been your main source for information about coronavirus?*<br>*From what other sources have you received information about coronavirus?* | **Main source (N=90)** | **All sources* (N=90)** |
|---|---|---|
| Government (National) | 1 (1%) | 41 (46%) |
| Government (State/Local) | 5 (6%) | 55 (61%) |
| Village leader | 3 (3%) | 41 (46%) |
| NGO | 1 (1%) | 11 (12%) |
| Anganwadi worker | 23 (26%) | 74 (82%) |
| ASHA worker | 19 (21%) | 71 (79%) |
| Other health worker | 7 (8%) | 28 (31%) |
| Neighbor in your village | 5 (6%) | 24 (27%) |
| Friend | 12 (13%) | 43 (48%) |
| Family | 4 (4%) | 35 (39%) |
| Social media/internet | 26 (29%) | 36 (40%) |
| News | 59 (66%) | 67 (74%) |
| Other | 8 (9%) | 12 (13%) |

* Includes both the main source reported and other sources.

Table S3. Respondent explanations by self-perceived risk level of contracting COVID-19.*

| **Perceived risk level** | **Qualitative explanations** |
|---|---|
| No risk<br>(56%, N=74) | • **Taking precautions (staying home, handwashing, mask-wearing, social distancing)**<br>• **Taking general precautions (no specific precautions described)**<br>• **No local cases**<br>• **Feels they are in good health or low risk**<br>• Taking homeopathic precautions (consuming warm water/food)<br>• Tested negative for COVID-19, lack of COVID-19 symptoms<br>• Depends on luck, hoping to not be infected<br>• Does not believe in the existence of COVID-19 (1 respondent) |
| Low risk<br>(5%, N=7) | • No local cases<br>• Taking precautions, being alert<br>• Feels small risk because of news of transmission or devastation in other countries |
| Medium risk<br>(4%, N=5) | • **Easy spread of coronavirus, transmission by contact with others**<br>• Surge in cases<br>• Feels risk because they have asthma<br>• Praying to not be infected |
| High risk<br>(5%, N=6) | • **Widespread transmission of coronavirus, including in nearby villages**<br>• Lack of testing of asymptomatic individuals<br>• Feels risk because they are elderly<br>• Instability of India |
| Don’t know<br>(27%, N=35) | • **Uncertainty of virus (always a chance of getting infected, hard to predict, who can say?)**<br>• **Stays home**<br>• **Does not know ‘anything’ about coronavirus**<br>• Easy spread, fluctuating local cases<br>• Taking homeopathic precautions (keeping their blood warm)<br>• Depends on luck, praying to not be infected |

* Bolded items are the more common explanations

Table S4. Respondent explanations for their level of concern if they or their family members contracted COVID-19.*

| Level of Concern | Qualitative explanations |
|---|---|
| Not at all concerned (5%, N=4) | • Family not considered high risk<br>• Feel comfortable dying (as they are spiritual, retired and their children have been married)<br>• Would call government helpline |
| Somewhat concerned (13%, N=11) | *Explanations that indicate more concerned*<br>• Rapid spread of the virus<br>• Don't know much about the virus<br>• Difficult to recognize symptoms<br>• Stigma surrounding infection<br>• Would need to seek healthcare<br>• Concerned because they are their family<br>*Explanations that indicate less concerned*<br>• High recovery rate but inevitable infection<br>• Just need to get the infected person proper healthcare<br>• Government has provided facilities |
| Very concerned (77%, N=68) | • **Transmission to other family members**<br>• **Concerned because they are their family**<br>• **Virus lethality, severity**<br>• Lack of vaccine/cure<br>• Virus uncertainty (novel disease)<br>• Difficult to recognize symptoms<br>• Family not present, would need to figure out who cares for the sick<br>• Lack of hospital accessibility, need for medical assistance<br>• Lack of money<br>• Placed faith in God<br>• Stigma surrounding infection |
| Don't know (6%, N=5) | • Difficult to recognize symptoms<br>• It may happen, it may not<br>• Cannot do anything about infection<br>• Following safety precautions† |

*Bolded items are the more common explanations

†Participant explained why they are less concerned instead of why they don't know

Table S5. Qualitative themes related to changes in daily life and social interactions.

| Interview Question | Qualitative Themes and Descriptions |
|---|---|
| How has coronavirus or the lockdown changed your daily life? Think about from the time you get up in the morning to the time you go to sleep at night. How is your day different now, if it is different? | • Less or no work and economic difficulties: Several respondents explained that they or a family member had less work or no work now due to COVID-19. Many also explained economic problems as a result of COVID-19. Many described how their work has stopped now and some described not being able to work normally due to the curfew or no longer being able to work outside of the state now. Many respondents also described about how they have less money now, how they are getting less money now for their work, or how they have used their money up. Some people said that things are more expensive now. Multiple respondents also explained that they were unable to sell their fruits or vegetables due to the lockdown and that they lost a lot of money due to that. A couple of respondents also talk specifically about government help, one saying that the benefits are not enough and another saying that the government has said that they will be paid 1000-2000 INR but that they have not gotten that money.<br>• Staying home more: Many respondents explained that they are now spending more time staying at home. Some explained that this was due to less work, whereas others discussed it as a precaution or due to curfew or lockdown. People also explained that they are no longer engaging in their normal leisure activities where they would gather and socialize with others outside of the home. A couple of respondents explained that they can no longer visit the temple in their village. Some respondents also explained that their children's or other children's school or college is closed due to coronavirus.<br>• Less or restricted travel or movement: Several respondents reported less travel and movement due to COVID-19 or lockdowns, including less roaming around. Some respondents specifically explained that they are not able to travel to another village or go to a nearby town or city. Some said that this was due to lockdowns, whereas others said that it was due to coronavirus more generally. Multiple respondents also reported fear of the police or law enforcement when they do out, including getting beaten by the police.<br>• Anxiety, thoughts of coronavirus, and other negative emotions: Many respondents explained that they have anxiety, think a lot about coronavirus, or have other negative emotions due to COVID-19. Some respondents described how daily life now being full of anxiety or fear. While some respondents report fear/anxiety about getting the disease, others described fear of going out now or that caution is always on their minds. Some respondents also said that they spend a lot of time thinking about the disease or that it is always on their mind. Multiple respondents expressed that they were happier before the pandemic or that they are sad or bored now.<br>• Daily life is very different: Many respondents described life being very different or having changed a lot now. This was often due to things brought up in other themes such as job loss, changes in social interactions, and reduced travel, school/college closings, or anxiety related to COVID-19. A couple respondents also talk about how many people have returned to the village now. Alternatively, about a third of caregivers and a few men working as farmers explained that there was little to no change in their daily life. For caregivers, this was usually because they "never went out" even before COVID-19/lockdowns. |

| | |
|---|---|
| | • More free time: Some HOH/VWSC respondents explained they have more free time due to COVID-19 because their work or other activities have stopped. Some also explained they now are spending more time with their family or friends, or that they have more time to sleep now.<br><br>• Minimal change in caregiver's household work: Among caregivers who commented on their workload/freetime, most explained that their housework/workload had remained the same because they had always stayed at home and did the housework anyways. However, a couple caregivers mentioned having more housework now since everyone was now at home or because housework is the only thing to do when stuck at home. A couple caregivers also mentioned having less housework now since there were no visitors now and everyone was at home. |
| Has coronavirus or the lockdown changed how you or your family interact with other members of your village? Please explain. | • Reduced interactions: Many respondents described interacting less with others. Some described staying in their home or staying away from other people as much as possible. Others described that they are no longer gathering or meeting in groups to hang out or that they are not going over to other people's homes. Some people also described fear associated with interacting, or that the reduced social interaction is distressing for them. One respondent specifically said that marriages, religious ceremonies, festivals, and village committee meetings cancelled.<br><br>• Maintain distance when interacting: Several respondents explained that they are maintaining distance when they interact with other people. Some respondents also described that they are conscious of the disease while talking to others and/or taking precautions such as not shaking hands.<br><br>• Cautious/careful/restrictions when interact: A few respondents explained that interactions have changed and they are being cautious or careful when they interact with others or that interactions have restrictions, but did not provide any specifics to fit into the above two categories.<br><br>• No change: 18% of respondents (N=22) reported no change in social interactions with other people in their village. |
