## Supplementary material for "Experience of the COVID-19 pandemic in rural Odisha, India: knowledge, preventative actions, and impacts on daily life": Interview tools

| **SECTION A: Respondent and Enumerator Information & Consent Process** | |
|---|---|
| **A1.** Village ID ଗ୍ରାମ ର କୋଡ: | **A2.** Household ID ଘଠ ଘରର ସଂଖ୍ୟା |
| **A3.** Village Name ଗ୍ରାମ ର ନାମ | |
| **A4.** Date ତାରିଖ | (y/d/m) |
| **A5.** Start Time ଆରମ୍ଭ ସମୟ | |
| **A6.** Enumerator name ଆପଣଙ୍କ ନାମ | ☐ 1. Alok<br>☐ 2. Dhiren<br>☐ 3. Varsha |
| **A7.** Confirm that the respondent and their family (if applicable) currently live in the intended village (and are not currently staying somewhere else).<br><br>ପୁଷ୍ଟି କରନ୍ତୁ କି ଯେ, ଉପଯୁକ୍ତ ଉତ୍ତରଦାତା ଏବଂ ତାଙ୍କ ପରିବାର (ଯଦି ଲାଗୁ ହୁଏ)ବର୍ତ୍ତମାନ ଉଦ୍ଦିଷ୍ଟ ଗ୍ରାମରେ ହିଁ ବସବାସ କରନ୍ତି (ଏବଂ ଅନ୍ୟ କେଉଁଠି ବସବାସ କରନ୍ତି ନାହିଁ) | ☐ 1. Yes ହଁ<br>☐ 2. No – respondent/family are staying outside the village and left *before* the pandemic → **END SURVEY**<br>ନା - ଉତ୍ତରଦାତା ଓ ତାଙ୍କ ପରିବାର ଗ୍ରାମ ବାହାରେ ବସବାସ କରନ୍ତି ଏବଂ ସର୍ବବ୍ୟାପୀ ମହାମାରୀ **ହେବା *ପୂର୍ବରୁ*** ଗ୍ରାମ ଛାଡି ଦେଇଛନ୍ତି - **ସର୍ଭେ ଶେଷ**<br>☐ 3. No – respondent/family are staying outside the village and left *because of* the pandemic → **END SURVEY**<br>ନା- ଉତ୍ତରଦାତା ଓ ତାଙ୍କ ପରିବାର ଗ୍ରାମ ବାହାରେ ବସବାସ କରନ୍ତି ଏବଂ ସର୍ବବ୍ୟାପୀ ମହାମାରୀ **ହେବା *କାରଣରୁ*** ଗ୍ରାମ ଛାଡି ଦେଇଛନ୍ତି - **ସର୍ଭେ ଶେଷ** |
| **A8a.** Read the consent form to the respondent and then ask for their consent to participate.<br><br>ଉତ୍ତରଦାତାଙ୍କ ପାଇଁ ସହମତି ପତ୍ରଟିକୁ ପଢନ୍ତୁ ଏବଂ ତାଙ୍କୁ **ସର୍ଭେରେ** ଅଂଶଗ୍ରହଣ ପାଇଁ ସହମତି ମାଗନ୍ତୁ।<br><br>Does the respondent consent to the survey?<br>କ'ଣ ଉତ୍ତରଦାତା ସର୍ଭେରେ ଅଂଶଗ୍ରହଣ ପାଇଁ ସହମତି ପ୍ରଦାନ କଲେ? | ☐ 1. Yes ହଁ<br>☐ 2. No → **END SURVEY** ନା - **ସର୍ଭେ ଶେଷ** |
| **A8b.** Does the respondent consent to having the phone call audio recorded?<br><br>କଣ ଉତ୍ତରଦାତା ଫୋନକଲର ରେକଡିଙ୍ଗ ପାଇଁ ସହମତି ପ୍ରଦାନ କଲେ? | ☐ 1. Yes ହଁ<br>☐ 2. No ନା → Pause when you need to in order to record detailed responses! ଯେତେବେଳେ ଉତ୍ତର ଲେଖିବା ଆବଶ୍ୟକ ପଡେ, ବିରାମ ନିଅନ୍ତୁ। |
| **ENUMERATOR NOTE:** Now you are going to begin the survey.<br>ଏନୁମରେଟରଙ୍କ ପାଇଁ ନୋଟ: ବର୍ତ୍ତମାନ ଆପଣ ସର୍ଭେ ଆରମ୍ଭ କରିବେ | |
| **A9.** What is your name?<br>ଆପଣଙ୍କ ନାମ କଣ ? | |

### COVID-19 Mobile Phone Interview – Head of Household

| | |
|---|---|
| **A10.** What is the sex of the respondent?<br>ଉତ୍ତରଦାତଙ୍କ ଲିଙ୍ଗ କଣ?<br><br>Enumerator Note: Clarify with the respondent if you are not sure.<br>ଏନୁମରେଟର ନୋଟ - ସ୍ୱଷ୍ଟ କରନ୍ତୁ ଯେବେ ଆପଣ ନିଶ୍ଚିତ ନୁହନ୍ତି | ☐ 1. Male ପୁରୁଷ<br>☐ 2. Female ମହିଳା |
| **A11.** What is your age?<br>ଆପଣଙ୍କ ବୟସ କେତେ? | (years) (ବର୍ଷ) |
| **A12.** Including yourself, how many people are currently staying in your house?<br>ଆପଣଙ୍କୁ ମିଶେଇ, ବର୍ତ୍ତମାନ ଆପଣଙ୍କ ଘରେ କେତେଜେଣ ଲୋକ ରୁହନ୍ତି? | |
| **A13.** Are there any children <5 years currently living in your household?<br>କଣ ପାଞ୍ଚ ବର୍ଷରୁ କମ ବୟସର କେହି ପିଲା ବର୍ତ୍ତମାନ ଏହି ଘରେ ବସବାସ କରନ୍ତି କି? | ☐ 1. Yes ହଁ<br>☐ 2. No ନାହିଁ<br><br>**<u>If yes, children ages:</u>**<br>Child 1: Years: ________ Months: _________<br>Child 2: Years: ________ Months: _________<br>Child 3: Years: ________ Months: _________ |
| **A14.** How many people living in your household are over 60 years old?<br><br>ଆପଣଙ୍କ ଘରେ ରହୁଥିବା ସଦସ୍ୟ ମାନଙ୍କ ମଧ୍ୟରୁ କେତେ ଜଣଙ୍କ ବୟସ ୬୦ ବର୍ଷରୁ ଉର୍ଦ୍ଧ୍ୱ? | |
| **A15.** How many people living in your household are currently pregnant?<br><br>ଆପଣଙ୍କ ଘରେ ରହୁଥିବା ସଦସ୍ୟ ମାନଙ୍କ ମଧ୍ୟରୁ କେତେ ଜଣ ଗର୍ଭବତୀ? | |
| **A16.** Is anyone in your household a daily wage labourer who usually works in another state?<br><br>କଣ ଆପଣଙ୍କ ଘରର କେହି ଦିନ ମଜୁରିଆ ଅଟନ୍ତି କି ଯେ ସାଧାରଣତଃ ଅନ୍ୟ ରାଜ୍ୟରେ କାମ କରନ୍ତି ? | ☐ 1. Yes ହଁ - And they are still residing in another state ହଁ- ସେମାନେ ବର୍ତ୍ତମାନ ମଧ୍ୟ ଅନ୍ୟ ରାଜ୍ୟରେ ରହୁଛନ୍ତି<br>☐ 2. Yes ହଁ - But now they reside in Odisha ହଁ- କିନ୍ତୁ ସେମାନେ ବର୍ତ୍ତମାନ ଓଡିଶାରେ ରୁହନ୍ତି<br>☐ 3. No ନା |

| | |
|---|---|
| **A17.** Do you or any other family member have any of the following conditions or diseases?<br><br>ଆପଣଙ୍କୁ ଅଥବା ଆପଣଙ୍କ ପରିବାରର କେହି ସଦସ୍ୟଙ୍କୁ ଉକ୍ତ ରୋଗ ଅଛି କି?<br><br>Enumerator note: Read each option and select all that apply.<br>ଏନୁମରେଟରଙ୍କ ପାଇଁ ନୋଟ: ବିକଳ୍ପ ଉତ୍ତର ଗୁଡିକୁ **ପଢନ୍ତୁ।** ଦେଇଥିବା ସମସ୍ତ ଉତ୍ତରକୁ ବାଛନ୍ତୁ | ☐ 1. Asthma ଶ୍ୱାଶ<br>☐ 2. Diabetes ମଧୁମେହ<br>☐ 3. Obesity ମୋଟାପା<br>☐ 4. Kidney disease ବୃକକ ରୋଗ<br>☐ 5. Heart disease ହୃଦରୋଗ<br>☐ 6. Liver disease ଯକୃତ ରୋଗ<br>☐ 7. Chronic lung disease ଦୀର୍ଘକାଳୀନ ଫୁସଫୁସ ରୋଗ<br>☐ 8. Cancer କ୍ୟାନସର<br>☐ 9. None କିଛି ନାହିଁ |
| **A18.** Do you have piped water to your household or compound?<br><br>କଣ ଆପଣଙ୍କ ଘରକୁ ପାଇପ ଯୋଗେ ପାଣିର ବ୍ୟବସ୍ଥା ଅଛି? | ☐ 1. Yes – functional ହଁ- ସକ୍ରିୟ<br>☐ 2. Yes – not functional ହଁ- ନିଷ୍କ୍ରିୟ<br>☐ 3. No ନା<br><br>**<u>If 2 or 3:</u>**<br>What is your primary source of water and where is it located?<br><br><u>ଯଦି 2 କିମ୍ବା 3:</u><br>ଆପଣଙ୍କ ଘରର ମୁଖ୍ୟ ଜଳଉତ୍ସଟି କଣ ଓ ଏହା କେଉଁ ଠାରେ ଅବସ୍ଥିତ?<br><br>Source (ଉତ୍ସ):<br>☐ 1. Public tap ପବ୍ଲିକକଳ/ସର୍ବସାଧାରଣଟ୍ୟାପ୍<br>☐ 2. Well/borehole/spring କୂଅ, ନଳକୂଅ, ଝରଣା<br>☐ 3. Surface water (river/dam/lake/pond/stream/canal) ଭୂତଳଜଳ(ନଦୀ/ବନ୍ଧ/ହ୍ରଦ/ପୋଖରୀ/ନାଳ/କେନାଲ)<br>☐ 4. Rainwater ବର୍ଷାପାଣି<br>☐ 5. Bottled water ବୋତଲପାଣି<br>☐ 6. Other ଅନ୍ୟାନ୍ୟ<br>☐ 7. Don't know ଜାଣିନାହିଁ<br><br>Location: ଅବସ୍ଥାନ<br>☐ 1. In own dwelling ନିଜ ରହିବା ଜାଗାରେ<br>☐ 2. In own yard/plot/compound ନିଜଜାଗା/ପ୍ଲଟରେ/ଅଗଣାରେ<br>☐ 3. Outside the household compound ଘର ଅଗଣା/ପରିସର ବାହାରେ |
| **A19.** Does your household have a latrine?<br><br>କଣ ଆପଣଙ୍କ ଘରେ ପାଇଖାନା ଅଛି? | ☐ 1. Yes – functional ହଁ- ସକ୍ରିୟ<br>☐ 2. Yes – not functional ହଁ- ନିଷ୍କ୍ରିୟ<br>☐ 3. No ନାହିଁ |
| **A20.** Do you have an enclosed bathing area in your household compound?<br><br>କଣ ଆପଣଙ୍କ ଘର ପରିସର ମଧ୍ୟରେ ଏକ ଆବଦ୍ଧ ଗାଧୁଆଘର ଅଛି କି? | ☐ 1. Yes ହଁ<br>☐ 2. No ନା |

### COVID-19 Mobile Phone Interview – Head of Household

#### SECTION B: COVID-19 Knowledge, Perceptions, and Actions

**Enumerator read:** Thank you for your responses so far. Now I’m going to ask you some questions related to coronavirus.

ଏନୁମରେଟର ପଢନ୍ତୁ : ବର୍ତ୍ତମାନ ପର୍ଯ୍ୟନ୍ତ ଦେଇଥିବା ଉତ୍ତର ମାନଙ୍କ ପାଇଁ ଧନ୍ୟବାଦ। ବର୍ତ୍ତମାନ, ମୁଁ ଆପଣଙ୍କୁ କରୋନାଭାଇରସ ସହିତ ଜଡିତ କିଛି ପ୍ରଶ୍ନ ପଚାରିବି।

| | |
|---|---|
| **B1.** Have you heard of a disease called covid-19, coronavirus, or corona?<br><br>କଣ ଆପଣ କୋଭିଡ-୧୯, କରୋନା-ଭାଇରସ କିମ୍ବା କରୋନା ନାମକ ରୋଗ ବିଷୟରେ ଶୁଣିଛନ୍ତି? | ☐ 1. Yes ହଁ<br>☐ 2. No → **If no, explain that this is the disease that has led to the recent lockdowns in India.**<br>ନା- ଯଦି ନା, ତାହେଲେ ଏହା କୁହନ୍ତୁ କି ଯେ ଏହି ରୋଗ ପାଇଁ ହିଁ ଏବେ ଭାରତରେ ଲକ-ଡାଉନ ହୋଇଛି |
| **B2.** What are the symptoms of coronavirus?<br><br>କରୋନା ଭାଇରସର ଲକ୍ଷଣ ଗୁଡିକ କଣ?<br><br>Enumerator notes:<br>Do **NOT** read response options. Select all that apply.<br><br>ସମ୍ଭାବ୍ୟ ଉତ୍ତର ଗୁଡିକୁ ପଢନ୍ତୁ ନାହିଁ. ଦେଇଥିବା ଉତ୍ତର ଗୁଡିକୁ ଟୀକ କରନ୍ତୁ.<br><br>Probe: Anything else?<br>ପ୍ରୋବ : ଆଉ କିଛି?<br><br>If respondent says sick/flu as their response, ask them to specify specific symptoms.<br><br>ଯଦି ଉତ୍ତରଦାତା ଅସୁସ୍ଥ ଲାଗିବା/ ଥଣ୍ଡା ହେବା ବିଷୟରେ କୁହନ୍ତି, ତାଙ୍କୁ ନିର୍ଦ୍ଦିଷ୍ଟ ଲକ୍ଷଣ ଗୁଡିକ ବିଷୟରେ କହିବାକୁ କୁହନ୍ତୁ | ☐ 1. No symptoms କିଛି ଲକ୍ଷଣ ନାହିଁ<br>☐ 2. Fever ଜ୍ୱର<br>☐ 3. Cough କାଶ<br>☐ 4. Dry cough ଶୁଖିଲା କାସ<br>☐ 5. Headache ମୁଣ୍ଡବଥା<br>☐ 6. Body aches ଶରୀର ବ୍ୟଥା<br>☐ 7. Difficulty breathing ନିଶ୍ୱାସ ନେବାରେ ଅସୁବିଧା<br>☐ 8. Chest pain ଛାତି ଯନ୍ତ୍ରଣା<br>☐ 9. Tiredness କ୍ଲାନ୍ତି<br>☐ 10. Diarrhea ଡ଼ାଇରିଆ<br>☐ 11. Chills ଥଣ୍ଡା ଲାଗିବା<br>☐ 12. Rash କୁଣ୍ଡିଆ<br>☐ 13. Loss of smell and/or taste ଗନ୍ଧ କିମ୍ବା ସ୍ୱାଦ ବାରିପାରୁନାହାନ୍ତି<br>☐ 14. Dizziness ମୁଣ୍ଡ ବୁଲେଇବା<br>☐ 15. Sneezing ଛିଙ୍କ/ଜିଙ୍କ<br>☐ 16. Sore throat ଖରାପ ଗଳା<br>☐ 88. Other ଅନ୍ୟାନ୍ୟ:<br>☐ 99. Don't know ଜାଣିନାହିଁ |
| **B3.** What causes a person to get sick with coronavirus?<br><br>କେଉଁ କାରଣ ମାନଙ୍କ ପାଇଁ ଜଣଙ୍କୁ କରୋନା ଭାଇରସ ହୋଇଥାଏ?<br><br>Enumerator note: Probe for respondent to describe in detail. If the respondent is having trouble answering, ask: “What actions would increase your risk of getting coronavirus? Why?”<br>ଏନୁମରେଟରଙ୍କ ପାଇଁ ନୋଟ: ଉତ୍ତରଦାତାଙ୍କୁ ବର୍ଣ୍ଣନା କରିବା ପାଇଁ ଅନୁସନ୍ଧାନ କରନ୍ତୁ। ଯଦି ଉତ୍ତରଦାତାଙ୍କୁ ଉତ୍ତର ଦେବାରେ ଅସୁବିଧା ହେଉଥାଏ, ପଚାରନ୍ତୁ, “କେଉଁ କମ ଆପଣଙ୍କର କରୋନାଭାଇରସ ହେବାର ଆଶଙ୍କାକୁ ବଢେଇଦେଇଥାଏ? କାହିଁକି?” | |

### COVID-19 Mobile Phone Interview – Head of Household

| | |
|---|---|
| **B4.** Are there any cures or treatments for coronavirus disease?<br>କ'ଣ କରୋନା ଭାଇରସର କିଛି ଉପଶମ କିମ୍ବା ଚିକିତ୍ସା ଅଛି କି?<br><br>If yes, please describe these cures or treatments.<br>ଯଦି ହଁ, ଏହି ଉପଶମ କିମ୍ବା ଚିକିତ୍ସା ଗୁଡିକୁ ବର୍ଣ୍ଣନା କରନ୍ତୁ<br><br>Enumerator note: If the respondent describes their answer, please take notes on their response even if they respond 'no' or 'don't know'.<br><br>ଏନୁମରେଟର ନୋଟ : ଯଦି ଉତ୍ତରଦାତା ତାଙ୍କ ଉତ୍ତରକୁ ବର୍ଣ୍ଣନା କରନ୍ତି, ତାଙ୍କ ଉତ୍ତରକୁ ଟିପନ୍ତୁ ଯଦିଓ ସେମାନେ 'ନାହିଁ' କିମ୍ବା 'ଜଣା ନାହିଁ' ଉତ୍ତର ଦିଅନ୍ତି। | ☐ 1. Yes ହଁ<br>☐ 2. No ନାହିଁ<br>☐ 99. Don't know ଜଣାନାହିଁ<br><br>**<u>Please describe:</u>**<br>**<u>ଦୟାକରି ବର୍ଣ୍ଣନା କରନ୍ତୁ</u>** |
| **B5.** Do you think your personal chance of getting coronavirus is low, medium, high, or do you have no risk at all? Explain why.<br><br>ଆପଣଙ୍କୁ ଲାଗୁଛି କି ଯେ ଆପଣଙ୍କୁ କରୋନାଭାଇରସ ହେବାର ବ୍ୟକ୍ତିଗତ ସମ୍ଭାବନା କମ, ମଧମ, ଅଧିକ ଅଥବା ଆପଣଙ୍କ ପ୍ରତି କିଛି ବି ବିପଦ ନାହିଁ? ବର୍ଣ୍ଣନା କରନ୍ତୁ କାହିଁକି। | ☐ 1. Low କମ<br>☐ 2. Medium ମାଧ୍ୟମ<br>☐ 3. High ଅଧିକ<br>☐ 4. No risk କିଛି ବିପଦ ନୁହେଁ<br>☐ 99. Don't know ଜାଣିନାହିଁ<br><br>**<u>Explain why: ବର୍ଣ୍ଣନା କରନ୍ତୁ କାହିଁକି</u>** |
| **B6.** In the past 7 days, what actions have you and your family taken to avoid getting coronavirus, if any?<br><br>ଗତ ସାତଦିନ ମଧ୍ୟରେ, ଆପଣ ଓ ଆପଣଙ୍କ ପରିବାରର ସଦସ୍ୟ ମାନେ ନିଜକୁ କରୋନା ଭାଇରସରୁ ଦୂରେଇ ରଖିବାକୁ କଣ ପଦକ୍ଷେପ ନେଇଛନ୍ତି? | |
| **B7.** In the past 7 days, what actions have you and your family taken to avoid getting coronavirus, if any? I am going to read a list of actions. For each one, please tell me yes or no.<br><br>ଗତ ସାତ ଦିନ ମଧ୍ୟରେ, ଆପଣ ଓ ଆପଣଙ୍କ ପରିବାରର ସଦସ୍ୟ ମାନେ ନିଜକୁ କରୋନାଭାଇରସରୁ ଦୂରେଇ ରଖିବାକୁ କଣ ପଦକ୍ଷେପ ନେଇଛନ୍ତି? ମୁଁ କିଛି ପଦକ୍ଷେପ ପଢୁଛି। ପ୍ରତିଟି ପଦକ୍ଷେପ ପାଇଁ ହଁ କିମ୍ବା ନାହିଁ କୁହନ୍ତୁ।<br><br>Enumerator note: Select each action that the | ☐ 1. Stayed at home as much as possible<br>ଯେତେ ପାରିଛନ୍ତି, ଘରେ ରହିଛନ୍ତି<br>☐ 2. Did not attend school or work<br>ସ୍କୁଲ କିମ୍ବା କାମକୁ ଯାଇନାହାନ୍ତି<br>☐ 3. Did not attend social gatherings (e.g., weddings, funerals, church, temple)<br>ସାମାଜିକ ସମାବେଶକୁ ଯାଇନାହାନ୍ତି (ଉଦାହରଣ : ବିବାହ,ଅନ୍ତିମ ସଂସ୍କାର, ଗୀର୍ଜାଘର, ମନ୍ଦିର)<br>☐ 4. Kept a distance from others → What distance? ____ meters<br>ଅନ୍ୟ ମାନଙ୍କ ଠାରୁ ଦୂରତ୍ୱ ରକ୍ଷା କରିଛନ୍ତି – କେତେ ମିଟର ଦୁରତ୍ୱ<br>☐ 5. Washed hands/used hand sanitizer more frequently |

| | |
|---|---|
| respondent says 'yes' for.<br><br>ଏନୁମରେଟରଙ୍କ ପାଇଁ ନୋଟ: ସବୁ ପଦକ୍ଷେପ ବାଛନ୍ତୁ, ଯାହାପାଇଁ ଉତ୍ତରଦାତା 'ହଁ' କହିଛନ୍ତି । | ହାତ ଧୋଇବା / ସାନିଟାଇଜର ର ବ୍ୟବହାର ବଢ଼େଇ ଦେଇଛନ୍ତି<br>☐ 6. Wore a (face) mask<br>ମାସ୍କ ପିନ୍ଧିଛନ୍ତି<br>☐ 7. Wore gloves (on your hands)<br>ଗ୍ଲୋବ୍ସ ପିନ୍ଧିଛନ୍ତି<br>☐ 8. Tried to stop touching face<br>ମୁହଁଟିକୁ ନ ଛୁଇଁବାର ଚେଷ୍ଟା କରିଛନ୍ତି<br>☐ 9. Did not shake hands with others<br>ଅନ୍ୟ ମାନଙ୍କ ସହିତ ହାତ ମିଶେଇନାହାନ୍ତି<br>☐ 10. Covered your mouth with elbow when you sneezed or coughed<br>ଛିଙ୍କିବା କିମ୍ବା କାଶିବା ସମୟରେ ପାଟିକୁ କହୁଣୀରେ ଘୋଡାଇ ଦେଇଛନ୍ତି<br>☐ 11. Drank local alcohol ଦେଶୀ ମଦ ପିଇଛନ୍ତି<br>☐ 12. Scrubbed/cleaned surfaces such as door handles, faucets, phone, etc.<br>କୌଣସି ପୃଷ୍ଠକୁ ଘସିଛନ୍ତି/ସଫା କରିଛନ୍ତି ଯେପରିକି ଡୋର ହାଣ୍ଡେଲ, ଟ୍ୟାପ, ଫୋନ ଇତ୍ୟାଦି<br>☐ 13. Informed people of illness symptoms<br>ଲୋକ ମାନଙ୍କୁ ରୋଗର ଲକ୍ଷ୍ୟଣ ବିଷୟରେ ଜଣେଇଛନ୍ତି<br>☐ 14. Contacted a coronavirus helpline<br>କୌଣସି କରୋନାଭାଇରସ ହେଲ୍ପଲାଇନ କୁ ଯୋଗାଯୋଗ କରିଛନ୍ତି<br>☐ 15. Avoided hospitals/clinics<br>ହସ୍ପିଟାଲ କିମ୍ବା କ୍ଲିନିକକୁ ଏଡେଇଛନ୍ତି<br>☐ 16. Avoided public transit/traveling<br>ସର୍ବ ସାଧାରଣ ପରିବହନ/ଭ୍ରମଣକୁ ଏଡେଇଛନ୍ତି<br>☐ 17. Nothing କିଛି ନୁହେଁ<br>☐ 88. Other ଅନ୍ୟାନ୍ୟ: ________ |
| **B8.** What are some of the challenges you and your family face in being able to stay at home at all times except when you are performing essential activities such as working or buying food?<br><br>ଖାଦ୍ୟ କିଣିବା କିମ୍ବା କାମ କରିବା ଭଳି ଆବଶ୍ୟକୀୟ କାମ ଗୁଡିକୁ ଛାଡି ଅନ୍ୟ ସମୟ ଘରେ ରହିବାରେ, ଆପଣ ଓ ଆପଣଙ୍କ ପରିବାର ସଦସ୍ୟ କେଉଁ ଅସୁବିଧାର ସମ୍ମୁଖୀନ ହୁଅନ୍ତି? | |
| **B9.** What are some of the challenges you and your family face in being able to maintain 2 meters of distance between other non-family members when you are outside of your house?<br>ଘରଠାରୁ ବାହାରେ ସମସ୍ତ ସମୟ ଅଣ-ପରିବାର ସଦସ୍ୟ ମାନଙ୍କ ଠାରୁ ୨ ମିଟର ଦୂରତ୍ୱ ରକ୍ଷା କରିବା ସମୟରେ ଆପଣ ଓ ଆପଣଙ୍କ ପରିବାର ସଦସ୍ୟ କେଉଁ ଅସୁବିଧାର ସମ୍ମୁଖୀନ ହୁଅନ୍ତି? | |
| **B10.** What are some of the challenges you and your family face in being able to frequently wash your hands with water and soap? | |

### COVID-19 Mobile Phone Interview – Head of Household

| | |
|---|---|
| ସାବୁନ ଓ ପାଣିରେ ବାରମ୍ବାର ହାତ ଧୋଇବାକୁ ନେଇ ଆପଣ ଓ ଆପଣଙ୍କ ପରିବାର ସଦସ୍ୟ କେଉଁ ଅସୁବିଧାର ସମ୍ମୁଖୀନ ହୁଅନ୍ତି? | |
| **B11.** What types of people are at high risk for getting very seriously sick if they get coronavirus?<br><br>କେଉଁ ପ୍ରକାର ଲୋକଙ୍କୁ କରୋନାଭାଇରସ ହେଲେ ସେମାନେ ସର୍ବାଧିକ ଅସୁସ୍ଥ ହେବାର ବିପଦ ଅଛି?<br><br>Enumerator note: Do **NOT** read response options. Select all that apply.<br>ଏନୁମରେଟର ନୋଟ: ବିକଳ୍ପ ଉତ୍ତର ଗୁଡିକୁ ପଢନ୍ତୁ **ନାହିଁ**. ଦେଇଥିବା ସମସ୍ତ ଉତ୍ତରକୁ ବାଛନ୍ତୁ। | ☐ 1. Elderly/Over 60 ବୃଦ୍ଧ/60 ରୁ ଉର୍ଦ୍ଧ୍ୱ<br>☐ 2. Men ପୁରୁଷ<br>☐ 3. Women ମହିଳା<br>☐ 4. Babies (<1 year) ଛୁଆ (<୧ ବର୍ଷ)<br>☐ 5. Children ପିଲା<br>☐ 6. Pregnant women ଗର୍ଭବତୀ ମହିଳା<br>☐ 7. People who are already sick/weak immune systems<br>ଯେଉଁ ଲୋକମାନେ ପୂର୍ବରୁ ଅସୁସ୍ଥ/ ଯାହାର ରୋଗପ୍ରତିରୋଧକ ପ୍ରଣାଳୀ ଦୁର୍ବଳ ଅଟେ<br>☐ 8. Everyone<br>☐ 88. Other ଅନ୍ୟାନ୍ୟ: ________<br>☐ 99. Don't know ଜଣା ନାହିଁ |
| **Enumerator read**: I am now going to read a few questions where I ask you to think about a scenario where you or a family member have coronavirus or show symptoms. I want to emphasize to you that this is just an imagined scenario and only for survey purposes.<br><br>ମୁଁ ବର୍ତ୍ତମାନ କିଛି ପ୍ରଶ୍ନ ପଢିବାକୁ ଯାଉଛି ଯେବେ ମୁଁ ଆପଣଙ୍କୁ ଏକ ପରିସ୍ଥିତି ବିଷୟରେ ଭାବିବାକୁ କହିବି ଯେବେ ଆପଣଙ୍କୁ କିମ୍ବା ଆପଣଙ୍କ ପରିବାରର କେହି ସଦସ୍ୟଙ୍କୁ କରୋନାଭାଇରସ ହୋଇଛି କିମ୍ବା ଲକ୍ଷଣ ଦେଖା ଦେଉଛି। ମୁଁ ଏହା ଜୋର ଦେଇ କହୁଛି କି ଯେ ଏହା ଏକ କଳ୍ପିତ ପରିସ୍ଥିତି ହିଁ ଏବଂ କେବଳ ସର୍ଭେ କାରଣ ପାଇଁ। | |
| **B12.** For survey purposes, if you don't mind, can I please ask how concerned would you be if you or someone in your household got coronavirus? Why?<br><br>ସର୍ଭେର ଉଦ୍ଦେଶ୍ୟ ପାଇଁ, ଯଦି ଆପଣ ଖରାପ ନ ଭାବନ୍ତି, କଣ ମୁଁ ପଚାରିପାରେ ଯେ, ଆପଣ କିମ୍ବା ଆପଣଙ୍କ ଘରେ କାହାକୁ କରୋନାଭାଇରସ ହେଲେ ଆପଣ କେତେ ଚିନ୍ତିତ ହେବେ? | ☐ 1. Not concerned ଚିନ୍ତିତ ନୁହେଁ<br>☐ 2. Somewhat concerned କିଛିମାତ୍ରାରେ ଚିନ୍ତିତ<br>☐ 3. Very concerned ବହୁତ ଚିନ୍ତିତ<br>☐ 99. Don't know ଜଣା ନାହିଁ<br><br><u>**Explain why: ବର୍ଣ୍ଣନା କରନ୍ତୁ କାହିଁକି**</u> |
| **B13.** If you begin exhibiting symptoms, what actions will you take?<br>ଯଦି ଆପଣଙ୍କ ମଧ୍ୟରେ ଲକ୍ଷ୍ୟଣ ଦେଖାଯିବା ଆରମ୍ଭ ହେଇଯାଏ, ଆପଣ କଣ କରିବେ?<br><br>Enumerator Note: Do **NOT** read response options. Select all that apply.<br>ଏନୁମରେଟରଙ୍କ ପାଇଁ ନୋଟ: ବିକଳ୍ପ ଉତ୍ତର ଗୁଡିକୁ ପଢନ୍ତୁ ନାହିଁ. ଦେଇଥିବା ସମସ୍ତ ଉତ୍ତରକୁ ବାଛନ୍ତୁ | ☐ 1. Stay at home more ଘରେ ଅଧିକ ରହିବେ<br>☐ 2. Stop attending school or work<br>ସ୍କୁଲ କିମ୍ବା କାମକୁ ଯିବା ବନ୍ଦ କରିଦେବେ<br>☐ 3. Stop attending social gatherings (e.g., weddings, funerals, church, temple)<br>ସାମାଜିକ ସମାବେଶକୁ ଯିବା ବନ୍ଦ କରିଦେବେ (ଉଦାହରଣ : ବିବାହ,ଅନ୍ତିମ ସଂସ୍କାର, ଗୀର୍ଜାଘର, ମନ୍ଦିର)<br>☐ 4. Keep a distance of at least 1 meter from others<br>ଅନ୍ୟ ମାନଙ୍କ ଠାରୁ ଅତି କମରେ ୧ ମିଟର ଦୂରତ୍ୱ ରକ୍ଷା କରିବେ<br>☐ 5. Keep a distance of at least 2 meters from others |

ଅନ୍ୟ ମାନଙ୍କ ଠାରୁ ଅତି କମରେ ୨ ମିଟର ଦୂରତ୍ୱ ରକ୍ଷା କରିବେ

- ☐ 6. Wash hands more frequently ବାରମ୍ବାର ହାତ ଧୋଇବେ
- ☐ 7. Start wearing a mask ମାସ୍କ ପିନ୍ଧିବା ଆରମ୍ଭ କରିବେ
- ☐ 8. Go to health clinic ଚିକିତ୍ସାଳୟକୁ ଯିବେ
- ☐ 9. Go to be tested for coronavirus କରୋନାଭାଇରସର ପରୀକ୍ଷଣ ପାଇଁ ଯିବେ
- ☐ 10. Drink local alcohol ଦେଶୀ ମଦ ପିଇବେ
- ☐ 11. Inform people of illness symptoms
  ଲୋକ ମାନଙ୍କୁ ରୋଗର ଲକ୍ଷ୍ୟଣ ବିଷୟରେ ଜଣେଇବେ
- ☐ 12. Nothing କିଛି ନୁହେଁ
- ☐ 88. Other ଅନ୍ୟାନ୍ୟ: _________
- ☐ 99. Don't know ଜାଣି ନାହିଁ

**B14.** What has been your **main** source for information about coronavirus?

ଆପଣଙ୍କୁ କରୋନାଭାଇରସ ସହ ଜଡିତ ମିଳିଥିବା ସୂଚନାର **ମୁଖ୍ୟ** ଉତ୍ସ କଣ?

Enumerator notes:
For each source recorded, ask:
How much do you trust this source - do you not trust any information from this source, trust some of the information, or trust all of the information?

ଏନୁମରେଟର ନୋଟ : ପ୍ରତିଟି ଉତ୍ସ ପାଇଁ, ପଚାରନ୍ତୁ
ଆପଣ ଏହି ଉତ୍ସକୁ କେତେ ବିଶ୍ୱାସ କରନ୍ତି (ଏହି ଉତ୍ସର କୌଣସି ସୂଚନାକୁ ବିଶ୍ୱାସକରନ୍ତି ନାହିଁ, କିଛି ସୂଚନା ଉପରେ ବିଶ୍ୱାସକରନ୍ତି ନା ସବୁ ସୂଚନାକୁ ବିଶ୍ୱାସ କରନ୍ତି?)?

If the respondent is struggling to understand what "a source" is, you can read a few of the reponse options as examples.
ଯଦି ଉତ୍ତରଦାତାଙ୍କୁ 'ଉତ୍ସ' ଶବ୍ଦଟିକୁ ବୁଝିବାରେ ଅସୁବିଧା ହେଉଛି, ଆପଣ କିଛି ବିକଳ୍ପ ଉଦାହରଣ ସ୍ୱରୂପ ପଢିପାରିବେ।

If the respondent has more than one main source, you can record multiple sources.

ଯଦି ଉତ୍ତରଦାତାଙ୍କ ପାଖରେ ଗୋଟିଏରୁ ଅଧିକ ମୁଖ୍ୟ ଉତ୍ସ ଅଛି, ଆପଣ ଏକାଧିକ ଉତ୍ସକୁ ରେକର୍ଡ କରିପାରିବେ।

| **Source** ଉତ୍ସ | **Trust Level** ବିଶ୍ୱାସର ମାତ୍ରା | | |
|---|---|---|---|
| | Do not trust | Trust some | Trust all |
| ☐ 1. No sources mentioned | | | |
| ☐ 2. Government (National) | ☐ | ☐ | ☐ |
| ☐ 3. Government (State/Local) | ☐ | ☐ | ☐ |
| ☐ 4. Village Leader | ☐ | ☐ | ☐ |
| ☐ 5. NGO | ☐ | ☐ | ☐ |
| ☐ 6. Anganwadi | ☐ | ☐ | ☐ |
| ☐ 7. ASHA Worker | ☐ | ☐ | ☐ |
| ☐ 8. Other health worker | ☐ | ☐ | ☐ |
| ☐ 9. Neighbor in your village | ☐ | ☐ | ☐ |
| ☐ 10. Friend | ☐ | ☐ | ☐ |
| ☐ 11. Family | ☐ | ☐ | ☐ |
| ☐ 12. Social media/internet | ☐ | ☐ | ☐ |
| ☐ 13. News | ☐ | ☐ | ☐ |
| ☐ 14. Other: | ☐ | ☐ | ☐ |

**B15.** How have you gotten your information from this **main** source?

ଆପଣଙ୍କୁ ନିଜ **ମୁଖ୍ୟ** ଉତ୍ସର ସୂଚନା କେଉଁଠାରୁ ମିଳିଲା?

- ☐ 1. TV ଟିଭି
- ☐ 2. Radio ରେଡିଓ
- ☐ 3. Loudspeaker from vehicle ଗାଡିରେ ଲାଗିଥିବା ସ୍ପିକର
- ☐ 4. Newspaper ଖବରକାଗଜ
- ☐ 5. Phone call/SMS ଫୋନ କଲ/ଏସଏମଏସ

| | |
|---|---|
| | ☐ 6. In person conversation ବ୍ୟକ୍ତିଗତ କଥାବାର୍ତ୍ତା<br>☐ 7. Internet or phone app ଇଣ୍ଟରନେଟ କିମ୍ବା ଫୋନ ଆପ<br>☐ 88. Other ଅନ୍ୟାନ୍ୟ ________<br>☐ 99. Don't know ଜାଣିନାହିଁ |
| **B16.** From what other sources have you received information about coronavirus?<br>I am going to read a list of sources. For each one, please tell me yes or no.<br><br>Enumerator note: Read each response option.<br><br>କରୋନାଭାଇରସ ସହ ଜଡିତ ସୂଚନା ଆପଣଙ୍କୁ ଅନ୍ୟ କେଉଁ ଉତ୍ସରୁ ମିଳିଛି? ମୁଁ କିଛ ଉତ୍ସର ସୂଚୀ ପଢିଛି। ପ୍ରତିଟି ପାଇଁ, ଦୟାକରି ହଁ କିମ୍ବା ନାହିଁ କୁହନ୍ତୁ। | ☐ 1. Government (National) କେନ୍ଦ୍ର ସରକାର<br>☐ 2. Government (State/Local) ରାଜ୍ୟ/ସ୍ଥାନୀୟ ସରକାର<br>☐ 3. Village Leader ଗ୍ରାମର ନେତା<br>☐ 4. NGO ସ୍ୱେଚ୍ଛାସେବୀ ସଂଗଠନ<br>☐ 5. Anganwadi ଅଙ୍ଗନବାଡି<br>☐ 6. ASHA Worker ଆଶା କର୍ମୀ<br>☐ 7. Other health worker ଅନ୍ୟ ସ୍ୱାସ୍ଥ୍ୟ କର୍ମଚାରୀ<br>☐ 8 Neighbor in your village ଆପଣଙ୍କ ଗାଁ ର ପଡୋଶୀ<br>☐ 9. Friend ସାଙ୍ଗ<br>☐ 10. Family ପରିବାର<br>☐ 11. Social media/internet ସାମାଜିକ ଗଣମାଧ୍ୟମ/ଇଣ୍ଟେରନେଟ<br>☐ 12. News ନ୍ୟୁଜ<br>☐ 13. No other sources କିଛି ଉତ୍ସ ନାହିଁ<br>☐ 14. Other: |

#### SECTION C: COVID-19 Impact on Daily Life and Practices

**Enumerator read:** Thank you for your responses so far. Now I'm going to ask you some questions about how coronavirus has impacted your daily life.

ଏନୁମରେଟର ପଢନ୍ତୁ: ବର୍ତ୍ତମାନ ପର୍ଯ୍ୟନ୍ତ ଦେଇଥିବା ଆପଣଙ୍କ ଉତ୍ତର ପାଇଁ ଧନ୍ୟବାଦ। ମୁଁ ବର୍ତ୍ତମାନ କିଛି ପ୍ରଶ୍ନ କରୋନାଭାଇରସ ଆପଣଙ୍କ ଜୀବନକୁ କିପରି ପ୍ରଭାବିତ କରିଛି, ସେ ବିଷୟରେ ପଚାରିବି

| |
|---|
| **C1.** How has coronavirus or the lockdown changed your daily life? Think about from the time you get up in the morning to the time you go to sleep at night. How is your day different now, if it is different?<br><br>କରୋନାଭାଇରସ କିମ୍ବା ଲକଡାଉନ ଆପଣଙ୍କ ଦୈନିକ ଜୀବନକୁ କିପରି ବଦଳେଇ ଦେଇଛି? ସକାଳୁ ଉଠିବା ସମୟରୁ ଆରମ୍ଭ କରି ରାତିରେ ଶୋଇବାକୁ ଯିବା ପର୍ଯ୍ୟନ୍ତ ସମୟ ବିଷୟରେ ଭାବନ୍ତୁ। ବର୍ତ୍ତମାନ ଆପଣଙ୍କ ଦିନ ଅଲଗା କିପରି, ଯଦି ଅଲଗା ହୋଇଥାଏ?<br><br>***Probes:*** *ଅନୁସନ୍ଧାନ:*<br>○ Staying at home more? ଘରେ ଅଧିକ ସମୟ ରହୁଛନ୍ତି?<br>○ More/less workload? ଅଧିକ/କମ କାମ<br>○ More/less free time? ଅଧିକ/କମ ଖାଲି ସମୟ<br>○ Is there anywhere in the village you *used* to go but no longer go? ଗାଁରେ ଏପରି କିଛି ଜାଗା ଅଛି କି |

| | |
|---|---|
| ଯେଉଠିକି ଆପଣ ଯାଉଥିଲେ କିନ୍ତୁ ଆଉ ଯାଉ ନାହାନ୍ତି? | |
| **C2.** Has coronavirus or the lockdown changed how you or your family **interact with other members of your village**? Please explain.<br><br>କଣ କରୋନାଭାଇରସ କିମ୍ବା ଲକଡାଉନ ଆପଣଙ୍କର କିମ୍ବା ଆପଣଙ୍କ ପରିବାରର ଗ୍ରାମର ଅନ୍ୟ ଲୋକମାନଙ୍କ ସହିତ ମିଳାମିଶାରେ କିଛି ପରିବର୍ତ୍ତନ ଆଣିଛି? ଦୟାକରି ବର୍ଣ୍ଣନା କରନ୍ତୁ। | ☐ 1. Yes ହଁ<br>☐ 2. No ନାହିଁ<br><br>**Please explain. ଦୟାକରି ବର୍ଣ୍ଣନା କରନ୍ତୁ।** |
| **C3.** In the past 7 days, did you take any of the following actions to cover your household's basic needs?<br>ଗତ ସାତଦିନ ମଧ୍ୟରେ, ନିଜ ଘରର ସାଧାରଣ ଆବଶ୍ୟକତାକୁ ପୂରଣ କରିବା ପାଇଁ, ଆପଣ ଉକ୍ତ କେଉଁ କେଉଁ ପଦକ୍ଷେପ ନେଇଛନ୍ତି?<br><br>Enumerator: read all response options.<br>**ଏନୁମରେଟର ନୋଟ** : ସମସ୍ତ ବିକଳ୍ପ ଉତ୍ତରଗୁଡ଼ିକୁ ପଢନ୍ତୁ<br><br>Record any explanations the respondent gives | ☐ 1. Use cash savings or bank savings<br>ସଞ୍ଚିତ ଟଙ୍କା କିମ୍ବା ବ୍ୟାଙ୍କ ଜମାରାଶି ଖର୍ଚ୍ଚ କରିଛନ୍ତି<br>☐ 2. Borrow money or food from neighbor, friend, or relative ପଡୋଶୀ, ସାଙ୍ଗ କିମ୍ବା ସମ୍ପର୍କୀୟଙ୍କ ଠାରୁ ଟଙ୍କା କିମ୍ବା ଖାଦ୍ୟ ଉଧାର ଆଣିଛନ୍ତି<br>☐ 3. Sell assets ସମ୍ପତ୍ତି ବିକିଛନ୍ତି<br>☐ 4. Rely on Government assistance → **Ask follow-up question C3A**<br>ସରକାରୀ ସାହାଯ୍ୟ ଉପରେ ନିର୍ଭର କରିଛନ୍ତି<br>☐ 5. Rely on NGO assistance<br>ଏନଜିଓର ସାହାଯ୍ୟ ଉପରେ ନିର୍ଭର କରିଛନ୍ତି<br>☐ 6. None of the above ଏଥିମଧ୍ୟରୁ କିଛି ନୁହେଁ<br>☐ 7. Other ଅନ୍ୟାନ୍ୟ ________<br><br>**Record any explanations the respondent gives for this question: ଉତ୍ତରଦାତା ଦେଇଥିବା ଯେକୌଣସି ବର୍ଣ୍ଣନା କୁ ଟିପନ୍ତୁ :**<br><br>**If rely on government assistance is yes, ask:**<br>**ଯଦି ସରକାରୀ ସାହାଯ୍ୟ ଉପରେ ନିର୍ଭର କରୁଛନ୍ତି, ତାହେଲେ ପଚାରନ୍ତୁ :**<br><br>**C3A**. Which, if any, of the following government schemes have you received benefits from?<br>ଆପଣ ଉକ୍ତ କେଉଁ ସରକାରୀ ନୀତିରୁ ଲାଭବାନ ହେଇଛନ୍ତି?<br><br>Enumerator: read all response options.<br>**ଏନୁମରେଟର ଏନୁମରେଟର:** ସମସ୍ତ ବିକଳ୍ପ ଉତ୍ତରଗୁଡ଼ିକୁ ପଢନ୍ତୁ<br><br>☐ 1. National Social Assistance Programme for widows, senior citizens, differently abled<br>ଭାରତ ସରକାରଙ୍କର ବିଧବା, ବୃଦ୍ଧ ଓ ବିକଳାଙ୍ଗ ସାମାଜିକ ଭତ୍ତା କାର୍ଯ୍ୟକ୍ରମ<br>☐ 2. Free ration for BPL cardholders over three months<br>ବିପିଏଲ କାର୍ଡଧାରୀଙ୍କ ପାଇଁ 3 ମାସର ମାଗଣା ରାସନ<br>☐ 3. Jan Dhan account deposited with INR 500 for three months<br>ଜନଧନ ଖାତାରେ ତିନି ମାସ ପାଇଁ 500 ଟଙ୍କ ଜମା |

| | |
|---|---|
| | ☐ 4. Farmer accounts deposited with INR 2000<br>କୃଷକ ମାନଙ୍କ ବ୍ୟାଙ୍କ ଖାତାରେ 2000 ଟଙ୍କା ଜମା<br>☐ 5. Employment guarantee under National Rural Employment Guarantee Scheme<br>ଜାତୀୟ ଗ୍ରାମୀଣ କର୍ମନିଯୁକ୍ତି ଯୋଜନା (ନରେଗାଁ) ଦ୍ୱାରା ନିଶ୍ଚିତ ନିଯୁକ୍ତି<br>☐ 6. None କିଛି ନୁହେଁ |
| **C4.** Have you or any member of your family lost a job or work due to coronavirus or the lockdown?<br>କରୋନାଭାଇରସ ପାଇଁ, କଣ ଆପଣ କିମ୍ବା ଆପଣଙ୍କ ପରିବାରର କେହି ସଦସ୍ୟ ନିଜ ଚାକିରୀ କିମ୍ବା କାମ ହରେଇଛନ୍ତି କି?<br><br>If yes, who has lost their job and what was the occupation for the job that was lost?<br>ଯଦି ହଁ, କିଏ ନିଜ ଚାକିରୀ ହରେଇଛନ୍ତି ଓ ତାଙ୍କର ବୃତ୍ତି କଣ ଥିଲା? | ☐ 1. Yes ହଁ<br>☐ 2. No ନାହିଁ<br><br>**<u>If yes, record relationship and occupation</u>**:<br>ଯଦି ହଁ, ସମ୍ପର୍କ ଓ ବୃତ୍ତି ନୋଟ କରନ୍ତୁ: |
| **C5.** In the past 7 days, have you experienced any difficulties in getting the material you use to cook with (such as wood, gas, etc.) as a result of coronavirus or the lockdown?<br><br>ଗତ ସାତ ଦିନ ମଧ୍ୟରେ, କରୋନା ଭାଇରସ ପାଇଁ, ଆପଣଙ୍କୁ ରାନ୍ଧିବାରେ ଆବଶ୍ୟକ ହେଉଥିବା କିଛି ସାମଗ୍ରୀ (ଯେପରିକି କାଠ, ଗ୍ୟାସ ଇତ୍ୟାଦି) ମିଳିବାରେ କିଛି ଅସୁବିଧା ହୋଇଛି କି?<br><br>If yes, what type of material and please describe difficulty. ଯଦି ହଁ, କେଉଁ କି ପ୍ରକାର ବସ୍ତୁ ଓ ଦୟାକରି ବର୍ଣ୍ଣନା କରନ୍ତୁ କି ଅସୁବିଧା ହୋଇଛି ? | ☐ 1. Yes ହଁ<br>☐ 2. No ନାହିଁ<br><br>**<u>If yes, what type of material and please describe difficulty</u>:**<br>ଯଦି ହଁ, ଏହା କି ପ୍ରକାର ବସ୍ତୁ ଓ ଦୟାକରି ବର୍ଣ୍ଣନା କରନ୍ତୁ କି ଅସୁବିଧା ହୋଇଛି ? |
| **C6.** Has coronavirus or the lockdown changed the **types or quantity of food** your family eats daily? If yes, describe how this has changed.<br><br>କଣ କରୋନାଭାଇରସ କିମ୍ବା ଲକଡାଉନ ଆପଣଙ୍କ ପରିବାରର ର ଖାଇବାର ପରିମାଣ କିମ୍ବା ପ୍ରକାରରେ କିଛି ପରିବର୍ତ୍ତନ ଆଣିଛି? ଯଦି ହଁ, ବର୍ଣ୍ଣନା କରନ୍ତୁ କି ଏହା କି ପରିବର୍ତ୍ତନ ଆଣିଛି। | Change in **types of food**? ଖାଦ୍ୟର ପ୍ରକାରରେ ପରିବର୍ତ୍ତନ<br>☐ 1. Yes ହଁ<br>☐ 2. No ନାହିଁ<br><br>Change in **quantity of food**? ଖାଦ୍ୟର ପରିମାଣରେ ପରିବର୍ତ୍ତନ<br>☐ 1. Yes ହଁ – Increase<br>☐ 2. Yes ହଁ - Decrease<br>☐ 3. No ନାହିଁ<br><br>**<u>If yes, describe</u>**: ଯଦି ହଁ, ବର୍ଣ୍ଣନା କରନ୍ତୁ |
| **C7.** Are the number of people currently staying in your house more, less, or the same as the number of people that were staying in your house before coronavirus and the lockdown | ☐ 1. Less କମ<br>☐ 2. The same ସମାନ |

| | |
|---|---|
| started?<br><br>ଆପଣଙ୍କ ଘରେ ବର୍ତ୍ତମାନ ରହୁଥିବା ଲୋକଙ୍କ ସଂଖ୍ୟା କରୋନାଭାଇରସ ଏବଂ ଲକଡାଉନ ଆରମ୍ଭ ହେବ ପୂର୍ବରୁ ଘରେ ରହୁଥିବା ଲୋକଙ୍କ ସଂଖ୍ୟା ଠାରୁ ଅଧିକ, କମ ନା ସମାନ? | ☐ 3. More ଅଧିକ |
| **C8.** The last time you defecated, did you defecate in the open or use the latrine?<br><br>କଣ ଶେଷ ଥର ଯେବେ ଆପଣ ଝାଡା ଯାଇଥିଲେ ,ତେବେ ଆପଣ ଖୋଲା ରେ ଝାଡା ଯାଇଥିଲେ ନା ପାଇଖାନା ବ୍ୟବହାର କରିଥିଲେ**?** | ☐ 1. Open ଖୋଲା ରେ<br>☐ 2. Latrine ପାଇଖାନା ରେ<br>☐ 3. Somewhere else (not open field or latrine)<br>ଅଲଗା କେଉଁଠି (ଖୋଲା ପଡିଆ ନୁହେଁ କିମ୍ବା ପାଇଖାନା ନୁହେଁ)<br>☐ 99. Don't know ଜାଣିନାହିଁ |
| **C9.** Has coronavirus or the lockdown changed the **defecation location of you or any family members?** Please explain.<br><br>କଣ କରୋନାଭାଇରସ କିମ୍ବା ଲକଡ଼ାଉନ ଆପଣ କିମ୍ବା ଆପଣଙ୍କ କେହି ପରିବାର ସଦସ୍ୟଙ୍କ ଝାଡା ଯିବା ଜାଗାକୁ ବଦଳେଇଛି କି? ଦୟାକରି ବର୍ଣ୍ଣନା କରନ୍ତୁ<br><br>**Probe**: Have there been any changes for **elderly adults** in your household?<br>Have there been any changes for **young children** in your household?<br><br>ଆପଣଙ୍କ ଘରର **ବୃଦ୍ଧ ବ୍ୟକ୍ତିଙ୍କ** କ୍ଷେତ୍ରରେ କିଛି ପରିବର୍ତ୍ତନ ଆସିଛି କି**?**<br>ଆପଣଙ୍କ ଘରର **ଛୋଟ ପିଲାଙ୍କ** କ୍ଷେତ୍ରରେ କିଛି ପରିବର୍ତ୍ତନ ଆସିଛି କି**?** | ☐ 1. Yes ହଁ<br>☐ 2. No ନାହିଁ<br><br>**<u>Please explain. ବର୍ଣ୍ଣନା କରନ୍ତୁ</u>** |
| **C10.** Due to coronavirus or the lockdown, do you think people in this village are defecating in the open more, defecating in a latrine more, or there has been no change in where people defecate? Explain why.<br><br>ଆପଣ ଭାବୁଛନ୍ତି କି ଯେ, କରୋନାଭାଇରସ କିମ୍ବା ଲକଡ଼ାଉନ ପାଇଁ ଆପଣଙ୍କ ଗ୍ରାମର ଲୋକମାନେ, ଖୋଲାରେ ଅଧିକ ଝାଡା ଯାଉଛନ୍ତି, ଲାଟ୍ରିନର ରେ ଅଧିକ ଝାଡା ଯାଉଛନ୍ତି ନା ଲୋକ ଝାଡା କରୁଥିବା ଜାଗାରେ କୌଣସି ପରିବର୍ତ୍ତନ ଆସିନାହିଁ? ବର୍ଣ୍ଣନା କରନ୍ତୁ କଣ ପାଇଁ। | ☐ 1. More open defecation ଖୋଲରେ ଝାଡା ଯିବା ବଢିଛି<br>☐ 2. More latrine use ଲାଟ୍ରିନ ବ୍ୟବହାର ବଢିଛି<br>☐ 3. No change କିଛି ପରିବର୍ତ୍ତନ ନୁହେଁ<br>☐ 99. Don't know ଜାଣିନାହିଁ<br><br>**<u>Explain why: ବର୍ଣ୍ଣନା କରନ୍ତୁ କାହିଁକି</u>** |
| **C11.** Has coronavirus or the lockdown changed your **handwashing practices**? Please explain.<br><br>କଣ କରୋନାଭାଇରସ କିମ୍ବା ଲକଡ଼ାଉନ ଆପଣଙ୍କ **ହାତଧୁଆ ପ୍ରକ୍ରିୟାକୁ** ବଦଳେଇଛି? ଦୟାକରି ବର୍ଣ୍ଣନା କରନ୍ତୁ।<br><br>**Probes**: Has there been any change in **how often** you wash your hands? | ☐ 1. Yes ହଁ<br>☐ 2. No ନାହିଁ<br><br>**<u>Please explain. ଦୟାକରି ବର୍ଣ୍ଣନା କରନ୍ତୁ</u>** |

| | |
|---|---|
| କଣ ଆପଣ ପ୍ରାୟତଃ କେତେଥର ହାତ ଧୁଅନ୍ତି, ସେଥିରେ କିଛି ପରିବର୍ତ୍ତନ ଆସିଛି କି ?<br><br>Has there been any change in **how much soap or water you use** to wash your hands?<br><br>କଣ ଆପଣ ପ୍ରାୟତଃ କେତେ ପରିମାଣର ସାବୁନ କିମ୍ବା ପାଣି ବ୍ୟବହାର କରନ୍ତି, ସେଥିରେ କିଛି ପରିବର୍ତ୍ତନ ଆସିଛି କି? | |
| **C12.** In the past 7 days, have you had any difficulties with getting the water that you need for your household? Please explain.<br><br>କରୋନାଭାଇରସ କିମ୍ବା ଲକଡାଉନ ପାଇଁ ଗତ ସାତ ଦିନ ମଧ୍ୟରେ, କଣ ଆପଣଙ୍କୁ ନିଜ ଘରର ଆବଶ୍ୟକତା ପାଇଁ ଦରକାର ହେଉଥିବା ପାଣି ମିଳିବାରେ କିଛି ଅସୁବିଧା ହେଇଛି କି ? | ☐ 1. Yes ହଁ<br>☐ 2. No ନାହିଁ<br><br>**Please explain. ଦୟାକରି ବର୍ଣ୍ଣନା କରନ୍ତୁ** |
| **C13.** Has coronavirus or the lockdown changed your **drinking water practices** such as where you get your water, how you store it, and whether or not you clean/treat the water? Please explain.<br><br>କଣ କରୋନାଭାଇରସ କିମ୍ବା ଲକଡ଼ାଉନ ଆପଣଙ୍କ **ପାଣି ପିଇବା ସହିତ ଜଡିତ ପ୍ରକ୍ରିୟା ମାନଙ୍କୁ** ବଦଳେଇଛି କି, ଯେପରିକି ଆପଣ ପାଣି କେଉଁଠାରୁ ଆଣନ୍ତି, ତାକୁ କିପରି ରଖନ୍ତି କିମ୍ବା କିପରି ପରିଷ୍କାର କରନ୍ତି? ଦୟାକରି ବର୍ଣ୍ଣନା କରନ୍ତୁ। | Change in **where you get** your drinking water?<br>**ଆପଣ ପିଇବା ପାଣି ଆଣୁଥିବା ସ୍ଥାନରେ ପରିବର୍ତ୍ତନ**?<br>☐ 1. Yes ହଁ<br>☐ 2. No ନାହିଁ<br><br>Change in **how you store** your drinking water?<br>ଆପଣ କିପରି ପିଇବା ପାଣିକୁ ରଖନ୍ତି, ସେଥିରେ ପରିବର୍ତ୍ତନ?<br>☐ 1. Yes ହଁ<br>☐ 2. No ନାହିଁ<br><br>Change in whether or not you **clean/treat** your drinking water?<br>ଆପଣ ନିଜ ପିଇବା ପାଣିକୁ କିପରି ସଫା/ପରିଷ୍କାର କରନ୍ତି, ସେଥିରେ ପରିବର୍ତ୍ତନ?<br>☐ 1. Yes ହଁ<br>☐ 2. No ନାହିଁ<br><br>**Please explain. ଦୟାକରି ବର୍ଣ୍ଣନା କରନ୍ତୁ** |
| **C14.** Has coronavirus or the lockdown changed your **household cleaning practices**? Please explain.<br><br>କଣ କରୋନାଭାଇରସ କିମ୍ବା ଲକଡ଼ାଉନ ଆପଣଙ୍କ **ଘର ସଫା କରିବା ପ୍ରକ୍ରିୟାକୁ** ବଦଳେଇଛି? ଦୟାକରି ବର୍ଣ୍ଣନା କରନ୍ତୁ | ☐ 1. Yes ହଁ<br>☐ 2. No ନାହିଁ<br><br>**Please explain. ଦୟାକରି ବର୍ଣ୍ଣନା କରନ୍ତୁ** |

| | |
|---|---|
| **Probes**: Has there been any change in **how often** you clean your home?<br><br>କଣ ଆପଣ ପ୍ରାୟତଃ କେତେଥର ଘର ସଫା କରନ୍ତି, ସେଥିରେ କିଛି ପରିବର୍ତ୍ତନ ଆସିଛି କି ?<br><br>Has there been any change in **how much detergent/disinfectant or water you use** to clean your home?<br><br>କଣ ଆପଣ ଘର ସଫା କରିବା ପାଇଁ ପ୍ରାୟତଃ ଯେତେ ଲୁଗାଧୁଆ ପାଉଡର/ ଫିନାଇଲ କିମ୍ବା ପାଣି ବ୍ୟବହାର କରନ୍ତି, ସେଥିରେ କିଛି ପରିବର୍ତ୍ତନ ହେଇଛି କି? | |
| **C15.** Do you currently have soap or detergent in your household?<br><br>ଆପଣଙ୍କ ଗହରେ ବର୍ତ୍ତମାନ ସାବୁନ କିମ୍ବା ଲୁଗାଧୁଆ ପାଉଡର ଅଛି କି? | ☐ 1. Yes ହଁ<br>☐ 2. No ନାହିଁ → **Ask followup question C15A**<br>☐ 99. Don't know ଜାଣି ନାହିଁ<br><br><u>If answered no, ask:</u><br>C15A. The last time you needed to purchase soap for your household, did you experience any of the following?<br><br>ଶେଷଥର ପାଇଁ, ଯେବେ ଆପଣ ନିଜ ଘର ପାଇଁ ସାବୁନ କିଣିବାକୁ ଭାବିଲେ, କଣ ଆପଣ ଉକ୍ତ ଅନୁଭବ କରିଛନ୍ତି କି?<br><br>Enumerator: read all response options. Record any explanations the respondent gives.<br>**ଏନୁମରେଟର ନୋଟ** : ସମସ୍ତ ବିକଳ୍ପ ଉତ୍ତରଗୁଡିକୁ ପଢନ୍ତୁ । ଉତ୍ତରଦାତା ଦେଇଥିବା ଯେକୌଣସି ବର୍ଣ୍ଣନା କୁ ଟିପନ୍ତୁ<br><br>☐ 1. Difficulties in going to obtain **<u>soap</u>** due to mobility restrictions imposed by government<br>ସରକାରଙ୍କ ପ୍ରତିବନ୍ଧକ ମାନଙ୍କ ପାଇଁ ଦୋକାନକୁ ସାବୁନ କିଣିବାକୁ ଯିବାରେ ଅସୁବିଧା ହେଉଛି<br>☐ 2. Unable to buy the amount of **<u>soap</u>** we usually buy because the household income has dropped<br>ଘରର ଆୟ କମିଯିବା ଯୋଗୁଁ ଆମେ ସାଧାରଣତଃ ଯେତେ ସାବୁନ କିଣୁ, ତାହା କିଣିପାରୁନାହୁଁ<br>☐ 3. Unable to buy the amount of **<u>soap</u>** we usually buy because of limited supply<br>ସୀମିତ ଯୋଗାଣ ଯୋଗୁଁ ଆମେ ସାଧାରଣତଃ ଯେତେ ସାବୁନ କିଣୁ, ତାହା କିଣିପାରୁନାହୁଁ<br>☐ 88. Other ଅନ୍ୟାନ୍ୟ: ________<br><br>**<u>Record any explanations the respondent gives for this question:</u>**<br>ଉତ୍ତରଦାତା ଦେଇଥିବା ଯେକୌଣସି ବର୍ଣ୍ଣନା କୁ ଟିପନ୍ତୁ |

### COVID-19 Mobile Phone Interview – Head of Household

| | |
|---|---|
| **SECTION D: Household Information**<br><br>**Enumerator read:** Thank you for your responses about coronavirus. Now I'm going to ask you some questions about your household. This is the last section of the survey!<br><br>କରୋନାଭାଇରସ ଉପରେ ଆପଣଙ୍କ ଉତ୍ତର ପାଇଁ ଧନ୍ୟବାଦ। ମୁଁ ବର୍ତ୍ତମାନ ଆପଣଙ୍କୁ କିଛି ପ୍ରଶ୍ନ ନିଜ ଘର ବିଷୟରେ ପଚାରିବି। ଏହା ସର୍ଭେ ର ଶେଷ ଭାଗ। | |
| **D1.** What is the highest level of education that you have had?<br><br>ଆପଣ ସର୍ବୋତ୍ତମ କେତେ ପାଠ ପଢିଛନ୍ତି? | ☐ 1. No formal schooling ସ୍କୁଲ ଯାଇନାହାନ୍ତି<br>☐ 2. Some primary education କିଛି ପ୍ରାଥମିକ ପାଠ<br>☐ 3. Completed primary education ପ୍ରାଥମିକ ପାଠ ଶେଷ<br>☐ 4. Some secondary school କିଛି ଟା ସେକେଣ୍ଡରି ପାଠ<br>☐ 5. Matriculation (10th class completed) ମେଟ୍ରିକୁଲେଶନ<br>☐ 6. Completed secondary school କିଛି ଟା ସେକେଣ୍ଡରି ପାଠ ଶେଷ<br>☐ 7. Some college କିଛି କଲେଜ<br>☐ 8. Completed college/university କଲେଜ/ୟୁନିଭରସିଟି ଶେଷ<br>☐ 99. Don't know ଜାଣି ନାହିଁ |
| **D2.** What is your occupation?<br><br>ଆପଣଙ୍କ ବୃତ୍ତି କଣ ? | ☐ 1. Unemployed (no work that earns money)<br>ବେରୋଜଗାର(ରୋଜଗାର କରିଲା ଭଳିଆ କିଛି କାମ କରନ୍ତିନାହିଁ)<br>☐ 2. Self-Employed (Work in home: agriculture, potter, etc.)<br>ନିଜସ୍ୱ ରୋଜଗାର(ଯେମିତିକି- ଚାଷ, କୁମ୍ଭାର, ଅନ୍ୟାନ୍ୟ)<br>☐ 3. Employed (Work outside the home: office, labourer, factory, etc.)<br>ଚାକିରି(ଘରଠୁ ବାହାରେ କାମ କରନ୍ତି : ଉଦାହରଣ –ଅଫିସ, ଫ୍ୟାକ୍ଟ୍ରି, ଅନ୍ୟାନ୍ୟ)<br>☐ 4. Student ଛାତ୍ର |
| **D3.** What is the caste or tribe of the household?<br>ଆପଣଙ୍କର କେଉଁ - *ଜାତି ବା ଜନଜାତି* ?<br><br>Enumerator note: Read response options<br>ଏନୁମରେଟର ନୋଟ : ଉତ୍ତର ଅପସନସ୍ ଗୁଡିକୁ ପଢନ୍ତୁ । | ☐ 1. Brahmin ବ୍ରାହ୍ମଣ<br>☐ 2. General ସାଧାରଣ<br>☐ 3. Scheduled caste ଅନୁସୂଚିତ ଜାତି<br>☐ 4. Other backward caste ଅନ୍ୟ ପଛୁଆ ବର୍ଗ<br>☐ 5. Scheduled tribe ଅନୁସୂଚିତ ଜନଜାତି<br>☐ 6. Do not want to indicate caste ଜାତି କହିବାକୁ ଚାହୁଁନାହାଁନ୍ତି<br>☐ 77. Not applicable ଉପଯୁକ୍ତ ନୁହେଁ<br>☐ 88. Other ଅନ୍ୟାନ୍ୟ: ________<br>☐ 99. Don't know ଜାଣି ନାହିଁ |
| **D4.** What is the religion of the household?<br>ଆପଣଙ୍କର *କେଉଁ ଧର୍ମ*? | ☐ 1. Hindu ହିନ୍ଦୁ<br>☐ 2. Muslim ମୁଁସଲମାନ୍<br>☐ 3. Christian ଖ୍ରୀଷ୍ଟିଆନ୍<br>☐ 4. Sikh ଶିଖ୍ |

### COVID-19 Mobile Phone Interview – Head of Household

| | |
|---|---|
| | ☐ 5. Buddhist/Neo-buddhist ବୌଦ୍ଧ ଧର୍ମ<br>☐ 6. Jain ଜୈନ ଧର୍ମ<br>☐ 7. Jewish ଜୀଉ ଧର୍ମ<br>☐ 8. Parsi/Zorastrian ର୍ପାସୀ ଧର୍ମ<br>☐ 9. No Religion କୌଣସି ଧର୍ମ ନୁହେଁ<br>☐ 88. Other ଅନ୍ଯାନ୍ଯ: ________ |
| **D5.** Do you have an Antodaya Card and/or ration card? (select all that apply)<br>ଆପଣଙ୍କର ଅନ୍ତୋଦୟ /ରାସନ୍ କାର୍ଡ ଅଛି କି**?** | ☐ 1. Antodaya ଅନ୍ତୋଦୟ<br>☐ 2. Ration card ରାସନ କାର୍ଡ<br>☐ 3. None କିଛି ନାହିଁ<br>☐ 99. Don't know ଜାଣି ନାହିଁ |
| **END OF INTERVIEW:** Thank the respondent for their time and give them the COVID-19 information and resources summary provided by Gram Vikas.<br><br>ସାକ୍ଷାତକରର ଶେଷ: ଉତ୍ତରଦାତାଙ୍କୁ ତାଙ୍କ ସମୟ ପାଇଁ ଧନ୍ୟବାଦ ଜଣାନ୍ତୁ ଏବଂ ତାଙ୍କୁ ଗ୍ରାମବିକାଶ ଦ୍ୱାରା ଦିଆଯାଇଥିବା COVID-19 ସୂଚନା ବିଷୟରେ ଅବଗତ କରନ୍ତୁ | |
| **End Time**<br>ଶେଷ ସମୟ | |

### COVID-19 Mobile Phone Interview – VWSC

| **SECTION A: Respondent and Enumerator Information & Consent Process** | |
|---|---|
| **A1.** Village ID ଗ୍ରାମ ର କୋଡ: | **A2.** Household ID ଘର ଘରର ସଂଖ୍ୟା |
| **A3.** Village Name ଗ୍ରାମ ର ନାମ | |
| **A4.** Date ତାରିଖ | (y/d/m) |
| **A5.** Start Time ଆରମ୍ଭ ସମୟ | |
| **A6.** Enumerator name ଆପଣଙ୍କ ନାମ | ☐ 1. Alok<br>☐ 2. Dhiren<br>☐ 3. Varsha |
| **A7.** Confirm that the respondent and their family (if applicable) currently live in the intended village (and are not currently staying somewhere else).<br><br>ପୁଷ୍ଟି କରନ୍ତୁ କି ଯେ, ଉପଯୁକ୍ତ ଉତ୍ତରଦାତା ଏବଂ ତାଙ୍କ ପରିବାର (ଯଦି ଲାଗୁ ହୁଏ)ବର୍ତ୍ତମାନ ଉଦ୍ଦିଷ୍ଟ ଗ୍ରାମରେ ହିଁ ବସବାସ କରନ୍ତି (ଏବଂ ଅନ୍ୟ କେଉଁଠି ବସବାସ କରନ୍ତି ନାହିଁ) | ☐ 1. Yes ହଁ<br>☐ 2. No – respondent/family are staying outside the village and left *before* the pandemic → **END SURVEY**<br>ନା - ଉତ୍ତରଦାତା ଓ ତାଙ୍କ ପରିବାର ଗ୍ରାମ ବାହାରେ ବସବାସ କରନ୍ତି ଏବଂ ସର୍ବବ୍ୟାପୀ ମହାମାରୀ **ହେବା *ପୂର୍ବରୁ*** ଗ୍ରାମ ଛାଡି ଦେଇଛନ୍ତି - **ସର୍ଭେ ଶେଷ**<br>☐ 3. No – respondent/family are staying outside the village and left *because of* the pandemic → **END SURVEY**<br>ନା- ଉତ୍ତରଦାତା ଓ ତାଙ୍କ ପରିବାର ଗ୍ରାମ ବାହାରେ ବସବାସ କରନ୍ତି ଏବଂ ସର୍ବବ୍ୟାପୀ ମହାମାରୀ **ହେବା *କାରଣରୁ*** ଗ୍ରାମ ଛାଡି ଦେଇଛନ୍ତି - **ସର୍ଭେ ଶେଷ** |
| **A8a.** Read the consent form to the respondent and then ask for their consent to participate.<br><br>ଉତ୍ତରଦାତାଙ୍କ ପାଇଁ ସହମତି ପତ୍ରଟିକୁ ପଢନ୍ତୁ ଏବଂ ତାଙ୍କୁ **ସର୍ଭେରେ** ଅଂଶଗ୍ରହଣ ପାଇଁ ସହମତି ମାଗନ୍ତୁ।<br><br>Does the respondent consent to the survey?<br>କ'ଣ ଉତ୍ତରଦାତା ସର୍ଭେରେ ଅଂଶଗ୍ରହଣ ପାଇଁ ସହମତି ପ୍ରଦାନ କଲେ? | ☐ 1. Yes ହଁ<br>☐ 2. No → **END SURVEY** ନା - **ସର୍ଭେ ଶେଷ** |
| **A8b.** Does the respondent consent to having the phone call audio recorded?<br><br>କଣ ଉତ୍ତରଦାତା ଫୋନକଲର ରେକଡିଙ୍ଗ ପାଇଁ ସହମତି ପ୍ରଦାନ କଲେ? | ☐ 1. Yes ହଁ<br>☐ 2. No ନା → Pause when you need to in order to record detailed responses! ଯେତେବେଳେ ଉତ୍ତର ଲେଖିବା ଆବଶ୍ୟକ ପଡେ, ବିରାମ ନିଅନ୍ତୁ। |
| **ENUMERATOR NOTE:** Now you are going to begin the survey.<br>ଏନୁମରେଟରଙ୍କ ପାଇଁ ନୋଟ: ବର୍ତ୍ତମାନ ଆପଣ ସର୍ଭେ ଆରମ୍ଭ କରିବେ | |
| **A9.** What is your name?<br>ଆପଣଙ୍କ ନାମ କଣ ? | |

### COVID-19 Mobile Phone Interview – VWSC

| | |
|---|---|
| **A10.** What is the sex of the respondent?<br>ଉତ୍ତରଦାତଙ୍କ ଲିଙ୍ଗ କଣ?<br><br>Enumerator Note: Clarify with the respondent if you are not sure.<br>ଏନୁମରେଟର ନୋଟ - ସ୍ୱଷ୍ଟ କରନ୍ତୁ ଯେବେ ଆପଣ ନିଶ୍ଚିତ ନୁହନ୍ତି | ☐ 1. Male ପୁରୁଷ<br>☐ 2. Female ମହିଳା |
| **A11.** What is your age?<br>ଆପଣଙ୍କ ବୟସ କେତେ? | (years) (ବର୍ଷ) |
| **SECTION E: Additional questions for village stakeholders ONLY (e.g., VWSC members, ASHA or Anganwadi workers)** | |
| **E1.** Type of stakeholder | ☐ 1. VWSC president<br>☐ 2. VWSC member<br>☐ 3. ASHA worker<br>☐ 4. Anganwadi worker<br>☐ 5. Other: _______ |
| **E2.** Has the government or any NGOs provided any materials to your village related to coronavirus?<br><br>କଣ ସରକାର କିମ୍ବା କୌଣସି ଏନଜିଓ ଆପଣଙ୍କ ଗ୍ରାମକୁ କରୋନାଭାଇରସ ସମ୍ବନ୍ଧୀୟ କିଛି ଜିନିଷ ଯୋଗେଇଛନ୍ତି କି?<br><br>If yes, please describe.<br>ଯଦି ହଁ, ଦୟାକରି ବର୍ଣ୍ଣନା କରନ୍ତୁ | ☐ 1. Yes<br>☐ 2. No<br><br>**If yes, describe:**<br>**ଯଦି ହଁ, ବର୍ଣ୍ଣନା କରନ୍ତୁ** |
| **E4.** Has any guidance been provided to your village from the government or an NGO about what someone should do if they think they have been exposed to or might have coronavirus?<br><br>କଣ କେଉଁ ଏନଜିଓ କିମ୍ବା ସରକାର ଆପଣଙ୍କୁ ଏହି ନିର୍ଦ୍ଦେଶାବଳୀ ଦେଇଛନ୍ତି କି ଯେ ଯଦି କାହାକୁ ଏହା ଲାଗିଲା ଯେ ସେମାନଙ୍କୁ | ☐ 1. Yes – Government ହଁ-ସରକାର<br>☐ 2. Yes – NGO ହଁ-ଏନଜିଓ<br>☐ 3. Yes – Both – ହଁ- ଉଭୟ<br>☐ 4. No – ନାହିଁ |

| | |
|---|---|
| କାରୋନାଭାଇରସ ହେଇଛି, ତାହେଲେ କଣ କରିବା ଉଚିତ?<br><br>If yes, please describe.<br><br>ଯଦି ହଁ, ଦୟାକରି ବର୍ଣ୍ଣନା କରନ୍ତୁ | **<u>If yes, describe</u>:**<br><br>**<u>ଯଦି ହଁ, ବର୍ଣ୍ଣନା କରନ୍ତୁ</u>** |
| **E5.** What actions has this village taken as a community to protect itself against coronavirus? How were these actions decided upon?<br><br>ଏକ ଗୋଷ୍ଠୀ ହିସାବରେ ଏହି ଗ୍ରାମରେ କାରୋନା ଠାରୁ ରକ୍ଷା ପାଇବାକୁ କିଛି ପଦକ୍ଷେପ ଗ୍ରହଣ କରା ଯାଇଛି କି? ଏହି ପଦକ୍ଷେପ ଗୁଡିକ ବିଷୟରେ କିପରି ନିଷ୍ପତି ନିଆଗଲା? | |
| **E6.** Do you feel most villagers are taking precautions for preventing coronavirus (such as avoiding gatherings, washing their hands, etc.)? Why or why not?<br><br>ଆପଣଙ୍କୁ ଏହା ଲାଗୁଛି କି ଯେ, ସର୍ବାଧିକ ଗ୍ରାମବାସୀ କରୋନଭାଇରସରୁ ରକ୍ଷା ପାଇବା ସକାଶେ କାଉନସାଇ ସତର୍କତା ଅବଲମ୍ବନ କରୁଛନ୍ତି ଯେପରିକି ସାମାଜିକ ସମାବେଶରୁ ଦୂରେଇ ରହବା, ହାତ ଧୋଇବା ଇତ୍ୟାଦି? କାହିଁକି ଏବଂ କାହିଁକି ନୁହେଁ? | |
| **E7.** Is there any place in your village for migrants coming back to the village to quarantine/isolate temporarily upon arrival?<br><br>ଆପଣଙ୍କ ଗ୍ରାମରେ ଏପରି କିଛି ଜାଗା ଅଛିକି ଯେଉଠିକି ପ୍ରବାସୀ ଶ୍ରମିକ ମାନେ ଆସିବା ପରେ ନିଜକୁ ସଙ୍ଗରୋଧରେ ରଖିପାରିବେ କିମ୍ବା ଅସ୍ଥାୟୀ ଭାବରେ ନିଜକୁ ଅଲଗା କରି ରଖି ପାରିବେ?<br><br>If yes, please describe.<br><br>ଯଦି ହଁ, ଦୟାକରି ବର୍ଣ୍ଣନା କରନ୍ତୁ | ☐ 1. Yes- ହଁ<br>☐ 2. No ନାହିଁ<br><br>**<u>If yes, describe. If no, why not?</u>**<br><br>**<u>ଯଦି ହଁ, ବର୍ଣ୍ଣନା କରନ୍ତୁ</u>** |

| | |
|---|---|
| **E8.** Have any migrants returned to the village? If yes, describe how they were treated by other village members.<br><br>କଣ କେହି ପ୍ରବାସୀ ଶ୍ରମିକ ଆପଣଙ୍କ ଗାଁକୁ ଫେରିଛନ୍ତି? ଯଦି ହଁ, ବର୍ଣ୍ଣନା କରନ୍ତୁ ଯେ କିପରି ଗ୍ରାମର ସଦସ୍ୟ ମାନେ ସେମାନଙ୍କ ସହିତ କି ପ୍ରକାର ଆଚରଣ କଲେ? | ☐ 1. Yes ହଁ<br>☐ 2. No ନାହିଁ<br><br>**<u>If yes, describe:</u>**<br><br>**<u>ଯଦି ହଁ, ବର୍ଣ୍ଣନା କରନ୍ତୁ</u>** |
| **E9.** If someone in your village wanted to get tested for coronavirus, where could they go to get tested? Please describe.<br><br>ଯଦି ଆପଣଙ୍କ ଗ୍ରାମର କେହି କରୋନାଭାଇରସ ପାଇଁ ନିଜର ପରୀକ୍ଷଣ କରିବାକୁ ଚାହାନ୍ତି, ସେ କେଉଁଠିକୁ ଯାଇ ପରୀକ୍ଷଣ କରି ପାରିବେ? ଦୟାକରି ବର୍ଣନା କରନ୍ତୁ। | |
| **E10.** Has anyone in your village tested positive for coronavirus?<br><br>ଆପଣଙ୍କ ଗ୍ରାମରେ କେହି କରୋନଭାଇରସ ପଜିଟିଭ ହେଇଛି କି? | ☐ 1. Yes ହଁ<br>☐ 2. No ନାହିଁ<br>☐ 3. Don't know ଜାଣି ନାହିଁ |
| **E11.** How has daily life in your village changed as a result of coronavirus? Please describe.<br><br>ଆପଣଙ୍କ ଗ୍ରାମର ଦୈନନ୍ଦିନ ଜୀବନରେ କରୋନଭାଇରସ ଯୋଗୁଁ କି ପରିବର୍ତନ ଆସିଛି? | |
| **E12.** How have social interactions in the village changed as a result of coronavirus? Please describe.<br><br>ଆପଣଙ୍କ ଗ୍ରାମରେ ସାମାଜିକ ମିଳାମିଶାରେ କରୋନାଭାଇରସ | |

| | |
|---|---|
| ଯୋଗୁଁ କି ପରିବର୍ତନ ଆସିଛି? ଦୟାକରି ବର୍ଣ୍ଣନା କରନ୍ତୁ। | |
| **E13.** Since coronavirus or the lockdown, have you noticed an increase in any of the following village issues?<br><br>କରୋନାଭାଇରସ କିମ୍ବା ଲକଡାଉନ ଠାରୁ କଣ ଆପଣ ଉକ୍ତ ଗ୍ରାମୀଣ ସମସ୍ୟା ରେ କିଛି ବୃଦ୍ଧି ଦେଖିଛନ୍ତି କି?<br><br>If yes, please describe.<br><br>ଯଦି ହଁ, ଦୟାକରି ବର୍ଣ୍ଣନା କରନ୍ତୁ | ☐ 1. Domestic Violence ଘରୋଇ ହିଂସା<br>☐ 2. Alcoholism ମଦ୍ୟପାନ<br>☐ 3. Fighting ଝଗଡା<br>☐ 4. Theft ଚୋରି<br>☐ 5. None କିଛି ନୁହେଁ<br>☐ 88. Other ଅନ୍ୟାୟ: ________<br><br>**<u>If yes, describe:</u>**<br><br>**<u>ଯଦି ହଁ, ବର୍ଣ୍ଣନା କରନ୍ତୁ</u>** |
| **E14.** What changes, if any, have been made to the piped water supply service as a result of coronavirus?<br><br>କରୋନଭାଇରସ ପାଇଁ ପାଇପ ଦ୍ୱାରା ଜଳଯୋଗାଣରେ କିଛି ପରିବର୍ତନ କରାଯାଇଛି କି?<br><br>Enumerator note: read response options | ☐ 1. Extend the hours of piped water ଜଳଯୋଗାଣର ଘଣ୍ଟାକୁ ବଢେଇ ଦିଆ ଯାଇଛି<br>☐ 2. Reduce the hours of piped water ଜଳଯୋଗାଣର ଘଣ୍ଟାକୁ କମେଇ ଦିଆ ଯାଇଛି<br>☐ 3. Treatment of piped water<br>☐ 4. None<br>☐ 88. Other ଅନ୍ୟାୟ: ________<br><br>**<u>If any explanations given during response:</u>**<br><br>**<u>କଣ କିଛି ବିବରିଣୀ ଦିଆଯାଇଛି କି ?</u>** |
| **E15.** Are there any resources or needs of your village related to coronavirus that you do NOT think have been adequately addressed? | ☐ 1. Yes ହଁ |

| | |
|---|---|
| ଆପଣଙ୍କୁ ଏହା ଲାଗୁଛି କି ଯେ, ଆପଣଙ୍କ ଗ୍ରାମରେ କରୋନଭାଇରସ ସହ ଜଡିତ କିଛି ସଂସାଧନ କିମ୍ବା ଆବଶ୍ୟକତାର ଅଭାବ ଅଛି?<br>If yes, please describe.<br>ଯଦି ହଁ, ଦୟାକରି ବର୍ଣ୍ଣନା କରନ୍ତୁ | ☐ 2. No ନାହିଁ<br>**<u>If yes, describe</u>:**<br>**<u>ଯଦି ହଁ, ବର୍ଣ୍ଣନା କରନ୍ତୁ</u>** |
| **SECTION A - continued**<br><br>**Enumerator read:** Thank you for telling me about experiences in your village. Now I’m going to ask you some questions about your own experiences in your household. | |
| **A12.** Including yourself, how many people are currently staying in your house?<br>ଆପଣଙ୍କୁ ମିଶେଇ, ବର୍ତ୍ତମାନ ଆପଣଙ୍କ ଘରେ କେତେଜଣ ଲୋକ ରୁହନ୍ତି? | |
| **A13.** Are there any children <5 years currently living in your household?<br>କଣ ପାଞ୍ଚ ବର୍ଷରୁ କମ ବୟସର କେହି ପିଲା ବର୍ତ୍ତମାନ ଏହି ଘରେ ବସବାସ କରନ୍ତି କି? | ☐ 1. Yes ହଁ<br>☐ 2. No ନାହିଁ<br>**<u>If yes, children ages</u>:**<br>Child 1: Years: ________ Months: _________<br>Child 2: Years: ________ Months: _________<br>Child 3: Years: ________ Months: _________ |
| **A14.** How many people living in your household are over 60 years old?<br><br>ଆପଣଙ୍କ ଘରେ ରହୁଥିବା ସଦସ୍ୟ ମାନଙ୍କ ମଧ୍ୟରୁ କେତେ ଜଣଙ୍କ ବୟସ ୬୦ ବର୍ଷରୁ ଉର୍ଦ୍ଧ୍ୱ? | |
| **A15.** How many people living in your household are currently pregnant?<br><br>ଆପଣଙ୍କ ଘରେ ରହୁଥିବା ସଦସ୍ୟ ମାନଙ୍କ ମଧ୍ୟରୁ କେତେ ଜଣ ଗର୍ଭବତୀ? | |
| **A16.** Is anyone in your household a daily wage labourer who usually works in another state?<br><br>କଣ ଆପଣଙ୍କ ଘରର କେହି ଦିନ ମଜୁରିଆ ଅଟନ୍ତି କି ଯେ ସାଧାରଣତଃ ଅନ୍ୟ ରାଜ୍ୟରେ କାମ କରନ୍ତି ? | ☐ 1. Yes ହଁ - And they are still residing in another state ହଁ- ସେମାନେ ବର୍ତ୍ତମାନ ମଧ୍ୟ ଅନ୍ୟ ରାଜ୍ୟରେ ରହୁଛନ୍ତି<br>☐ 2. Yes ହଁ - But now they reside in Odisha ହଁ- କିନ୍ତୁ ସେମାନେ ବର୍ତ୍ତମାନ ଓଡିଶାରେ ରୁହନ୍ତି |

| | |
|---|---|
| | ☐ 3. No ନା |
| **A17.** Do you or any other family member have any of the following conditions or diseases?<br><br>ଆପଣଙ୍କୁ ଅଥବା ଆପଣଙ୍କ ପରିବାରର କେହି ସଦସ୍ୟଙ୍କୁ ଉକ୍ତ ରୋଗ ଅଛି କି?<br><br>Enumerator note: Read each option and select all that apply.<br>ଏନୁମରେଟରଙ୍କ ପାଇଁ ନୋଟ: ବିକଳ୍ପ ଉତ୍ତର ଗୁଡିକୁ **ପଢନ୍ତୁ।** ଦେଇଥିବା ସମସ୍ତ ଉତ୍ତରକୁ ବାଛନ୍ତୁ | ☐ 1. Asthma ଶ୍ୱାଶ<br>☐ 2. Diabetes ମଧୁମେହ<br>☐ 3. Obesity ମୋଟାପା<br>☐ 4. Kidney disease ବୃକକ ରୋଗ<br>☐ 5. Heart disease ହୃଦରୋଗ<br>☐ 6. Liver disease ଯକୃତ ରୋଗ<br>☐ 7. Chronic lung disease ଦୀର୍ଘକାଳୀନ ଫୁସଫୁସ ରୋଗ<br>☐ 8. Cancer କ୍ୟାନସର<br>☐ 9. None କିଛି ନାହିଁ |
| **A18.** Do you have piped water to your household or compound?<br><br>କଣ ଆପଣଙ୍କ ଘରକୁ ପାଇପ ଯୋଗେ ପାଣିର ବ୍ୟବସ୍ଥା ଅଛି? | ☐ 1. Yes – functional ହଁ- ସକ୍ରିୟ<br>☐ 2. Yes – not functional ହଁ- ନିଷ୍କ୍ରିୟ<br>☐ 3. No ନା<br><br>**<u>If 2 or 3:</u>**<br>What is your primary source of water and where is it located?<br><br><u>ଯଦି 2 କିମ୍ବା 3:</u><br>ଆପଣଙ୍କ ଘରର ମୁଖ୍ୟ ଜଳଉତ୍ସଟି କଣ ଓ ଏହା କେଉଁ ଠାରେ ଅବସ୍ଥିତ?<br><br>Source (ଉତ୍ସ):<br>☐ 1. Public tap ପବ୍ଲିକକଳ/ସର୍ବସାଧାରଣଟ୍ୟାପ୍<br>☐ 2. Well/borehole/spring କୂଅ, ନଳକୂଅ, ଝରଣା<br>☐ 3. Surface water (river/dam/lake/pond/stream/canal) ଭୂତଳଜଳ(ନଦୀ/ବନ୍ଧ/ହ୍ରଦ/ପୋଖରୀ/ନାଳ/କେନାଲ)<br>☐ 4. Rainwater ବର୍ଷାପାଣି<br>☐ 5. Bottled water ବୋତଲପାଣି<br>☐ 6. Other ଅନ୍ୟାନ୍ୟ<br>☐ 7. Don't know ଜାଣିନାହିଁ<br><br>Location: ଅବସ୍ଥାନ<br>☐ 1. In own dwelling ନିଜ ରହିବା ଜାଗାରେ<br>☐ 2. In own yard/plot/compound ନିଜଜାଗା/ପ୍ଲଟରେ/ଅଗଣାରେ<br>☐ 3. Outside the household compound ଘର ଅଗଣା/ପରିସର ବାହାରେ |
| **A19.** Does your household have a latrine?<br><br>କଣ ଆପଣଙ୍କ ଘରେ ପାଇଖାନା ଅଛି? | ☐ 1. Yes – functional ହଁ- ସକ୍ରିୟ<br>☐ 2. Yes – not functional ହଁ- ନିଷ୍କ୍ରିୟ<br>☐ 3. No ନାହିଁ |
| **A20.** Do you have an enclosed bathing area in | |

| | |
|---|---|
| your household compound?<br><br>କଣ ଆପଣଙ୍କ ଘର ପରିସର ମଧରେ ଏକ ଆବଦ୍ଧ ଗାଧୁଆଘର ଅଛି କି? | ☐ 1. Yes ହଁ<br>☐ 2. No ନା |
| **SECTION B: COVID-19 Knowledge, Perceptions, and Actions**<br><br>**Enumerator read:** Thank you for your responses so far. Now I'm going to ask you some questions related to coronavirus.<br><br>ଏନୁମରେଟର ପଢନ୍ତୁ : ବର୍ତ୍ତମାନ ପର୍ଯ୍ୟନ୍ତ ଦେଇଥିବା ଉତ୍ତର ମାନଙ୍କ ପାଇଁ ଧନ୍ୟବାଦ। ବର୍ତ୍ତମାନ, ମୁଁ ଆପଣଙ୍କୁ କରୋନାଭାଇରସ ସହିତ ଜଡିତ କିଛି ପ୍ରଶ୍ନ ପଚାରିବି। | |
| **B1.** Have you heard of a disease called covid-19, coronavirus, or corona?<br><br>କଣ ଆପଣ କୋଭିଡ-୧୯, କରୋନା-ଭାଇରସ କିମ୍ବା କରୋନା ନାମକ ରୋଗ ବିଷୟରେ ଶୁଣିଛନ୍ତି? | ☐ 1. Yes ହଁ<br>☐ 2. No → **If no, explain that this is the disease that has led to the recent lockdowns in India.**<br>ନା- ଯଦି ନା, ତାହେଲେ ଏହା କୁହନ୍ତୁ କି ଯେ ଏହି ରୋଗ ପାଇଁ ହିଁ ଏବେ ଭାରତରେ ଲକ-ଡାଉନ ହୋଇଛି |
| **B2.** What are the symptoms of coronavirus?<br><br>କରୋନା ଭାଇରସର ଲକ୍ଷଣ ଗୁଡିକ କଣ?<br><br><u>Enumerator notes:</u><br>Do **NOT** read response options. Select all that apply.<br><br>ସମ୍ଭାବ୍ୟ ଉତ୍ତର ଗୁଡିକୁ ପଢନ୍ତୁ ନାହିଁ. ଦେଇଥିବା ଉତ୍ତର ଗୁଡିକୁ ଟୀକ କରନ୍ତୁ.<br><br>Probe: Anything else?<br>ପ୍ରୋବ : ଆଉ କିଛି?<br><br>If respondent says sick/flu as their response, ask them to specify specific symptoms.<br><br>ଯଦି ଉତ୍ତରଦାତା ଅସୁସ୍ଥ ଲାଗିବା/ ଥଣ୍ଡା ହେବା ବିଷୟରେ କୁହନ୍ତି, ତାଙ୍କୁ ନିର୍ଦ୍ଦିଷ୍ଟ ଲକ୍ଷଣ ଗୁଡିକ ବିଷୟରେ କହିବାକୁ କୁହନ୍ତୁ | ☐ 1. No symptoms କିଛି ଲକ୍ଷଣ ନାହିଁ<br>☐ 2. Fever ଜ୍ୱର<br>☐ 3. Cough କାଶ<br>☐ 4. Dry cough ଶୁଖିଲା କାସ<br>☐ 5. Headache ମୁଣ୍ଡବଥା<br>☐ 6. Body aches ଶରୀର ବ୍ୟଥା<br>☐ 7. Difficulty breathing ନିଶ୍ୱାସ ନେବାରେ ଅସୁବିଧା<br>☐ 8. Chest pain ଛାତି ଯନ୍ତ୍ରଣା<br>☐ 9. Tiredness କ୍ଲାନ୍ତି<br>☐ 10. Diarrhea ଡ଼ାଇରିଆ<br>☐ 11. Chills ଥଣ୍ଡା ଲାଗିବା<br>☐ 12. Rash କୁଣ୍ଡିଆ<br>☐ 13. Loss of smell and/or taste ଗନ୍ଧ କିମ୍ବା ସ୍ୱାଦ ବାରିପାରୁନାହାନ୍ତି<br>☐ 14. Dizziness ମୁଣ୍ଡ ବୁଲେଇବା<br>☐ 15. Sneezing ଛିଙ୍କ/ଜିଙ୍କ<br>☐ 16. Sore throat ଖରାପ ଗଳା<br>☐ 88. Other ଅନ୍ୟାନ୍ୟ:<br>☐ 99. Don't know ଜାଣିନାହିଁ |
| **B3.** What causes a person to get sick with coronavirus?<br><br>କେଉଁ କାରଣ ମାନଙ୍କ ପାଇଁ ଜଣଙ୍କୁ କରୋନା ଭାଇରସ ହୋଇଥାଏ?<br><br>Enumerator note: Probe for respondent to describe in detail. If the respondent is having trouble answering, ask: "What actions would increase your risk of getting coronavirus? Why?" | |

| | |
|---|---|
| ଏନୁମରେଟରଙ୍କ ପାଇଁ ନୋଟ: ଉତ୍ତରଦାତାଙ୍କୁ ବର୍ଣ୍ଣନା କରିବା ପାଇଁ ଅନୁସନ୍ଧାନ କରନ୍ତୁ। ଯଦି ଉତ୍ତରଦାତାଙ୍କୁ ଉତ୍ତର ଦେବାରେ ଅସୁବିଧା ହେଉଥାଏ, ପଚାରନ୍ତୁ, "କେଉଁ କମ ଆପଣଙ୍କର କରୋନାଭାଇରସ ହେବାର ଆଶଙ୍କାକୁ ବଢେଇଦେଇଥାଏ? କାହିଁକି?" | |
| **B4.** Are there any cures or treatments for coronavirus disease?<br>କ'ଣ କରୋନା ଭାଇରସର କିଛି ଉପଶମ କିମ୍ବା ଚିକିତ୍ସା ଅଛି କି?<br><br>If yes, please describe these cures or treatments.<br>ଯଦି ହଁ, ଏହି ଉପଶମ କିମ୍ବା ଚିକିତ୍ସା ଗୁଡିକୁ ବର୍ଣ୍ଣନା କରନ୍ତୁ<br><br>Enumerator note: If the respondent describes their answer, please take notes on their response even if they respond 'no' or 'don't know'.<br><br>ଏନୁମରେଟର ନୋଟ : ଯଦି ଉତ୍ତରଦାତା ତାଙ୍କ ଉତ୍ତରକୁ ବର୍ଣ୍ଣନା କରନ୍ତି, ତାଙ୍କ ଉତ୍ତରକୁ ଟିପନ୍ତୁ ଯଦିଓ ସେମାନେ 'ନାହିଁ' କିମ୍ବା 'ଜଣା ନାହିଁ' ଉତ୍ତର ଦିଅନ୍ତି। | ☐ 1. Yes ହଁ<br>☐ 2. No ନାହିଁ<br>☐ 99. Don't know ଜଣାନାହିଁ<br><br>**Please describe:**<br>**ଦୟାକରି ବର୍ଣ୍ଣନା କରନ୍ତୁ** |
| **B5.** Do you think your personal chance of getting coronavirus is low, medium, high, or do you have no risk at all? Explain why.<br><br>ଆପଣଙ୍କୁ ଲାଗୁଛି କି ଯେ ଆପଣଙ୍କୁ କରୋନାଭାଇରସ ହେବାର ବ୍ୟକ୍ତିଗତ ସମ୍ଭାବନା କମ, ମଧ୍ୟମ, ଅଧିକ ଅଥବା ଆପଣଙ୍କ ପ୍ରତି କିଛି ବି ବିପଦ ନାହିଁ? ବର୍ଣ୍ଣନା କରନ୍ତୁ କାହିଁକି। | ☐ 1. Low କମ<br>☐ 2. Medium ମାଧ୍ୟମ<br>☐ 3. High ଅଧିକ<br>☐ 4. No risk କିଛି ବିପଦ ନୁହେଁ<br>☐ 99. Don't know ଜାଣିନାହିଁ<br><br>**Explain why: ବର୍ଣ୍ଣନା କରନ୍ତୁ କାହିଁକି** |
| **B6.** In the past 7 days, what actions have you and your family taken to avoid getting coronavirus, if any?<br><br>ଗତ ସାତଦିନ ମଧ୍ୟରେ, ଆପଣ ଓ ଆପଣଙ୍କ ପରିବାରର ସଦସ୍ୟ ମାନେ ନିଜକୁ କରୋନା ଭାଇରସରୁ ଦୂରେଇ ରଖିବାକୁ କଣ ପଦକ୍ଷେପ ନେଇଛନ୍ତି? | |
| **B7.** In the past 7 days, what actions have you and your family taken to avoid getting coronavirus, if any? I am going to read a list of actions. For each one, please tell me yes or no.<br><br>ଗତ ସାତ ଦିନ ମଧ୍ୟରେ, ଆପଣ ଓ ଆପଣଙ୍କ ପରିବାରର ସଦସ୍ୟ | ☐ 1. Stayed at home as much as possible<br>ଯେତେ ପାରିଛନ୍ତି, ଘରେ ରହିଛନ୍ତି<br>☐ 2. Did not attend school or work<br>ସ୍କୁଲ କିମ୍ବା କାମକୁ ଯାଇନାହାନ୍ତି<br>☐ 3. Did not attend social gatherings (e.g., weddings, funerals, church, temple) |

### COVID-19 Mobile Phone Interview – VWSC

| | |
|---|---|
| ମାନେ ନିଜକୁ କରୋନାଭାଇରସରୁ ଦୂରେଇ ରଖିବାକୁ କଣ ପଦକ୍ଷେପ ନେଇଛନ୍ତି? ମୁଁ କିଛି ପଦକ୍ଷେପ ପଢୁଛି। ପ୍ରତିଟି ପଦକ୍ଷେପ ପାଇଁ ହଁ କିମ୍ବା ନାହିଁ କୁହନ୍ତୁ।<br><br>Enumerator note: Select each action that the respondent says 'yes' for.<br><br>ଏନୁମରେଟରଙ୍କ ପାଇଁ ନୋଟ: ସବୁ ପଦକ୍ଷେପ ବାଛନ୍ତୁ, ଯାହାପାଇଁ ଉତ୍ତରଦାତା 'ହଁ' କହିଛନ୍ତି । | ସାମାଜିକ ସମାବେଶକୁ ଯାଇନାହାନ୍ତି (ଉଦାହରଣ : ବିବାହ,ଅନ୍ତିମ ସଂସ୍କାର, ଗୀର୍ଜାଘର, ମନ୍ଦିର)<br>☐ 4. Kept a distance from others → What distance? ____ meters<br>ଅନ୍ୟ ମାନଙ୍କ ଠାରୁ ଦୂରତ୍ୱ ରକ୍ଷା କରିଛନ୍ତି – କେତେ ମିଟର ଦୁରତ୍ୱ<br>☐ 5. Washed hands/used hand sanitizer more frequently<br>ହାତ ଧୋଇବା / ସାନିଟାଇଜର ର ବ୍ୟବହାର ବଢ଼େଇ ଦେଇଛନ୍ତି<br>☐ 6. Wore a (face) mask<br>ମାସ୍କ ପିନ୍ଧିଛନ୍ତି<br>☐ 7. Wore gloves (on your hands)<br>ଗ୍ଲୋବ୍ସ ପିନ୍ଧିଛନ୍ତି<br>☐ 8. Tried to stop touching face<br>ମୁହଁଟିକୁ ନ ଛୁଇଁବାର ଚେଷ୍ଟା କରିଛନ୍ତି<br>☐ 9. Did not shake hands with others<br>ଅନ୍ୟ ମାନଙ୍କ ସହିତ ହାତ ମିଶେଇନାହାନ୍ତି<br>☐ 10. Covered your mouth with elbow when you sneezed or coughed<br>ଛିଙ୍କିବା କିମ୍ବା କାଶିବା ସମୟରେ ପାଟିକୁ କହୁଣୀରେ ଘୋଡାଇ ଦେଇଛନ୍ତି<br>☐ 11. Drank local alcohol ଦେଶୀ ମଦ ପିଇଛନ୍ତି<br>☐ 12. Scrubbed/cleaned surfaces such as door handles, faucets, phone, etc.<br>କୌଣସି ପୃଷ୍ଠକୁ ଘସିଛନ୍ତି/ସଫା କରିଛନ୍ତି ଯେପରିକି ଡୋର ହାଣ୍ଡେଲ, ଟ୍ୟାପ, ଫୋନ ଇତ୍ୟାଦି<br>☐ 13. Informed people of illness symptoms<br>ଲୋକ ମାନଙ୍କୁ ରୋଗର ଲକ୍ଷ୍ୟଣ ବିଷୟରେ ଜଣେଇଛନ୍ତି<br>☐ 14. Contacted a coronavirus helpline<br>କୌଣସି କରୋନାଭାଇରସ ହେଲ୍ପଲାଇନ କୁ ଯୋଗାଯୋଗ କରିଛନ୍ତି<br>☐ 15. Avoided hospitals/clinics<br>ହସ୍ପିଟାଲ କିମ୍ବା କ୍ଲିନିକକୁ ଏଡେଇଛନ୍ତି<br>☐ 16. Avoided public transit/traveling<br>ସର୍ବ ସାଧାରଣ ପରିବହନ/ଭ୍ରମଣକୁ ଏଡେଇଛନ୍ତି<br>☐ 17. Nothing କିଛି ନୁହେଁ<br>☐ 88. Other ଅନ୍ୟାନ୍ୟ: ________ |
| **B8.** What are some of the challenges you and your family face in being able to stay at home at all times except when you are performing essential activities such as working or buying food?<br><br>ଖାଦ୍ୟ କିଣିବା କିମ୍ବା କାମ କରିବା ଭଳି ଆବଶ୍ୟକୀୟ କାମ ଗୁଡିକୁ ଛାଡି ଅନ୍ୟ ସମୟ ଘରେ ରହିବାରେ, ଆପଣ ଓ ଆପଣଙ୍କ ପରିବାର ସଦସ୍ୟ କେଉଁ ଅସୁବିଧାର ସମ୍ମୁଖୀନ ହୁଅନ୍ତି? | |
| **B9.** What are some of the challenges you and your family face in being able to maintain 2 meters of distance between other non-family members when you are outside of your house?<br>ଘରଠାରୁ ବାହାରେ ସମସ୍ତ ସମୟ ଅଣ-ପରିବାର ସଦସ୍ୟ ମାନଙ୍କ ଠାରୁ ୨ ମିଟର ଦୂରତ୍ୱ ରକ୍ଷା କରିବା ସମୟରେ ଆପଣ ଓ ଆପଣଙ୍କ ପରିବାର ସଦସ୍ୟ କେଉଁ ଅସୁବିଧାର ସମ୍ମୁଖୀନ ହୁଅନ୍ତି? | |

| | |
|---|---|
| **B10.** What are some of the challenges you and your family face in being able to frequently wash your hands with water and soap?<br><br>ସାବୁନ ଓ ପାଣିରେ ବାରମ୍ବାର ହାତ ଧୋଇବାକୁ ନେଇ ଆପଣ ଓ ଆପଣଙ୍କ ପରିବାର ସଦସ୍ୟ କେଉଁ ଅସୁବିଧାର ସମ୍ମୁଖୀନ ହୁଅନ୍ତି? | |
| **B11.** What types of people are at high risk for getting very seriously sick if they get coronavirus?<br><br>କେଉଁ ପ୍ରକାର ଲୋକଙ୍କୁ କରୋନାଭାଇରସ ହେଲେ ସେମାନେ ସର୍ବାଧିକ ଅସୁସ୍ଥ ହେବାର ବିପଦ ଅଛି?<br><br>Enumerator note: Do **NOT** read response options. Select all that apply.<br>ଏନୁମରେଟର ନୋଟ: ବିକଳ୍ପ ଉତ୍ତର ଗୁଡିକୁ ପଢନ୍ତୁ **ନାହିଁ**. ଦେଇଥିବା ସମସ୍ତ ଉତ୍ତରକୁ ବାଛନ୍ତୁ। | ☐ 1. Elderly/Over 60 ବୃଦ୍ଧ/60 ରୁ ଊର୍ଦ୍ଧ୍ୱ<br>☐ 2. Men ପୁରୁଷ<br>☐ 3. Women ମହିଳା<br>☐ 4. Babies (<1 year) ଛୁଆ (<୧ ବର୍ଷ)<br>☐ 5. Children ପିଲା<br>☐ 6. Pregnant women ଗର୍ଭବତୀ ମହିଳା<br>☐ 7. People who are already sick/weak immune systems<br>ଯେଉଁ ଲୋକମାନେ ପୂର୍ବରୁ ଅସୁସ୍ଥ/ ଯାହାର ରୋଗପ୍ରତିରୋଧକ ପ୍ରଣାଳୀ ଦୁର୍ବଳ ଅଟେ<br>☐ 8. Everyone<br>☐ 88. Other ଅନ୍ୟାନ୍ୟ: ________<br>☐ 99. Don't know ଜଣା ନାହିଁ |
| **Enumerator read**: I am now going to read a few questions where I ask you to think about a scenario where you or a family member have coronavirus or show symptoms. I want to emphasize to you that this is just an imagined scenario and only for survey purposes.<br><br>ମୁଁ ବର୍ତ୍ତମାନ କିଛି ପ୍ରଶ୍ନ ପଢିବାକୁ ଯାଉଛି ଯେବେ ମୁଁ ଆପଣଙ୍କୁ ଏକ ପରିସ୍ଥିତି ବିଷୟରେ ଭାବିବାକୁ କହିବି ଯେବେ ଆପଣଙ୍କୁ କିମ୍ବା ଆପଣଙ୍କ ପରିବାରର କେହି ସଦସ୍ୟଙ୍କୁ କରୋନାଭାଇରସ ହୋଇଛି କିମ୍ବା ଲକ୍ଷଣ ଦେଖା ଦେଉଛି। ମୁଁ ଏହା ଜୋର ଦେଇ କହୁଛି କି ଯେ ଏହା ଏକ କଳ୍ପିତ ପରିସ୍ଥିତି ହିଁ ଏବଂ କେବଳ ସର୍ଭେ କାରଣ ପାଇଁ। | |
| **B12.** For survey purposes, if you don't mind, can I please ask how concerned would you be if you or someone in your household got coronavirus? Why?<br><br>ସର୍ଭେର ଉଦ୍ଦେଶ୍ୟ ପାଇଁ, ଯଦି ଆପଣ ଖରାପ ନ ଭାବନ୍ତି, କଣ ମୁଁ ପଚାରିପାରେ ଯେ, ଆପଣ କିମ୍ବା ଆପଣଙ୍କ ଘରେ କାହାକୁ କରୋନାଭାଇରସ ହେଲେ ଆପଣ କେତେ ଚିନ୍ତିତ ହେବେ? | ☐ 1. Not concerned ଚିନ୍ତିତ ନୁହେଁ<br>☐ 2. Somewhat concerned କିଛିମାତ୍ରାରେ ଚିନ୍ତିତ<br>☐ 3. Very concerned ବହୁତ ଚିନ୍ତିତ<br>☐ 99. Don't know ଜଣା ନାହିଁ<br><br>**Explain why: ବର୍ଣ୍ଣନା କରନ୍ତୁ କାହିଁକି** |
| **B13.** If you begin exhibiting symptoms, what actions will you take?<br>ଯଦି ଆପଣଙ୍କ ମଧ୍ୟରେ ଲକ୍ଷ୍ୟଣ ଦେଖାଯିବା ଆରମ୍ଭ ହେଇଯାଏ, ଆପଣ କଣ କରିବେ?<br><br>Enumerator Note: Do **NOT** read response options. Select all that apply. | ☐ 1. Stay at home more ଘରେ ଅଧିକ ରହିବେ<br>☐ 2. Stop attending school or work<br>ସ୍କୁଲ କିମ୍ବା କାମକୁ ଯିବା ବନ୍ଦ କରିଦେବେ<br>☐ 3. Stop attending social gatherings (e.g., weddings, funerals, church, temple)<br>ସାମାଜିକ ସମାବେଶକୁ ଯିବା ବନ୍ଦ କରିଦେବେ (ଉଦାହରଣ : ବିବାହ,ଅନ୍ତିମ ସଂସ୍କାର, ଗୀର୍ଜାଘର, |

ଏନୁମରେଟରଙ୍କ ପାଇଁ ନୋଟ: ବିକଳ୍ପ ଉତ୍ତର ଗୁଡିକୁ ପଢନ୍ତୁ ନାହିଁ. ଦେଇଥିବା ସମସ୍ତ ଉତ୍ତରକୁ ବାଛନ୍ତୁ

ମନ୍ଦିର)

- ☐ 4. Keep a distance of at least 1 meter from others
  ଅନ୍ୟ ମାନଙ୍କ ଠାରୁ ଅତି କମରେ ୧ ମିଟର ଦୂରତ୍ୱ ରକ୍ଷା କରିବେ
- ☐ 5. Keep a distance of at least 2 meters from others
  ଅନ୍ୟ ମାନଙ୍କ ଠାରୁ ଅତି କମରେ ୨ ମିଟର ଦୂରତ୍ୱ ରକ୍ଷା କରିବେ
- ☐ 6. Wash hands more frequently ବାରମ୍ବାର ହାତ ଧୋଇବେ
- ☐ 7. Start wearing a mask ମାସ୍କ ପିନ୍ଧିବା ଆରମ୍ଭ କରିବେ
- ☐ 8. Go to health clinic ଚିକିତ୍ସାଳୟକୁ ଯିବେ
- ☐ 9. Go to be tested for coronavirus କରୋନାଭାଇରସର ପରୀକ୍ଷଣ ପାଇଁ ଯିବେ
- ☐ 10. Drink local alcohol ଦେଶୀ ମଦ ପିଇବେ
- ☐ 11. Inform people of illness symptoms
  ଲୋକ ମାନଙ୍କୁ ରୋଗର ଲକ୍ଷ୍ୟଣ ବିଷୟରେ ଜଣେଇବେ
- ☐ 12. Nothing କିଛି ନୁହେଁ
- ☐ 88. Other ଅନ୍ୟାନ୍ୟ: ________
- ☐ 99. Don’t know ଜାଣି ନାହିଁ

**B14.** What has been your **main** source for information about coronavirus?

ଆପଣଙ୍କୁ କରୋନାଭାଇରସ ସହ ଜଡିତ ମିଳିଥିବା ସୂଚନାର **ମୁଖ୍ୟ** ଉତ୍ସ କଣ?

Enumerator notes:
For each source recorded, ask:
How much do you trust this source - do you not trust any information from this source, trust some of the information, or trust all of the information?

ଏନୁମରେଟର ନୋଟ : ପ୍ରତିଟି ଉତ୍ସ ପାଇଁ, ପଚାରନ୍ତୁ
ଆପଣ ଏହି ଉତ୍ସକୁ କେତେ ବିଶ୍ୱାସ କରନ୍ତି (ଏହି ଉତ୍ସର କୌଣସି ସୂଚନାକୁ ବିଶ୍ୱାସକରନ୍ତି ନାହିଁ, କିଛି ସୂଚନା ଉପରେ ବିଶ୍ୱାସକରନ୍ତି ନା ସବୁ ସୂଚନାକୁ ବିଶ୍ୱାସ କରନ୍ତି?)?

If the respondent is struggling to understand what “a source” is, you can read a few of the reponse options as examples.
ଯଦି ଉତ୍ତରଦାତାଙ୍କୁ 'ଉତ୍ସ' ଶବ୍ଦଟିକୁ ବୁଝିବାରେ ଅସୁବିଧା ହେଉଛି, ଆପଣ କିଛି ବିକଳ୍ପ ଉଦାହରଣ ସ୍ୱରୂପ ପଢିପାରିବେ।

If the respondent has more than one main source, you can record multiple sources.

ଯଦି ଉତ୍ତରଦାତାଙ୍କ ପାଖରେ ଗୋଟିଏରୁ ଅଧିକ ମୁଖ୍ୟ ଉତ୍ସ ଅଛି, ଆପଣ ଏକାଧିକ ଉତ୍ସକୁ ରେକର୍ଡ କରିପାରିବେ।

| **Source** ଉତ୍ସ | **Trust Level** ବିଶ୍ୱାସର ମାତ୍ରା | | |
|---|---|---|---|
| | Do not trust | Trust some | Trust all |
| ☐ 1. No sources mentioned | | | |
| ☐ 2. Government (National) | ☐ | ☐ | ☐ |
| ☐ 3. Government (State/Local) | ☐ | ☐ | ☐ |
| ☐ 4. Village Leader | ☐ | ☐ | ☐ |
| ☐ 5. NGO | ☐ | ☐ | ☐ |
| ☐ 6. Anganwadi | ☐ | ☐ | ☐ |
| ☐ 7. ASHA Worker | ☐ | ☐ | ☐ |
| ☐ 8. Other health worker | ☐ | ☐ | ☐ |
| ☐ 9. Neighbor in your village | ☐ | ☐ | ☐ |
| ☐ 10. Friend | ☐ | ☐ | ☐ |
| ☐ 11. Family | ☐ | ☐ | ☐ |
| ☐ 12. Social media/internet | ☐ | ☐ | ☐ |
| ☐ 13. News | ☐ | ☐ | ☐ |
| ☐ 14. Other: | ☐ | ☐ | ☐ |

**B15.** How have you gotten your information from this **main** source?

- ☐ 1. TV ଟିଭି

### COVID-19 Mobile Phone Interview – VWSC

| | |
|---|---|
| ଆପଣଙ୍କୁ ନିଜ **ମୁଖ୍ୟ** ଉତ୍ସର ସୂଚନା କେଉଁଠାରୁ ମିଳିଲା? | ☐ 2. Radio ରେଡିଓ<br>☐ 3. Loudspeaker from vehicle ଗାଡିରେ ଲାଗିଥିବା ସ୍ପିକର<br>☐ 4. Newspaper ଖବରକାଗଜ<br>☐ 5. Phone call/SMS ଫୋନ କଲ/ଏସଏମଏସ<br>☐ 6. In person conversation ବ୍ୟକ୍ତିଗତ କଥାବାର୍ତ୍ତା<br>☐ 7. Internet or phone app ଇଣ୍ଟରନେଟ କିମ୍ବା ଫୋନ ଆପ<br>☐ 88. Other ଅନ୍ୟାନ୍ୟ ________<br>☐ 99. Don't know ଜାଣିନାହିଁ |
| **B16.** From what other sources have you received information about coronavirus?<br>I am going to read a list of sources. For each one, please tell me yes or no.<br><br>Enumerator note: Read each response option.<br><br>କରୋନାଭାଇରସ ସହ ଜଡିତ ସୂଚନା ଆପଣଙ୍କୁ ଅନ୍ୟ କେଉଁ ଉତ୍ସରୁ ମିଳିଛି**?** ମୁଁ କିଛ ଉତ୍ସର ସୂଚୀ ପଢିଛି। ପ୍ରତିଟି ପାଇଁ**,** ଦୟାକରି ହଁ କିମ୍ବା ନାହିଁ କୁହନ୍ତୁ। | ☐ 1. Government (National) କେନ୍ଦ୍ର ସରକାର<br>☐ 2. Government (State/Local) ରାଜ୍ୟ/ସ୍ଥାନୀୟ ସରକାର<br>☐ 3. Village Leader ଗ୍ରାମର ନେତା<br>☐ 4. NGO ସ୍ୱେଚ୍ଛାସେବୀ ସଂଗଠନ<br>☐ 5. Anganwadi ଅଙ୍ଗନବାଡି<br>☐ 6. ASHA Worker ଆଶା କର୍ମୀ<br>☐ 7. Other health worker ଅନ୍ୟ ସ୍ୱାସ୍ଥ୍ୟ କର୍ମଚାରୀ<br>☐ 8 Neighbor in your village ଆପଣଙ୍କ ଗାଁ ର ପଡୋଶୀ<br>☐ 9. Friend ସାଙ୍ଗ<br>☐ 10. Family ପରିବାର<br>☐ 11. Social media/internet ସାମାଜିକ ଗଣମାଧ୍ୟମ/ଇଣ୍ଟେରନେଟ<br>☐ 12. News ନ୍ୟୁଜ<br>☐ 13. No other sources କିଛି ଉତ୍ସ ନାହିଁ<br>☐ 14. Other: |

#### SECTION C: COVID-19 Impact on Daily Life and Practices

**Enumerator read:** Thank you for your responses so far. Now I'm going to ask you some questions about how coronavirus has impacted your daily life.

ଏନୁମରେଟର ପଢନ୍ତୁ: ବର୍ତ୍ତମାନ ପର୍ଯ୍ୟନ୍ତ ଦେଇଥିବା ଆପଣଙ୍କ ଉତ୍ତର ପାଇଁ ଧନ୍ୟବାଦ। ମୁଁ ବର୍ତ୍ତମାନ କିଛି ପ୍ରଶ୍ନ କରୋନାଭାଇରସ ଆପଣଙ୍କ ଜୀବନକୁ କିପରି ପ୍ରଭାବିତ କରିଛି, ସେ ବିଷୟରେ ପଚାରିବି

| |
|---|
| **C1.** How has coronavirus or the lockdown changed your daily life? Think about from the time you get up in the morning to the time you go to sleep at night. How is your day different now, if it is different?<br><br>କରୋନାଭାଇରସ କିମ୍ବା ଲକଡାଉନ ଆପଣଙ୍କ ଦୈନିକ ଜୀବନକୁ କିପରି ବଦଳେଇ ଦେଇଛି? ସକାଳୁ ଉଠିବା ସମୟରୁ ଆରମ୍ଭ କରି ରାତିରେ ଶୋଇବାକୁ ଯିବା ପର୍ଯ୍ୟନ୍ତ ସମୟ ବିଷୟରେ ଭାବନ୍ତୁ। ବର୍ତ୍ତମାନ ଆପଣଙ୍କ ଦିନ ଅଲଗା କିପରି, ଯଦି ଅଲଗା ହୋଇଥାଏ?<br><br>***Probes:*** *ଅନୁସନ୍ଧାନ:*<br>○ Staying at home more? ଘରେ ଅଧିକ ସମୟ ରହୁଛନ୍ତି? |

| | |
|---|---|
| ○ More/less workload? ଅଧିକ/କମ କାମ<br>○ More/less free time? ଅଧିକ/କମ ଖାଲି ସମୟ<br>○ Is there anywhere in the village you *used* to go but no longer go? ଗାଁରେ ଏପରି କିଛି ଜାଗା ଅଛି କି ଯେଉଠିକି ଆପଣ ଯାଉଥିଲେ କିନ୍ତୁ ଆଉ ଯାଉ ନାହାନ୍ତି? | |
| **C2.** Has coronavirus or the lockdown changed how you or your family **interact with other members of your village**? Please explain.<br><br>କଣ କରୋନାଭାଇରସ କିମ୍ବା ଲକଡାଉନ ଆପଣଙ୍କର କିମ୍ବା ଆପଣଙ୍କ ପରିବାରର ଗ୍ରାମର ଅନ୍ୟ ଲୋକମାନଙ୍କ ସହିତ ମିଳାମିଶାରେ କିଛି ପରିବର୍ତ୍ତନ ଆଣିଛି? ଦୟାକରି ବର୍ଣ୍ଣନା କରନ୍ତୁ। | ☐ 1. Yes ହଁ<br>☐ 2. No ନାହିଁ<br><br>**Please explain. ଦୟାକରି ବର୍ଣ୍ଣନା କରନ୍ତୁ।** |
| **C3.** In the past 7 days, did you take any of the following actions to cover your household's basic needs?<br>ଗତ ସାତଦିନ ମଧ୍ୟରେ, ନିଜ ଘରର ସାଧାରଣ ଆବଶ୍ୟକତାକୁ ପୂରଣ କରିବା ପାଇଁ, ଆପଣ ଉକ୍ତ କେଉଁ କେଉଁ ପଦକ୍ଷେପ ନେଇଛନ୍ତି?<br><br>Enumerator: read all response options.<br>**ଏନୁମରେଟର ନୋଟ** : ସମସ୍ତ ବିକଳ୍ପ ଉତ୍ତରଗୁଡିକୁ ପଢନ୍ତୁ<br><br>Record any explanations the respondent gives | ☐ 1. Use cash savings or bank savings ସଞ୍ଚିତ ଟଙ୍କା କିମ୍ବା ବ୍ୟାଙ୍କ ଜମାରାଶି ଖର୍ଚ୍ଚ କରିଛନ୍ତି<br>☐ 2. Borrow money or food from neighbor, friend, or relative ପଡୋଶୀ, ସାଙ୍ଗ କିମ୍ବା ସମ୍ପର୍କୀୟଙ୍କ ଠାରୁ ଟଙ୍କା କିମ୍ବା ଖାଦ୍ୟ ଉଧାର ଆଣିଛନ୍ତି<br>☐ 3. Sell assets ସମ୍ପତ୍ତି ବିକିଛନ୍ତି<br>☐ 4. Rely on Government assistance → **Ask follow-up question C3A** ସରକାରୀ ସାହାଯ୍ୟ ଉପରେ ନିର୍ଭର କରିଛନ୍ତି<br>☐ 5. Rely on NGO assistance ଏନଜିଓର ସାହାଯ୍ୟ ଉପରେ ନିର୍ଭର କରିଛନ୍ତି<br>☐ 6. None of the above ଏଥିମଧ୍ୟରୁ କିଛି ନୁହେଁ<br>☐ 7. Other ଅନ୍ୟାନ୍ୟ ________<br><br>**Record any explanations the respondent gives for this question: ଉତ୍ତରଦାତା ଦେଇଥିବା ଯେକୌଣସି ବର୍ଣ୍ଣନା କୁ ଟିପନ୍ତୁ :**<br><br>**If rely on government assistance is yes, ask:**<br>**ଯଦି ସରକାରୀ ସାହାଯ୍ୟ ଉପରେ ନିର୍ଭର କରୁଛନ୍ତି, ତାହେଲେ ପଚାରନ୍ତୁ :**<br><br>**C3A**. Which, if any, of the following government schemes have you received benefits from?<br>ଆପଣ ଉକ୍ତ କେଉଁ ସରକାରୀ ନୀତିରୁ ଲାଭବାନ ହେଇଛନ୍ତି?<br><br>Enumerator: read all response options.<br>**ଏନୁମରେଟର ଏନୁମରେଟର:** ସମସ୍ତ ବିକଳ୍ପ ଉତ୍ତରଗୁଡିକୁ ପଢନ୍ତୁ<br><br>☐ 1. National Social Assistance Programme for widows, senior citizens, differently abled |

### COVID-19 Mobile Phone Interview – VWSC

| | |
|---|---|
| | ଭାରତ ସରକାରଙ୍କର ବିଧବା, ବୃଦ୍ଧ ଓ ବିକଳାଙ୍ଗ ସାମାଜିକ ଭତ୍ତା କାର୍ଯ୍ୟକ୍ରମ<br>☐ 2. Free ration for BPL cardholders over three months<br>ବିପିଏଲ କାର୍ଡଧାରୀଙ୍କ ପାଇଁ 3 ମାସର ମାଗଣା ରାସନ<br>☐ 3. Jan Dhan account deposited with INR 500 for three months<br>ଜନଧନ ଖାତାରେ ତିନି ମାସ ପାଇଁ 500 ଟଙ୍କା ଜମା<br>☐ 4. Farmer accounts deposited with INR 2000<br>କୃଷକ ମାନଙ୍କ ବ୍ୟାଙ୍କ ଖାତାରେ 2000 ଟଙ୍କା ଜମା<br>☐ 5. Employment guarantee under National Rural Employment Guarantee Scheme<br>ଜାତୀୟ ଗ୍ରାମୀଣ କର୍ମନିଯୁକ୍ତି ଯୋଜନା (ନରେଗାଁ) ଦ୍ୱାରା ନିଶ୍ଚିତ ନିଯୁକ୍ତି<br>☐ 6. None କିଛି ନୁହେଁ |
| **C4.** Have you or any member of your family lost a job or work due to coronavirus or the lockdown?<br>କରୋନାଭାଇରସ ପାଇଁ, କଣ ଆପଣ କିମ୍ବା ଆପଣଙ୍କ ପରିବାରର କେହି ସଦସ୍ୟ ନିଜ ଚାକିରୀ କିମ୍ବା କାମ ହରେଇଛନ୍ତି କି?<br><br>If yes, who has lost their job and what was the occupation for the job that was lost?<br>ଯଦି ହଁ, କିଏ ନିଜ ଚାକିରୀ ହରେଇଛନ୍ତି ଓ ତାଙ୍କର ବୃତ୍ତି କଣ ଥିଲା? | ☐ 1. Yes ହଁ<br>☐ 2. No ନାହିଁ<br><br>**If yes, record relationship and occupation**:<br>ଯଦି ହଁ, ସମ୍ପର୍କ ଓ ବୃତ୍ତି ନୋଟ କରନ୍ତୁ: |
| **C5.** In the past 7 days, have you experienced any difficulties in getting the material you use to cook with (such as wood, gas, etc.) as a result of coronavirus or the lockdown?<br><br>ଗତ ସାତ ଦିନ ମଧ୍ୟରେ, କରୋନା ଭାଇରସ ପାଇଁ, ଆପଣଙ୍କୁ ରାନ୍ଧିବାରେ ଆବଶ୍ୟକ ହେଉଥିବା କିଛି ସାମଗ୍ରୀ (ଯେପରିକି କାଠ, ଗ୍ୟାସ ଇତ୍ୟାଦି) ମିଳିବାରେ କିଛି ଅସୁବିଧା ହୋଇଛି କି?<br><br>If yes, what type of material and please describe difficulty. ଯଦି ହଁ, କେଉଁ କି ପ୍ରକାର ବସ୍ତୁ ଓ ଦୟାକରି ବର୍ଣ୍ଣନା କରନ୍ତୁ କି ଅସୁବିଧା ହୋଇଛି ? | ☐ 1. Yes ହଁ<br>☐ 2. No ନାହିଁ<br><br>**If yes, what type of material and please describe difficulty**:<br>ଯଦି ହଁ, ଏହା କି ପ୍ରକାର ବସ୍ତୁ ଓ ଦୟାକରି ବର୍ଣ୍ଣନା କରନ୍ତୁ କି ଅସୁବିଧା ହୋଇଛି ? |
| **C6.** Has coronavirus or the lockdown changed the **types or quantity of food** your family eats daily? If yes, describe how this has changed.<br><br>କଣ କରୋନାଭାଇରସ କିମ୍ବା ଲକଡାଉନ ଆପଣଙ୍କ ପରିବାରର ର ଖାଇବାର ପରିମାଣ କିମ୍ବା ପ୍ରକାରରେ କିଛି ପରିବର୍ତ୍ତନ ଆଣିଛି? ଯଦି ହଁ, ବର୍ଣ୍ଣନା କରନ୍ତୁ କି ଏହା କି ପରିବର୍ତ୍ତନ ଆଣିଛି। | Change in **types of food**? ଖାଦ୍ୟର ପ୍ରକାରରେ ପରିବର୍ତ୍ତନ<br>☐ 1. Yes ହଁ<br>☐ 2. No ନାହିଁ<br><br>Change in **quantity of food**? ଖାଦ୍ୟର ପରିମାଣରେ ପରିବର୍ତ୍ତନ<br>☐ 1. Yes ହଁ – Increase<br>☐ 2. Yes ହଁ - Decrease<br>☐ 3. No ନାହିଁ<br><br>**If yes, describe:** ଯଦି ହଁ, ବର୍ଣ୍ଣନା କରନ୍ତୁ |

### COVID-19 Mobile Phone Interview – VWSC

| | |
|---|---|
| **C7.** Are the number of people currently staying in your house more, less, or the same as the number of people that were staying in your house before coronavirus and the lockdown started?<br><br>ଆପଣଙ୍କ ଘରେ ବର୍ତ୍ତମାନ ରହୁଥିବା ଲୋକଙ୍କ ସଂଖ୍ୟା କରୋନାଭାଇରସ ଏବଂ ଲକଡାଉନ ଆରମ୍ଭ ହେବ ପୂର୍ବରୁ ଘରେ ରହୁଥିବା ଲୋକଙ୍କ ସଂଖ୍ୟା ଠାରୁ ଅଧିକ**,** କମ ନା ସମାନ**?** | ☐ 1. Less କମ<br>☐ 2. The same ସମାନ<br>☐ 3. More ଅଧିକ |
| **C8.** The last time you defecated, did you defecate in the open or use the latrine?<br><br>କଣ ଶେଷ ଥର ଯେବେ ଆପଣ ଝାଡା ଯାଇଥିଲେ **,**ତେବେ ଆପଣ ଖୋଲା ରେ ଝାଡା ଯାଇଥିଲେ ନା ପାଇଖାନା ବ୍ୟବହାର କରିଥିଲେ**?** | ☐ 1. Open ଖୋଲା ରେ<br>☐ 2. Latrine ପାଇଖାନା ରେ<br>☐ 3. Somewhere else (not open field or latrine) ଅଲଗା କେଉଁଠି (ଖୋଲା ପଡିଆ ନୁହେଁ କିମ୍ବା ପାଇଖାନା ନୁହେଁ)<br>☐ 99. Don't know ଜାଣିନାହିଁ |
| **C9.** Has coronavirus or the lockdown changed the **defecation location of you or any family members?** Please explain.<br><br>କଣ କରୋନାଭାଇରସ କିମ୍ବା ଲକଡ଼ାଉନ ଆପଣ କିମ୍ବା ଆପଣଙ୍କ କେହି ପରିବାର ସଦସ୍ୟଙ୍କ ଝାଡା ଯିବା ଜାଗାକୁ ବଦଳେଇଛି କି? ଦୟାକରି ବର୍ଣ୍ଣନା କରନ୍ତୁ<br><br>**Probe**: Have there been any changes for **elderly adults** in your household?<br>Have there been any changes for **young children** in your household?<br><br>ଆପଣଙ୍କ ଘରର **ବୃଦ୍ଧ ବ୍ୟକ୍ତିଙ୍କ** କ୍ଷେତ୍ରରେ କିଛି ପରିବର୍ତ୍ତନ ଆସିଛି କି**?**<br>ଆପଣଙ୍କ ଘରର **ଛୋଟ ପିଲାଙ୍କ** କ୍ଷେତ୍ରରେ କିଛି ପରିବର୍ତ୍ତନ ଆସିଛି କି**?** | ☐ 1. Yes ହଁ<br>☐ 2. No ନାହିଁ<br><br>**Please explain. ବର୍ଣ୍ଣନା କରନ୍ତୁ** |
| **C10.** Due to coronavirus or the lockdown, do you think people in this village are defecating in the open more, defecating in a latrine more, or there has been no change in where people defecate? Explain why.<br><br>ଆପଣ ଭାବୁଛନ୍ତି କି ଯେ, କରୋନାଭାଇରସ କିମ୍ବା ଲକଡ଼ାଉନ ପାଇଁ ଆପଣଙ୍କ ଗ୍ରାମର ଲୋକମାନେ, ଖୋଲାରେ ଅଧିକ ଝାଡା ଯାଉଛନ୍ତି, ଲାଟ୍ରିନର ରେ ଅଧିକ ଝାଡା ଯାଉଛନ୍ତି ନା ଲୋକ ଝାଡା କରୁଥିବା ଜାଗାରେ କୌଣସି ପରିବର୍ତ୍ତନ ଆସିନାହିଁ? ବର୍ଣ୍ଣନା କରନ୍ତୁ କଣ ପାଇଁ। | ☐ 1. More open defecation ଖୋଲରେ ଝାଡା ଯିବା ବଢ଼ିଛି<br>☐ 2. More latrine use ଲାଟ୍ରିନ ବ୍ୟବହାର ବଢ଼ିଛି<br>☐ 3. No change କିଛି ପରିବର୍ତ୍ତନ ନୁହେଁ<br>☐ 99. Don't know ଜାଣିନାହିଁ<br><br>**Explain why: ବର୍ଣ୍ଣନା କରନ୍ତୁ କାହିଁକି** |
| **C11.** Has coronavirus or the lockdown changed your **handwashing practices**? Please explain. | ☐ 1. Yes ହଁ<br>☐ 2. No ନାହିଁ |

| | |
|---|---|
| କଣ କରୋନାଭାଇରସ କିମ୍ବା ଲକଡ଼ାଉନ ଆପଣଙ୍କ **ହାତଧୁଆ ପ୍ରକ୍ରିୟାକୁ** ବଦଳେଇଛି? ଦୟାକରି ବର୍ଣ୍ଣନା କରନ୍ତୁ।<br><br>**Probes**: Has there been any change in **how often** you wash your hands?<br><br>କଣ ଆପଣ ପ୍ରାୟତଃ କେତେଥର ହାତ ଧୁଅନ୍ତି, ସେଥିରେ କିଛି ପରିବର୍ତ୍ତନ ଆସିଛି କି **?**<br><br>Has there been any change in **how much soap or water you use** to wash your hands?<br><br>କଣ ଆପଣ ପ୍ରାୟତଃ କେତେ ପରିମାଣର ସାବୁନ କିମ୍ବା ପାଣି ବ୍ୟବହାର କରନ୍ତି, ସେଥିରେ କିଛି ପରିବର୍ତ୍ତନ ଆସିଛି କି**?**<br>? | **<u>Please explain. ଦୟାକରି ବର୍ଣ୍ଣନା କରନ୍ତୁ</u>** |
| **C12.** In the past 7 days, have you had any difficulties with getting the water that you need for your household? Please explain.<br><br>କରୋନାଭାଇରସ କିମ୍ବା ଲକଡାଉନ ପାଇଁ ଗତ ସାତ ଦିନ ମଧ୍ୟରେ, କଣ ଆପଣଙ୍କୁ ନିଜ ଘରର ଆବଶ୍ୟକତା ପାଇଁ ଦରକାର ହେଉଥିବା ପାଣି ମିଳିବାରେ କିଛି ଅସୁବିଧା ହେଇଛି କି **?** | ☐ 1. Yes ହଁ<br>☐ 2. No ନାହିଁ<br><br>**<u>Please explain. ଦୟାକରି ବର୍ଣ୍ଣନା କରନ୍ତୁ</u>** |
| **C13.** Has coronavirus or the lockdown changed your **drinking water practices** such as where you get your water, how you store it, and whether or not you clean/treat the water? Please explain.<br><br>କଣ କରୋନାଭାଇରସ କିମ୍ବା ଲକଡ଼ାଉନ ଆପଣଙ୍କ **ପାଣି ପିଇବା ସହିତ ଜଡିତ ପ୍ରକ୍ରିୟା ମାନଙ୍କୁ** ବଦଳେଇଛି କି, ଯେପରିକି ଆପଣ ପାଣି କେଉଁଠାରୁ ଆଣନ୍ତି, ତାକୁ କିପରି ରଖନ୍ତି କିମ୍ବା କିପରି ପରିଷ୍କାର କରନ୍ତି? ଦୟାକରି ବର୍ଣ୍ଣନା କରନ୍ତୁ। | Change in **where you get** your drinking water?<br>**ଆପଣ ପିଇବା ପାଣି ଆଣୁଥିବା ସ୍ଥାନରେ ପରିବର୍ତ୍ତନ**?<br>☐ 1. Yes ହଁ<br>☐ 2. No ନାହିଁ<br><br>Change in **how you store** your drinking water?<br>ଆପଣ କିପରି ପିଇବା ପାଣିକୁ ରଖନ୍ତି, ସେଥିରେ ପରିବର୍ତ୍ତନ?<br>☐ 1. Yes ହଁ<br>☐ 2. No ନାହିଁ<br><br>Change in whether or not you **clean/treat** your drinking water?<br>ଆପଣ ନିଜ ପିଇବା ପାଣିକୁ କିପରି ସଫା/ପରିଷ୍କାର କରନ୍ତି, ସେଥିରେ ପରିବର୍ତ୍ତନ?<br>☐ 1. Yes ହଁ<br>☐ 2. No ନାହିଁ<br><br>**<u>Please explain. ଦୟାକରି ବର୍ଣ୍ଣନା କରନ୍ତୁ</u>** |

| | |
|---|---|
| **C14.** Has coronavirus or the lockdown changed your **household cleaning practices**? Please explain.<br><br>କଣ କରୋନାଭାଇରସ କିମ୍ବା ଲକଡ଼ାଉନ ଆପଣଙ୍କ **ଘର ସଫା କରିବା ପ୍ରକ୍ରିୟା**କୁ ବଦଳେଇଛି? ଦୟାକରି ବର୍ଣ୍ଣନା କରନ୍ତୁ<br><br>**Probes**: Has there been any change in **how often** you clean your home?<br><br>କଣ ଆପଣ ପ୍ରାୟତଃ କେତେଥର ଘର ସଫା କରନ୍ତି, ସେଥିରେ କିଛି ପରିବର୍ତ୍ତନ ଆସିଛି କି **?**<br><br>Has there been any change in **how much detergent/disinfectant or water you use** to clean your home?<br><br>କଣ ଆପଣ ଘର ସଫା କରିବା ପାଇଁ ପ୍ରାୟତଃ ଯେତେ ଲୁଗାଧୁଆ ପାଉଡର/ ଫିନାଇଲ କିମ୍ବା ପାଣି ବ୍ୟବହାର କରନ୍ତି, ସେଥିରେ କିଛି ପରିବର୍ତ୍ତନ ହେଇଛି କି**?** | ☐ 1. Yes ହଁ<br>☐ 2. No ନାହିଁ<br><br>**Please explain.** **ଦୟାକରି ବର୍ଣ୍ଣନା କରନ୍ତୁ** |
| **C15.** Do you currently have soap or detergent in your household?<br><br>ଆପଣଙ୍କ ଗହରେ ବର୍ତ୍ତମାନ ସାବୁନ କିମ୍ବା ଲୁଗାଧୁଆ ପାଉଡର ଅଛି କି**?** | ☐ 1. Yes ହଁ<br>☐ 2. No ନାହିଁ → **Ask followup question C15A**<br>☐ 99. Don't know ଜାଣି ନାହିଁ<br><br>**If answered no, ask: ଯଦି ଉତ୍ତରରେ 'ନାହିଁ' କୁହନ୍ତି, ତା'ହେଲେ ପଚାରନ୍ତୁ :**<br><br>C15A. The last time you needed to purchase soap for your household, did you experience any of the following<br><br>ଶେଷଥର ପାଇଁ, ଯେବେ ଆପଣ ନିଜ ଘର ପାଇଁ ସାବୁନ କିଣିବାକୁ ଭାବିଲେ, କଣ ଆପଣ ଉକ୍ତ ଅନୁଭବ କରିଛନ୍ତି କି**?**<br><br>Enumerator: read all response options. Record any explanations the respondent gives.<br>**ଏନୁମରେଟର ନୋଟ** : ସମସ୍ତ ବିକଳ୍ପ ଉତ୍ତରଗୁଡ଼ିକୁ ପଢନ୍ତୁ । ଉତ୍ତରଦାତା ଦେଇଥିବା ଯେକୌଣସି ବର୍ଣ୍ଣନା କୁ ଟିପନ୍ତୁ<br><br>☐ 1. Difficulties in going to obtain **soap** due to mobility restrictions imposed by government<br>ସରକାରଙ୍କ ପ୍ରତିବନ୍ଧକ ମାନଙ୍କ ପାଇଁ ଦୋକାନକୁ ସାବୁନ କିଣିବାକୁ ଯିବାରେ ଅସୁବିଧା ହେଉଛି<br>☐ 2. Unable to buy the amount of **soap** we usually buy because the household income has dropped<br>ଘରର ଆୟ କମିଯିବା ଯୋଗୁଁ ଆମେ ସାଧାରଣତଃ ଯେତେ ସାବୁନ କିଣୁ, ତାହା କିଣିପାରୁନାହୁଁ<br>☐ 3. Unable to buy the amount of **soap** we usually buy because of limited supply<br>ସୀମିତ ଯୋଗାଣ ଯୋଗୁଁ ଆମେ ସାଧାରଣତଃ ଯେତେ ସାବୁନ କିଣୁ, ତାହା କିଣିପାରୁନାହୁଁ |

| | |
|---|---|
| | ☐ 4. None of the above ଏଥିମଧ୍ୟରୁ କିଛି ନୁହେଁ<br><br>**Record any explanations the respondent gives for this question:**<br>ଏହି ପ୍ରଶ୍ନ ପାଇଁ ଉତ୍ତରଦାତା ଦେଇଥିବା ଯେକୌଣସି ବର୍ଣ୍ଣନା କୁ ଟିପନ୍ତୁ। |
| **SECTION D: Household Information**<br><br>**Enumerator read:** Thank you for your responses about coronavirus. Now I'm going to ask you some questions about your household. This is the last section of the survey!<br><br>କରୋନାଭାଇରସ ଉପରେ ଆପଣଙ୍କ ଉତ୍ତର ପାଇଁ ଧନ୍ୟବାଦ। ମୁଁ ବର୍ତ୍ତମାନ ଆପଣଙ୍କୁ କିଛି ପ୍ରଶ୍ନ ନିଜ ଘର ବିଷୟରେ ପଚାରିବି। ଏହା ସର୍ଭେ ର ଶେଷ ଭାଗ। | |
| **D1.** What is the highest level of education that you have had?<br><br>ଆପଣ ସର୍ବୋତ୍ତମ କେତେ ପାଠ ପଢିଛନ୍ତି? | ☐ 1. No formal schooling ସ୍କୁଲ ଯାଇନାହାନ୍ତି<br>☐ 2. Some primary education କିଛି ପ୍ରାଥମିକ ପାଠ<br>☐ 3. Completed primary education ପ୍ରାଥମିକ ପାଠ ଶେଷ<br>☐ 4. Some secondary school କିଛି ଟା ସେକେଣ୍ଡରି ପାଠ<br>☐ 5. Matriculation (10th class completed) ମେଟ୍ରିକୁଲେଶନ<br>☐ 6. Completed secondary school କିଛି ଟା ସେକେଣ୍ଡରି ପାଠ ଶେଷ<br>☐ 7. Some college କିଛି କଲେଜ<br>☐ 8. Completed college/university କଲେଜ/ୟୁନିଭରସିଟି ଶେଷ<br>☐ 99. Don't know ଜାଣି ନାହିଁ |
| **D2.** What is your occupation?<br><br>ଆପଣଙ୍କ ବୃତ୍ତି କଣ ? | ☐ 1. Unemployed (no work that earns money)<br>ବେରୋଜଗାର(ରୋଜଗାର କରିଲା ଭଳିଆ କିଛି କାମ କରନ୍ତିନାହିଁ)<br>☐ 2. Self-Employed (Work in home: agriculture, potter, etc.)<br>ନିଜସ୍ୱ ରୋଜଗାର(ଯେମିତିକି- ଚାଷ, କୁମ୍ଭାର, ଅନ୍ୟାନ୍ୟ)<br>☐ 3. Employed (Work outside the home: office, labourer, factory, etc.)<br>ଚାକିରି(ଘରଠୁ ବାହାରେ କାମ କରନ୍ତି : ଉଦାହରଣ –ଅଫିସ, ଫ୍ୟାକ୍ଟ୍ରି, ଅନ୍ୟାନ୍ୟ)<br>☐ 4. Student ଛାତ୍ର |
| **D3.** What is the caste or tribe of the household?<br>ଆପଣଙ୍କର କେଉଁ - *ଜାତି ବା ଜନଜାତି* ?<br><br>Enumerator note: Read response options<br>ଏନୁମରେଟର ନୋଟ : ଉତ୍ତର ଅପସନସ୍ ଗୁଡିକୁ ପଢନ୍ତୁ । | ☐ 1. Brahmin ବ୍ରାହ୍ମଣ<br>☐ 2. General ସାଧାରଣ<br>☐ 3. Scheduled caste ଅନୁସୂଚିତ ଜାତି<br>☐ 4. Other backward caste ଅନ୍ୟ ପଛୁଆ ବର୍ଗ<br>☐ 5. Scheduled tribe ଅନୁସୂଚିତ ଜନଜାତି<br>☐ 6. Do not want to indicate caste ଜାତି କହିବାକୁ ଚାହୁଁନାହାଁନ୍ତି<br>☐ 77. Not applicable ଉପଯୁକ୍ତ ନୁହେଁ<br>☐ 88. Other ଅନ୍ଯାନ୍ଯ: ________<br>☐ 99. Don't know ଜାଣି ନାହିଁ |

| | |
|---|---|
| **D4.** What is the religion of the household?<br>ଆପଣଙ୍କର *କେଉଁ ଧର୍ମ*? | ☐ 1. Hindu ହିନ୍ଦୁ<br>☐ 2. Muslim ମୁଁସଲମାନ୍<br>☐ 3. Christian ଖ୍ରୀଷ୍ଟିଆନ୍<br>☐ 4. Sikh ଶିଖ୍<br>☐ 5. Buddhist/Neo-buddhist ବୌଦ୍ଧ ଧର୍ମ<br>☐ 6. Jain ଜୈନ ଧର୍ମ<br>☐ 7. Jewish ଜୀଉ ଧର୍ମ<br>☐ 8. Parsi/Zorastrian ପାର୍ସୀ ଧର୍ମ<br>☐ 9. No Religion କୌଣସି ଧର୍ମ ନୁହେଁ<br>☐ 88. Other ଅନ୍ୟାନ୍ୟ: ________ |
| **D5.** Do you have an Antodaya Card and/or ration card? (select all that apply)<br>ଆପଣଙ୍କର ଅନ୍ତୋଦୟ /ରାସନ୍ କାର୍ଡ ଅଛି କି? | ☐ 1. Antodaya ଅନ୍ତୋଦୟ<br>☐ 2. Ration card ରାସନ କାର୍ଡ<br>☐ 3. None କିଛି ନାହିଁ<br>☐ 99. Don't know ଜାଣି ନାହିଁ |
| **END OF INTERVIEW:** Thank the respondent for their time and give them the COVID-19 information and resources summary provided by Gram Vikas.<br><br>ସାକ୍ଷାତକରର ଶେଷ: ଉତ୍ତରଦାତାଙ୍କୁ ତାଙ୍କ ସମୟ ପାଇଁ ଧନ୍ୟବାଦ ଜଣାନ୍ତୁ ଏବଂ ତାଙ୍କୁ ଗ୍ରାମବିକାଶ ଦ୍ୱାରା ଦିଆଯାଇଥିବା COVID-19 ସୂଚନା ବିଷୟରେ ଅବଗତ କରନ୍ତୁ | |
| **End Time**<br>ଶେଷ ସମୟ | |

### COVID-19 Mobile Phone Interview – Primary Caregiver

| SECTION A: Respondent and Enumerator Information & Consent Process | |
|---|---|
| **A1.** Village ID ଗ୍ରାମ ର କୋଡ: | **A2.** Household ID ଘର ଘରର ସଂଖ୍ୟା |
| **A3.** Village Name ଗ୍ରାମ ର ନାମ | |
| **A4.** Date ତାରିଖ | (y/d/m) |
| **A5.** Start Time ଆରମ୍ଭ ସମୟ | |
| **A6.** Enumerator name ଆପଣଙ୍କ ନାମ | ☐ 1. Alok<br>☐ 2. Dhiren<br>☐ 3. Varsha |
| **A7.** Confirm that the respondent and their family (if applicable) currently live in the intended village (and are not currently staying somewhere else).<br>ପୁଷ୍ଟି କରନ୍ତୁ କି ଯେ, ଉପଯୁକ୍ତ ଉତ୍ତରଦାତା ଏବଂ ତାଙ୍କ ପରିବାର (ଯଦି ଲାଗୁ ହୁଏ)ବର୍ତ୍ତମାନ ଉଦ୍ଦିଷ୍ଟ ଗ୍ରାମରେ ହିଁ ବସବାସ କରନ୍ତି (ଏବଂ ଅନ୍ୟ କେଉଁଠି ବସବାସ କରନ୍ତି ନାହିଁ) | ☐ 1. Yes ହଁ<br>☐ 2. No – respondent/family are staying outside the village and left *before* the pandemic → **END SURVEY**<br>ନା - ଉତ୍ତରଦାତା ଓ ତାଙ୍କ ପରିବାର ଗ୍ରାମ ବାହାରେ ବସବାସ କରନ୍ତି ଏବଂ ସର୍ବବ୍ୟାପୀ ମହାମାରୀ **ହେବା ପୂର୍ବରୁ** ଗ୍ରାମ ଛାଡି ଦେଇଛନ୍ତି - **ସର୍ଭେ ଶେଷ**<br>☐ 3. No – respondent/family are staying outside the village and left *because of* the pandemic → **END SURVEY**<br>ନା- ଉତ୍ତରଦାତା ଓ ତାଙ୍କ ପରିବାର ଗ୍ରାମ ବାହାରେ ବସବାସ କରନ୍ତି ଏବଂ ସର୍ବବ୍ୟାପୀ ମହାମାରୀ **ହେବା କାରଣରୁ** ଗ୍ରାମ ଛାଡି ଦେଇଛନ୍ତି - **ସର୍ଭେ ଶେଷ** |
| **A8a.** Read the consent form to the respondent and then ask for their consent to participate.<br>ଉତ୍ତରଦାତାଙ୍କ ପାଇଁ ସହମତି ପତ୍ରଟିକୁ ପଢନ୍ତୁ ଏବଂ ତାଙ୍କୁ **ସର୍ଭେରେ** ଅଂଶଗ୍ରହଣ ପାଇଁ ସହମତି ମାଗନ୍ତୁ।<br><br>Does the respondent consent to the survey?<br>କ'ଣ ଉତ୍ତରଦାତା ସର୍ଭେରେ ଅଂଶଗ୍ରହଣ ପାଇଁ ସହମତି ପ୍ରଦାନ କଲେ? | ☐ 1. Yes ହଁ<br>☐ 2. No → **END SURVEY** ନା - **ସର୍ଭେ ଶେଷ** |
| **A8b.** Does the respondent consent to having the phone call audio recorded?<br>କଣ ଉତ୍ତରଦାତା ଫୋନକଲର ରେକଡିଙ୍ଗ ପାଇଁ ସହମତି ପ୍ରଦାନ କଲେ? | ☐ 1. Yes ହଁ<br>☐ 2. No ନା → Pause when you need to in order to record detailed responses! ଯେତେବେଳେ ଉତ୍ତର ଲେଖିବା ଆବଶ୍ୟକ ପଡେ, ବିରାମ ନିଅନ୍ତୁ। |
| **ENUMERATOR NOTE:** Now you are going to begin the survey. ଏନୁମରେଟରଙ୍କ ପାଇଁ ନୋଟ: ବର୍ତ୍ତମାନ ଆପଣ ସର୍ଭେ ଆରମ୍ଭ କରିବେ | |
| **A9.** What is your name?<br>ଆପଣଙ୍କ ନାମ କଣ ? | |
| **A10.** What is the sex of the respondent?<br>ଉତ୍ତରଦାତଙ୍କ ଲିଙ୍ଗ କଣ?<br>***Enumerator Note:*** Clarify with the respondent if you are not sure. ଏନୁମରେଟର ନୋଟ - ସ୍ପଷ୍ଟ କରନ୍ତୁ ଯେବେ ଆପଣ ନିଶ୍ଚିତ ନୁହନ୍ତି | ☐ 1. Male ପୁରୁଷ<br>☐ 2. Female ମହିଳା |
| **A11.** What is your age? ଆପଣଙ୍କ ବୟସ କେତେ? | ____________ (years) (ବର୍ଷ) |

### COVID-19 Mobile Phone Interview – Primary Caregiver

| | |
|---|---|
| **A12.** Including yourself, how many people are currently staying in your house?<br><br>ଆପଣଙ୍କୁ ମିଶେଇ, ବର୍ତ୍ତମାନ ଆପଣଙ୍କ ଘରେ କେତେଜଣ ଲୋକ ରୁହନ୍ତି? | ______________ |
| **A13.** How many children <5 years old currently live in your household?<br><br>ପାଞ୍ଚ ବର୍ଷରୁ କମ ବୟସର କେତେଜଣ ପିଲା ବର୍ତ୍ତମାନ ଏହି ଘରେ ବସବାସ କରନ୍ତି? | ______________<br><br>**Record Age of each child <5:**<br><br>Child 1: Years: ________ Months: _________<br>Child 2: Years: ________ Months: _________<br>Child 3: Years: ________ Months: _________ |
| **A14.** Is anyone in your household a daily wage labourer who usually works in another state?<br><br>କଣ ଆପଣଙ୍କ ଘରର କେହି ଦିନ ମଜୁରିଆ ଅଟନ୍ତି କି ଯେ ସାଧାରଣତଃ ଅନ୍ୟ ରାଜ୍ୟରେ କାମ କରନ୍ତି ? | ☐ 1. Yes ହଁ - And they are still residing in another state ହଁ- ସେମାନେ ବର୍ତ୍ତମାନ ମଧ୍ୟ ଅନ୍ୟ ରାଜ୍ୟରେ ରହୁଛନ୍ତି<br>☐ 2. Yes ହଁ - But now they reside in Odisha ହଁ- କିନ୍ତୁ ସେମାନେ ବର୍ତ୍ତମାନ ଓଡ଼ିଶାରେ ରୁହନ୍ତି<br>☐ 3. No ନା<br><br>**→ If Yes (1 or 2): ଯଦି ଉତ୍ତର ହଁ (1 କିମ୍ବା 2)**<br>How is this household member related to you?<br>(for example, husband, brother-in-law, etc.)<br><br>ଘରର ଏହି ସଦସ୍ୟ ଆପଣଙ୍କର କଣ ଲାଗିବେ?<br>(ଉଦାହରଣ ସ୍ୱରୂପ: ସ୍ୱାମୀ, ଦିଅର, ଇତ୍ୟାଦି) |
| **A15.** Do you have piped water to your household or compound?<br><br>କଣ ଆପଣଙ୍କ ଘରକୁ ପାଇପ ଯୋଗେ ପାଣିର ବ୍ୟବସ୍ଥା ଅଛି? | ☐ 1. Yes – functional ହଁ- ସକ୍ରିୟ<br>☐ 2. Yes – not functional ହଁ- ନିଷ୍କ୍ରିୟ<br>☐ 3. No ନା<br><br>**If 2 or 3:**<br>What is your primary source of water and where is it located?<br><br>ଯଦି 2 କିମ୍ବା 3:<br>ଆପଣଙ୍କ ଘରର ମୁଖ୍ୟ ଜଳଉତ୍ସଟି କଣ ଓ ଏହା କେଉଁ ଠାରେ ଅବସ୍ଥିତ?<br><br>Source (ଉତ୍ସ):<br>☐ 1. Public tap ପବ୍ଲିକକଳ/ସର୍ବସାଧାରଣଟ୍ୟାପ୍<br>☐ 2. Well/borehole/spring କୂଅ, ନଳକୂଅ, ଝରଣା<br>☐ 3. Surface water (river/dam/lake/pond/stream/canal) ଭୂତଳଜଳ(ନଦୀ/ବନ୍ଧ/ହ୍ରଦ/ପୋଖରୀ/ନାଳ/କେନାଲ)<br>☐ 4. Rainwater ବର୍ଷାପାଣି |

| | |
|---|---|
| | ☐ 5. Bottled water ବୋତଲପାଣି<br>☐ 6. Other ଅନ୍ୟାନ୍ୟ<br>☐ 7. Don't know ଜାଣିନାହିଁ<br><br>Location: ଅବସ୍ଥାନ<br>☐ 1. In own dwelling ନିଜ ରହିବା ଜାଗାରେ<br>☐ 2. In own yard/plot/compound ନିଜଜାଗା/ପ୍ଲଟରେ/ଅଗଣାରେ<br>☐ 3. Outside the household compound ଘର ଅଗଣା/ପରିସର ବାହାରେ |
| **A16.** Does your household have a latrine?<br><br>କଣ ଆପଣଙ୍କ ଘରେ ପାଇଖାନା ଅଛି? | ☐ 1. Yes – functional ହଁ- ସକ୍ରିୟ<br>☐ 2. Yes – not functional ହଁ- ନିଷ୍କ୍ରିୟ<br>☐ 3. No ନାହିଁ |
| **A17.** Do you have an enclosed bathing area in your household compound?<br><br>କଣ ଆପଣଙ୍କ ଘର ପରିସର ମଧ୍ୟରେ ଏକ ଆବଦ୍ଧ ଗାଧୁଆଘର ଅଛି କି? | ☐ 1. Yes ହଁ<br>☐ 2. No ନା |
| **A18.** Do you currently have soap or detergent in your household?<br><br>ଆପଣଙ୍କ ଗହରେ ବର୍ତ୍ତମାନ ସାବୁନ କିମ୍ବା ଲୁଗାଧୁଆ ପାଉଡର ଅଛି କି**?** | ☐ 1. Yes ହଁ<br>☐ 2. No ନାହିଁ<br>☐ 99. Don't know ଜାଣି ନାହିଁ |
| **SECTION B: COVID-19 Knowledge, Perceptions, and Actions**<br>**ENUMERATO READ:** Thank you for your responses so far. Now I'm going to ask you some questions related to coronavirus.<br>ଏନୁମରେଟର ପଢନ୍ତୁ : ବର୍ତ୍ତମାନ ପର୍ଯ୍ୟନ୍ତ ଦେଇଥିବା ଉତ୍ତର ମାନଙ୍କ ପାଇଁ ଧନ୍ୟବାଦ। ବର୍ତ୍ତମାନ, ମୁଁ ଆପଣଙ୍କୁ କରୋନାଭାଇରସ ସହିତ ଜଡିତ କିଛି ପ୍ରଶ୍ନ ପଚାରିବି। | |
| **B1.** Have you heard of a disease called covid-19, coronavirus, or corona?<br><br>କଣ ଆପଣ କୋଭିଡ-୧୯, କରୋନାଭାଇରସ କିମ୍ବା କରୋନା ନାମକ ରୋଗ ବିଷୟରେ ଶୁଣିଛନ୍ତି? | ☐ 1. Yes ହଁ<br>☐ 2. No → **If no, explain that this is the disease that has led to the recent lockdowns in India.** ନା → ଯଦି ନା, ତାହେଲେ ଏହା କୁହନ୍ତୁ କି ଯେ ଏହି ରୋଗ ପାଇଁ ହିଁ ଏବେ ଭାରତରେ ଲକଡାଉନ ହୋଇଛି |
| **B2.** What are the symptoms of coronavirus?<br>କରୋନାଭାଇରସର ଲକ୍ଷଣ ଗୁଡିକ କଣ?<br><br>***Probe:*** Anything else?<br>ପ୍ରୋବ : ଆଉ କିଛି?<br><br>***Enumerator Notes:***<br>Do **NOT** read response options. Select all that apply.<br>ସମ୍ଭାବ୍ୟ ଉତ୍ତର ଗୁଡିକୁ ପଢନ୍ତୁ ନାହିଁ. ଦେଇଥିବା ଉତ୍ତର ଗୁଡିକୁ ଟୀକ କରନ୍ତୁ.<br><br>If respondent says sick/flu as their response, ask them to specify specific symptoms. | ☐ 1. No symptoms କିଛି ଲକ୍ଷଣ ନାହିଁ<br>☐ 2. Fever ଜ୍ୱର<br>☐ 3. Cough କାଶ<br>☐ 4. Dry cough ଶୁଖିଲା କାସ<br>☐ 5. Headache ମୁଣ୍ଡବଥା<br>☐ 6. Body aches ଶରୀର ବ୍ୟଥା<br>☐ 7. Difficulty breathing ନିଶ୍ୱାସ ନେବାରେ ଅସୁବିଧା<br>☐ 8. Chest pain ଛାତି ଯନ୍ତ୍ରଣା<br>☐ 9. Tiredness କ୍ଳାନ୍ତି<br>☐ 10. Diarrhea ଡ଼ାଇରିଆ<br>☐ 11. Chills ଥଣ୍ଡା ଲାଗିବା<br>☐ 12. Rash କୁଣ୍ଡିଆ |

| | |
|---|---|
| ଯଦି ଉତ୍ତରଦାତା ଅସୁସ୍ଥ ଲାଗିବା/ ଥଣ୍ଡା ହେବା ବିଷୟରେ କୁହନ୍ତି, ତାଙ୍କୁ ନିର୍ଦ୍ଦିଷ୍ଟ ଲକ୍ଷଣ ଗୁଡିକ ବିଷୟରେ କହିବାକୁ କୁହନ୍ତୁ | ☐ 13. Loss of smell and/or taste ଗନ୍ଧ କିମ୍ବା ସ୍ୱାଦ ବାରିପାରୁନାହାନ୍ତି<br>☐ 14. Dizziness ମୁଣ୍ଡ ବୁଲେଇବା<br>☐ 15. Sneezing ଛିଙ୍କ/ଚିଙ୍କ<br>☐ 16. Sore throat ଖରାପ ଗଳା<br>☐ 88. Other ଅନ୍ୟାନ୍ୟ:<br>☐ 99. Don't know ଜାଣିନାହିଁ |
| **B3.** What causes a person to get sick with coronavirus?<br><br>କେଉଁ କାରଣ ମାନଙ୍କ ପାଇଁ ଜଣଙ୍କୁ କରୋନା ଭାଇରସ ହୋଇଥାଏ?<br><br>***Enumerator Notes:***<br>Probe for respondent to describe in detail. If the respondent is having trouble answering, ask: "What actions would increase your risk of getting coronavirus? Why?"<br><br>ଏନୁମରେଟରଙ୍କ ପାଇଁ ନୋଟ:<br>ଉତ୍ତରଦାତାଙ୍କୁ ବର୍ଣ୍ଣନା କରିବା ପାଇଁ ଅନୁସନ୍ଧାନ କରନ୍ତୁ। ଯଦି ଉତ୍ତରଦାତାଙ୍କୁ ଉତ୍ତର ଦେବାରେ ଅସୁବିଧା ହେଉଥାଏ, ପଚାରନ୍ତୁ, "କେଉଁ କମ ଆପଣଙ୍କର କରୋନାଭାଇରସ ହେବାର ଆଶଙ୍କାକୁ ବଢେଇଦେଇଥାଏ? କାହିଁକି?" | |
| **B4.** Do you think your personal chance of getting coronavirus is low, medium, high, or do you have no risk at all? Explain why.<br><br>ଆପଣଙ୍କୁ ଲାଗୁଛି କି ଯେ ଆପଣଙ୍କୁ କରୋନାଭାଇରସ ହେବାର ବ୍ୟକ୍ତିଗତ ସମ୍ଭାବନା କମ, ମଧ୍ୟମ, ଅଧିକ ଅଥବା ଆପଣଙ୍କ ପ୍ରତି କିଛି ବି ବିପଦ ନାହିଁ? ବର୍ଣ୍ଣନା କରନ୍ତୁ କାହିଁକି। | ☐ 1. Low କମ<br>☐ 2. Medium ମାଧ୍ୟମ<br>☐ 3. High ଅଧିକ<br>☐ 4. No risk କିଛି ବିପଦ ନୁହେଁ<br>☐ 99. Don't know ଜାଣିନାହିଁ<br><br>**Explain why: ବର୍ଣ୍ଣନା କରନ୍ତୁ କାହିଁକି** |
| **B5.** What types of people are at high risk for getting very seriously sick if they get coronavirus?<br><br>କେଉଁ ପ୍ରକାର ଲୋକଙ୍କୁ କରୋନା ଭାଇରସ ହେଲେ ସେମାନେ ସର୍ବାଧିକ ଅସୁସ୍ଥ ହେବାର ବିପଦ ଅଛି?<br><br>***Enumerator Notes:***<br>Do **NOT** read response options. Select all that apply.<br>ଏନୁମରେଟରଙ୍କ ପାଇଁ ନୋଟ: ବିକଳ୍ପ ଉତ୍ତର ଗୁଡିକୁ ପଢନ୍ତୁ ନାହିଁ. ଦେଇଥିବା ସମସ୍ତ ଉତ୍ତରକୁ ବାଛନ୍ତୁ। | ☐ 1. Elderly/Over 60 ବୃଦ୍ଧ/60 ରୁ ଊର୍ଦ୍ଧ୍ୱ<br>☐ 2. Men ପୁରୁଷ<br>☐ 3. Women ମହିଳା<br>☐ 4. Babies (<1 year) ଛୁଆ (<୧ ବର୍ଷ)<br>☐ 5. Children ପିଲା<br>☐ 6. Pregnant women ଗର୍ଭବତୀ ମହିଳା<br>☐ 7. People who are already sick/weak immune systems ଯେଉଁ ଲୋକମାନେ ପୂର୍ବରୁ ଅସୁସ୍ଥ/ ଯାହାର ରୋଗପ୍ରତିରୋଧକ ପ୍ରଣାଳୀ ଦୁର୍ବଳ ଅଟେ<br>☐ 8. Everyone ସମସ୍ତଙ୍କୁ<br>☐ 88. Other ଅନ୍ୟାନ୍ୟ: ________<br>☐ 99. Don't know ଜଣା ନାହିଁ |

### COVID-19 Mobile Phone Interview – Primary Caregiver

| | |
|---|---|
| **B6.** In the past 7 days, what actions have you and your family taken to avoid getting coronavirus? I am going to read a list of actions. For each one, please tell me yes or no.<br><br>ଗତ ସାତ ଦିନ ମଧରେ, ଆପଣ ଓ ଆପଣଙ୍କ ପରିବାରର ସଦସ୍ୟ ମାନେ ନିଜକୁ କରୋନାଭାଇରସରୁ ଦୂରେଇ ରଖିବାକୁ କଣ ପଦକ୍ଷେପ ନେଇଛନ୍ତି? ମୁଁ କିଛି ପଦକ୍ଷେପ ପଢୁଛି। ପ୍ରତିଟି ପଦକ୍ଷେପ ପାଇଁ ହଁ କିମ୍ବା ନାହିଁ କୁହନ୍ତୁ।<br><br>***Enumerator Notes:***<br>Select each action that the respondent says 'yes' for.<br><br>ଏନୁମରେଟର ନୋଟ: ସବୁ ପଦକ୍ଷେପ ବାଛନ୍ତୁ, ଯାହାପାଇଁ ଉତ୍ତରଦାତା 'ହଁ କହିଛନ୍ତି । | ☐ 1. Stayed at home as much as possible<br>ଯେତେ ପାରିଛନ୍ତି, ଘରେ ରହିଛନ୍ତି<br>☐ 2. Did not attend school or work<br>ସ୍କୁଲ କିମ୍ବା କାମକୁ ଯାଇନାହାନ୍ତି<br>☐ 3. Did not attend social gatherings (e.g., weddings, funerals, church, temple)<br>ସାମାଜିକ ସମାବେଶକୁ ଯାଇନାହାନ୍ତି (ଉଦାହରଣ : ବିବାହ,ଅନ୍ତିମ ସଂସ୍କାର, ଗୀର୍ଜାଘର, ମନ୍ଦିର)<br>☐ 4. Kept a distance from others → What distance? ____ meters<br>ଅନ୍ୟ ମାନଙ୍କ ଠାରୁ ଦୂରତ୍ୱ ରକ୍ଷା କରିଛନ୍ତି – କେତେ ମିଟର ଦୁରତ୍ୱ<br>☐ 6. Washed hands/used hand sanitizer more frequently<br>ହାତ ଧୋଇବା / ସାନିଟାଇଜର ର ବ୍ୟବହାର ବଢ଼େଇ ଦେଇଛନ୍ତି<br>☐ 7. Wore a (face) mask<br>ମାସ୍କ ପିନ୍ଧିଛନ୍ତି<br>☐ 8. Wore gloves (on your hands)<br>ଗ୍ଲୋଭ୍ସ ପିନ୍ଧିଛନ୍ତି<br>☐ 9. Tried to stop touching face<br>ମୁହଁଟିକୁ ନ ଛୁଇଁବାର ଚେଷ୍ଟା କରିଛନ୍ତି<br>☐ 10. Did not shake hands with others<br>ଅନ୍ୟମାନଙ୍କ ସହିତ ହାତ ମିଶେଇ ନାହାନ୍ତି<br>☐ 11. Covered your mouth with elbow when you sneezed or coughed<br>ଛିଙ୍କିବା କିମ୍ବା କାଶିବା ସମୟରେ ପାଟିକୁ ବାହୁରେ ଘୋଡାଇ ଦେଇଛନ୍ତି<br>☐ 12. Drank local alcohol ଦେଶୀ ମଦ ପିଇଛନ୍ତି<br>☐ 13. Scrubbed/cleaned surfaces such as door handles, faucets, phone, etc.<br>କୌଣସି ପୃଷ୍ଠକୁ ଘସିଛନ୍ତି/ସଫା କରିଛନ୍ତି ଯେପରିକି ଡୋର ହାଣ୍ଡେଲ, ଟ୍ୟାପ, ଫୋନ ଇତ୍ୟାଦି<br>☐ 14. Informed people of illness symptoms<br>ଲୋକ ମାନଙ୍କୁ ରୋଗର ଲକ୍ଷ୍ୟଣ ବିଷୟରେ ଜଣେଇଛନ୍ତି<br>☐ 15. Contacted a coronavirus helpline<br>କୌଣସି କରୋନାଭାଇରସ ହେଲ୍ପଲାଇନ କୁ ଯୋଗାଯୋଗ କରିଛନ୍ତି<br>☐ 16. Avoided hospitals/clinics<br>ହସ୍ପିଟାଲ କିମ୍ବା କ୍ଲିନିକକୁ ଏଡେଇଛନ୍ତି<br>☐ 17. Avoided public transit/traveling<br>ସର୍ବ ସାଧାରଣ ପରିବହନ/ଭ୍ରମଣକୁ ଏଡେଇଛନ୍ତି<br>☐ 18. Nothing କିଛି ନୁହେଁ<br>☐ 88. Other ଅନ୍ୟାନ୍ୟ: ________ |
| **B7.** What are some of the challenges you and your family face in being able to frequently wash your hands with water and soap?<br><br>ସାବୁନ ଓ ପାଣିରେ ବାରମ୍ବାର ହାତ ଧୋଇବାକୁ ନେଇ ଆପଣ ଓ ଆପଣଙ୍କ ପରିବାର ସଦସ୍ୟ କେଉଁ ଅସୁବିଧାର ସମ୍ମୁଖୀନ ହୁଅନ୍ତି? | ***Follow-up:***<br>The last time you needed to purchase soap for your household, did you experience any difficulties with getting the soap? Please explain.<br><br>ଶେଷଥର ପାଇଁ, ଯେବେ ଆପଣ ନିଜ ଘର ପାଇଁ ସାବୁନ କିଣିବାକୁ ଚାହିଁଲେ, କଣ ଆପଣଙ୍କୁ ସାବୁନ ମିଳିବାରେ କିଛି ଅସୁବିଧା ଅନୁଭବ ହେଲା କି? ଦୟାକରି ବର୍ଣ୍ଣନା କରନ୍ତୁ। |

### COVID-19 Mobile Phone Interview – Primary Caregiver

#### SECTION C: COVID-19 Impact on Daily Life and Child Feces Management Practices

**ENUMERATOR READ:** Now I am going to ask you some questions about whether or not life is different because of coronavirus or the lockdown. I'm going to ask you some questions about how your daily life has changed and how your childcare practices have changed, if changed. So for these questions, you want to think about what conditions were like ***BEFORE*** coronavirus and the lockdown and compare that to what things are like ***NOW***.

ଏନୁମରେଟର ପଢନ୍ତୁ : ବର୍ତ୍ତମାନ ମୁଁ ଆପଣଙ୍କୁ କିଛି ପ୍ରଶ୍ନ, କଣ ଆପଣଙ୍କ ଜୀବନ କାରୋନାଭାଇରସ କିମ୍ବା ଲକଡ଼ାଉନ ପାଇଁ ଅଲଗା କି ନୁହେଁ ପଚାରିବି। ମୁଁ ଆପଣଙ୍କୁ କିଛି ପ୍ରଶ୍ନ ଆପଣଙ୍କ ଦୈନିକ ଜୀବନ କିପରି ବଦଳିଛି ଏବଂ ଆପଣଙ୍କ ଶିଶୁଯତ୍ନ ଅଭ୍ୟାସ କିପରି ବଦଳିଛି, ଯଦି ବଦଳିଛି, ସେ ଉପରେ ପଚାରିବି। ଏହି ପ୍ରଶ୍ନମାନଙ୍କ ପାଇଁ, ଆପଣଙ୍କୁ ଭାବିବାକୁ ପଡିବ ଯେ କରୋନାଭାଇରସ କିମ୍ବା ଲକଡାଉନ ପୂର୍ବରୁ ସ୍ଥିତି କିପରି ଥିଲା ଏବଂ ଏହାକୁ ବର୍ତ୍ତମାନର ସ୍ଥିତି ସହିତ ତୁଳନା କରନ୍ତୁ।

**C1.** How has coronavirus or the lockdown changed your daily life? Think about from the time you get up in the morning to the time you go to sleep at night. How is your day different now, if it is different?

କରୋନାଭାଇରସ କିମ୍ବା ଲକଡାଉନ ଆପଣଙ୍କ ଦୈନିକ ଜୀବନକୁ କିପରି ବଦଳେଇ ଦେଇଛି? ସକାଳୁ ଉଠିବା ସମୟରୁ ଆରମ୍ଭ କରି ରାତିରେ ଶୋଇବାକୁ ଯିବା ପର୍ଯ୍ୟନ୍ତ ସମୟ ବିଷୟରେ ଭାବନ୍ତୁ। ବର୍ତ୍ତମାନ ଆପଣଙ୍କ ଦିନ ଅଲଗା କିପରି, ଯଦି ଅଲଗା ହୋଇଥାଏ?

***Probes:*** *ଅନୁସନ୍ଧାନ:*

- Staying at home more? ଘରେ ଅଧିକ ସମୟ ରହୁଛନ୍ତି?
- More/less workload? ଅଧିକ/କମ କାମ
- More/less free time? ଅଧିକ/କମ ଖାଲି ସମୟ
- Is there anywhere in the village you *used* to go but no longer go? ଗାଁରେ ଏପରି କିଛି ଜାଗା ଅଛି କି ଯେଉଠିକି ଆପଣ ଯାଉଥିଲେ କିନ୍ତୁ ଆଉ ଯାଉ ନାହାନ୍ତି?

**C2.** Has coronavirus or the lockdown changed how you or your family **interact with other members of your village**? Please explain.

କଣ କରୋନାଭାଇରସ କିମ୍ବା ଲକଡାଉନ ଆପଣଙ୍କର କିମ୍ବା ଆପଣଙ୍କ ପରିବାରର ଗ୍ରାମର ଅନ୍ୟ ଲୋକମାନଙ୍କ ସହିତ ମିଳାମିଶାରେ କିଛି ପରିବର୍ତ୍ତନ ଆଣିଛି? ଦୟାକରି ବର୍ଣ୍ଣନା କରନ୍ତୁ।

- ☐ 1. Yes ହଁ
- ☐ 2. No ନାହିଁ

**Please explain. ଦୟାକରି ବର୍ଣ୍ଣନା କରନ୍ତୁ।**

**ENUMERATOR READ:** Now I am going to ask you some questions about your youngest child. Please think about this child.

ଏନୁମରେଟର ପଢନ୍ତୁ : ବର୍ତ୍ତମାନ ମୁଁ ଆପଣଙ୍କୁ ଆପଣଙ୍କ ସବୁଠୁ ଛୋଟ ପିଲା ବିଷୟରେ କିଛି ପ୍ରଶ୍ନ ପଚାରିବାକୁ ଯାଉଛି। ଏହି ପିଲାଟି ବିଷୟରେ ଭାବନ୍ତୁ।

**ENUMERATOR CONFIRM:** Please record again the age of the youngest child <5 years old, who is being discussed:
ଏନୁମରେଟର ପୁଷ୍ଟି କରନ୍ତୁ: ଦୟାକରି, 5 ବର୍ଷରୁ କମ ସବୁଠୁ ଛୋଟ ପିଲା, ଯାହା ବିଷୟରେ ଚର୍ଚ୍ଚା କରାଯାଉଛି, ତା'ର ବୟସ ଟିପନ୍ତୁ।

Years: ବର୍ଷ ________ Months: ମାସ_________

<table>
<tr>
<td>C3. How has coronavirus or the lockdown changed your child’s daily life? Think about from the time your child wakes up in the morning to when they go to bed at night. How is your child’s day different now, if at all?<br><br>କରୋନାଭାଇରସ କିମ୍ବା ଲକଡାଉନ ଆପଣଙ୍କ ପିଲାର ଦୈନିକ ଜୀବନକୁ କିପରି ବଦଳେଇ ଦେଇଛି? ସକାଳୁ ଉଠିବା ସମୟରୁ ଆରମ୍ଭ କରି ରାତିରେ ଶୋଇବାକୁ ଯିବା ପର୍ଯ୍ୟନ୍ତ ସମୟ ବିଷୟରେ ଭାବନ୍ତୁ। ବର୍ତ୍ତମାନ ଆପଣଙ୍କ ପିଲାର ଦିନ ଅଲଗା କିପରି, ଯଦି ଅଲଗା ହୋଇଥାଏ?<br><br>Probes:<br>○ Staying at home more? ଘରେ ଅଧିକ ସମୟ ରହୁଛି?<br>○ Able to go to Anganwadi? ଅଙ୍ଗନବାଡି ଯାଇ ପାରୁଛି?<br>○ Roaming around more/less? ଅଧିକ/କମ ବୁଲୁଛି?</td>
<td></td>
</tr>
<tr>
<td>C4. Who typically helps you with caring for your child? For example, helps watch the child while you do something else or helps with things like bathing, feeding, dressing, and managing the child’s feces.<br><br>ସାଧାରଣତଃ, ଆପଣଙ୍କୁ ନିଜ ପିଲାର ଯତ୍ନ ନେବାରେ କିଏ ସାହାଯ୍ୟ କରନ୍ତି? ଉଦାହରଣ ସ୍ୱରୂପ, ଆପଣ କିଛି କାମ କଲାବେଳେ ପିଲାକୁ ରଖନ୍ତି କିମ୍ବା ପିଲାକୁ ଗାଧୋଇବାରେ, ଖୁଏଇବାରେ, କପଡା ପିନ୍ଧେଇବାରେ ଏବଂ ପିଲାର ଝାଡା ପରିଚାଳନାରେ ସାହାଯ୍ୟ।</td>
<td>Follow-up #1:<br>How has coronavirus or the lockdown changed the help you get with childcare?<br>କରୋନାଭାଇରସ କିମ୍ବା ଲକଡାଉନ ଆପଣଙ୍କ ପିଲା ଯତ୍ନ ସହ ଜଡିତ ମିଳୁଥିବା ସାହାଯ୍ୟକୁ କିପରି ପରିବର୍ତ୍ତିତ କରିଛି?<br><br>○ Getting more, less, or same amount of help?<br>କମ, ସମାନ ନା ଅଧିକ ପରିମାଣର ସାହାଯ୍ୟ ମିଳୁଛି?<br>○ Men in the household help more?<br>ଘରର ପୁରୁଷମାନେ ଅଧିକ ସାହାଯ୍ୟ କରୁଛନ୍ତି?<br>○ Neighbors are no longer able to help?<br>ପଡୋଶୀ ମାନେ ଆଉ ସାହାଯ୍ୟ କରିପାରୁ ନାହାନ୍ତି?<br><br>Follow-up #2:<br>Due to coronavirus or the lockdown, has there been any change in who helps with managing the child’s feces?<br>କରୋନାଭାଇରସ କିମ୍ବା ଲକଡ଼ାଉନ ପାଇଁ, କଣ ପିଲା ଝାଡା ପରିଚାଳନରେ କିଏ ସାହାଯ୍ୟ କରେ, ସେଥିରେ କିଛି ପରିବର୍ତ୍ତନ ଆସିଛି କି?</td>
</tr>
</table>

| | |
|---|---|
| **C5.** The last time your youngest child defecated, where did they defecate?<br><br>ଶେଷ ଥର ପାଇଁ ଆପଣଙ୍କ ସବୁଠୁ ସାନ ପିଲା କେଉଁଠାରେ ଝାଡା କରିଥିଲା? | ☐ 1. Inside the household ଘର ଭିତରେ<br>☐ 2. Inside household compound ଘର ଅଗଣା ଭିତରେ<br>☐ 3. Just outside the household compound ଘର ଅଗଣାରୁ ଟିକେ ବାହାରେ<br>☐ 4. Away from the household compound ଘର ଅଗଣା ବାହାରେ<br>☐ 5. In latrine ପାଇଖାନା ରେ → **SKIP to C6**<br>☐ 6. In latrine but not over pan ପାଇଖାନା ଘର ରେ କିନ୍ତୁ ପ୍ୟାନ ରେ ନୁହେଁ<br>☐ 88. Other ଅନ୍ୟାନ୍ୟ ________<br>☐ 99. Don't know ଜାଣି ନାହିଁ |
| **C5a.** The last time your youngest child defecated, what did they defecate on?<br><br>ଶେଷ ଥର ପାଇଁ ଆପଣଙ୍କ ସବୁଠୁ ଛୋଟ ପିଲା ଯେବେ ଝାଡା କରିଥିଲା, ସେ କେଉଁଥିରେ ଝାଡା କରିଥିଲେ ? | ☐ 1. On ground directly (soil) ସିଧା ତଳେ (ମାଟିରେ )<br>☐ 2. On ground directly (cement) ସିଧା ତଳେ (ସିମେଣ୍ଟରେ )<br>☐ 3. On cloth on ground or other surface କପଡ଼ାରେ ତଳେ ବା ଚଟାଣ ରେ<br>☐ 4. On cloth on bed ଖଟ ଉପରେ କପଡା ରେ<br>☐ 5. On oil cloth on ground or other surface ତଳେ ବା ଚଟାଣ ରେ ଅଇଲ କଲଥ ଉପରେ<br>☐ 6. On oil cloth on bed ଖଟ ଉପରେ ଅଇଲ କଲଥ ଉପରେ<br>☐ 7. On waste newspaper/paper ପୁରୁଣା କାଗଜ/ଖାଲି କାଗଜ<br>☐ 8. On polythene bag ପଲିଥିନ୍ ଜରି/ବ୍ୟାଗ<br>☐ 9. On caregiver/person holding the child ପିଲାକୁ ଯିଏ ଧରି ଥାଏ ତା ଉପରେ<br>☐ 10. In cloth nappy କପଡା ନାପକିନ୍<br>☐ 11. In disposable diaper (Huggies, Pampers) ଡାଇପର୍ ରେ<br>☐ 12. In pants/clothing ପ୍ୟାଣ୍ଟ/ ପିନ୍ଧିଥିବା ଡ୍ରେସ୍ ରେ<br>☐ 13. On dustpan ଡଷ୍ଟବିନ ରେ<br>☐ 14. In potty ପୋଟ୍ଟି ରେ<br>☐ 88. Other ଅନ୍ୟାନ୍ୟ ________<br>☐ 99. Don't know ଜାଣି ନାହିଁ |
| **C5b.** This last time your youngest child defecated, where were their feces disposed?<br><br>ଶେଷ ଥର ପାଇଁ ଆପଣଙ୍କ ସବୁଠୁ ସାନ ପିଲା କରିଥିବା ଝାଡାକୁ କେଉଁଠାରେ ଫୋପଡା ଯାଇଥିଲା? | ☐ 1. Into latrine ପାଇଖାନା ରେ<br>☐ 2. Buried ପୋତି ଦିଆଯାଇଥିଲା<br>☐ 3. Into backyard of household compound ଘର ଅଗଣା ପଛପଟେ<br>☐ 4. Into household garbage pile ଘର ଆବର୍ଜନା ଜାଗାରେ<br>☐ 5. Into household compost pile ଘର କମ୍ପୋଷ୍ଟ ରେ<br>☐ 6. Into open field (NOT on household compound) ଖୋଲା ପଡିଆ ରେ (ଘର ଅଗଣା ନୁହେଁ )<br>☐ 7. Into community garbage pile ଗୋଷ୍ଠି ଆବର୍ଜନା ଜାଗାରେ<br>☐ 8. Into community compost pile ଗୋଷ୍ଠି କମ୍ପୋଷ୍ଟ ରେ<br>☐ 9. Along roadside ରୋଡ଼ ସାଇଡ ରେ<br>☐ 10. Into drain/ditch ନଳା ବା ଗାତ ରେ<br>☐ 11. Washed directly in bathroom ବାଥ ରୁମ ରେ ସିଧା ଧୋଇ ଦେଲି<br>☐ 12. Washed directly in body of water ନଦୀ,ନଳା ରେ ସିଧା ଧୋଇ ଦେଲି<br>☐ 13. Left in open (in household compound) ଖୋଲା ରେ ଛାଡ଼ି ଦିଆଯାଇଥିଲା (ଘର ପରିସର ମଧ୍ୟରେ)<br>☐ 88. Other ଅନ୍ୟାନ୍ୟ: ________ |

| | |
|---|---|
| | ☐ 99. Don't know ଜାଣି ନାହିଁ |
| **C5c.** How strongly do you intend to ALWAYS dispose of your child's feces into the latrine?<br><br>ଆପଣ କେତେ ସୁଦୃଢ ଭାବେ ଉଦ୍ଦେଶ୍ୟ ରଖିଛନ୍ତି କି ଯେ ଆପଣଙ୍କ ପିଲାର ମଳ ଆପଣ ସବୁବେଳେ ପାଇଖାନାରେ ପକାଇବେ?<br><br>***Enumerator Notes:***<br>Read the response options<br>ଉତ୍ତର ଅପସନସ୍ ଗୁଡିକୁ ପଢନ୍ତୁ । | ☐ 1. Not at all ବିଲକୁଲ ନୁହଁ<br>☐ 2. A little ଅଳ୍ପ<br>☐ 3. Medium ମଧ୍ୟମ<br>☐ 4. Strongly ସୁଦୃଢ<br>☐ 5. Very strongly ବହୁତ ସୁଦୃଢ |
| **C6.** Is the location where your child defecates *now* the same or different from where your child defecated *before* coronavirus or the lockdown?<br><br>କଣ ଆପଣଙ୍କ ପିଲାର ବର୍ତ୍ତମାନର ଝାଡା କରୁଥିବା ଜାଗା ; ଆପଣଙ୍କ ପିଲାର କରୋନାଭାଇରସ କିମ୍ବା ଲକଡାଉନ ପୂର୍ବରୁ ଝାଡା କରୁଥିବା ଜାଗା ସହିତ ସମାନ ନା ଅଲଗା? | ☐ 1. Same ସମାନ<br>☐ 2. Different → How has the location where your child defecates changed? Why has it changed? Please explain.<br>ଅଲଗା → ଆପଣଙ୍କ ପିଲା ଝାଡା କରୁଥିବା ଜାଗାଟି କିପରି ପରିବର୍ତିତ ହେଲା? ଦୟାକରି, ବର୍ଣ୍ଣନା କରନ୍ତୁ। |
| **C7.** Is the location where you dispose of your child feces *now* the same or different from where you disposed of your child's feces *before* coronavirus or the lockdown?<br><br>କଣ ସେହି ଜାଗା ଯେଉଁଠାରେ ଆପଣ ନିଜ ପିଲାର ଝାଡାକୁ ବର୍ତ୍ତମାନ ଫୋପାଡନ୍ତି , ଆପଣ ପୂର୍ବରୁ ନିଜ ପିଲାର ଝାଡାକୁ ଫୋପାଡୁଥିବା ଜାଗା ସହିତ ସମାନ ନା ଅଲଗା? | ☐ 1. Same ସମାନ<br>☐ 2. Different → How has the location where you dispose of your child's feces changed? Why has it changed? Please explain.<br>ଅଲଗା → ଆପଣଙ୍କ ପିଲାର ଝାଡାକୁ ଫୋପାଡୁଥିବା ଜାଗାଟି କିପରି ପରିବର୍ତ୍ତିତ ହେଲା? ଏହା କାହିଁ ପରିବର୍ତ୍ତିତ ହେଲା? ଦୟାକରି ବର୍ଣ୍ଣନା କରନ୍ତୁ। |
| **C8. *ONLY ASK** if caregiver uses cloth/nappies: Tell me about how you wash your child's soiled cloths/nappies. How has this changed because of coronavirus or the lockdown, if it has changed?<br><br>**କେବଳ ସେବେ ପଚାରନ୍ତୁ** ଯେବେ ଉତ୍ତରଦାତା କପଡା/କନା ବ୍ୟବହାର କରନ୍ତି: ଆପଣ ନିଜ ପିଲାର ଓଦା କପଡା/ କନା କିପରି ସଫା କରନ୍ତି? ଏହା କରୋନାଭାଇରସ କିମ୍ବା ଲକଡ଼ାଉନ ପାଇଁ କିପରି ବଦଳିଛି, ଯଦି ବଦଳିଛି?<br><br>***Probes:***<br>○ Change in type or amount of soap you use?<br>ଆପଣ ବ୍ୟବହାର କରୁଥିବା ସାବୁନରେ ପରିବର୍ତ୍ତନ?<br>○ Change in amount of water? |  |

| | |
|---|---|
| ଆପଣ ବ୍ୟବହାର କରୁଥିବା ପାଣିର ପରିମାଣରେ ପରିବର୍ତ୍ତନ?<br>○ Change in *where* you go to wash?<br>ଆପଣ ଧୋଇବାକୁ ଯାଉଥିବା ଜାଗାରେ ପରିବର୍ତ୍ତନ? | |
| **C9.** Are you currently teaching your youngest child how to use the latrine for defecation?<br><br>କଣ ଆପଣ ବର୍ତ୍ତମାନ ଆପଣଙ୍କ ସବୁଠୁ ଛୋଟ ପିଲାକୁ ଝାଡା ଯିବା ପାଇଁ କିପରି ପାଇଖାନାକୁ ଯିବେ ଶିଖାଉଛନ୍ତି କି ? | ☐ 1. Yes – ହଁ → **Ask C9a and C10** and complete **latrine training** questions below<br>☐ 2. No - my child is not old enough to start learning how to use the latrine ନା- ମୋ ପିଲା ସେତିକି ବଡ ହୋଇ ନାହିଁ କି ପାଇଖାନା ବ୍ୟବହାର ଶିକ୍ଷା ଆରମ୍ଭ କରିବ → **Skip to C11** and complete **disposal** questions below<br>☐ 3. No – my child already knows how to use the latrine on their own ନାହିଁ- ମୋ ପିଲା ନିଜେ ପାଇଖାନା ବ୍ୟବହାର କରି ଜାଣିଚି (ପାଣି ଢାଳି ଓ ଛାଞ୍ଚେଇବା ମଧ୍ୟ ଜାଣିଛି) → **Skip to C15** (**skip** disposal and latrine training quesitons)<br>☐ 4. No – I am not teaching my child how to use the latrine though my child is old enough ନା- ମୁଁ ମୋ ପିଲା କୁ ପାଇଖାନା ବ୍ୟବହାରର ଶିକ୍ଷା ଦେବା ଆରମ୍ଭ କରିନାହିଁ ଯଦିଓ ମୋ ପିଲାର ବୟସ ହୋଇଯାଇଛି → **Ask C10** and complete **latrine training** questions below |
| **C9a.**<br>**ONLY ASK if C9 = 1**<br>During the last week, when your child needed to defecate, how often did you take your child to the latrine and teach them how to use it?<br><br>ଗତ ସପ୍ତାହରେ, ଯେତେବେଳେ ଆପଣଙ୍କ ପିଲା ଝାଡା ଯିବାକୁ ଇଚ୍ଛା କରିଥିଲା, ଆପଣ ତାକୁ କେତେଥର ପାଇଖାନାକୁ ନେଇ ଯାଇଥିଲେ ଏବଂ ଏହାକୁ ବ୍ୟବହାର କରିବାକୁ ଶିଖେଇଥିଲେ?<br><br>***Enumerator Notes:***<br>Read the response options<br>ଉତ୍ତର ଅପସନସ୍ ଗୁଡିକୁ ପଢନ୍ତୁ । | ☐ 1. (Almost) never (0%) କେବେ ନୁହେ(୦%)<br>☐ 2. Seldom (25%) କେବେ କେବେ (୦%)<br>☐ 3. Sometimes (50%) କେବେ କେବେ (୫୦%)<br>☐ 4. Often (75%) ପ୍ରାୟତଃ (75%)<br>☐ 5. (Almost) always (100%) ସବୁବେଳେ (100%) |
| **C10.** Were you teaching your youngest child how to use the latrine for defecation *before* coronavirus and the lockdown?<br><br>କରୋନାଭାଇରସ କିମ୍ବା ଲକଡାଉନ ପୂର୍ବରୁ କଣ ଆପଣ ନିଜର ସବୁଠୁ ଛୋଟ ପିଲାକୁ ଝାଡା ଯିବା ପାଇଁ ପାଇଖାନା ବ୍ୟବହାର କରିବା ଶିଖାଉଥିଲେ କି? | ☐ 1. Yes ହଁ<br>☐ 2. No ନାହିଁ<br><br>**If there is a change, ask:** ଯଦି କିଛି ପରିବର୍ତ୍ତନ ଅଛି, ତାହେଲେ ପଚାରନ୍ତୁ :<br>Why did you [*start* / stop] teaching your child how to use the latrine?<br>ଆପଣ ନିଜ ପିଲାକୁ କାହିଁକି ପାଇଖାନା ବ୍ୟବହାର ବିଷୟରେ ଶିଖେଇବା ଆରମ୍ଭ କିମ୍ବା ବନ୍ଦ କରିଦେଲେ? |

### COVID-19 Mobile Phone Interview – Primary Caregiver

**ENUMERATOR NOTE:** Make sure you complete the correct set of either ‘disposal’ or ‘latrine training’ questions based on the respondent’s answer to C16 above.
ଏନୁମରେଟର ନୋଟ : ଏହା ନିଶ୍ଚିନ୍ତ କରନ୍ତୁ କି ଯେ ଆପଣ ଉତ୍ତରଦାତାଙ୍କ C16 ପ୍ରଶ୍ନର ଉତ୍ତରର ଆଧାରରେ ହିଁ ‘ନିଷ୍କାସନ’ କିମ୍ବା ‘ପାଇଖାନା ପ୍ରଶିକ୍ଷଣ’ ଭାଗର ପ୍ରଶ୍ନକୁ ସମ୍ପୂର୍ଣ କରିବେ।

| DISPOSAL | LATRINE TRAINING |
|---|---|
| **C11. RISKS**<br>○ Do you think a baby’s feces (<1 year old) can make someone sick? Why or why not?<br>ଆପଣଙ୍କୁ ଲାଗୁଛି କି ଯେ ଏକ ଶିଶୁର ଝାଡା (ଏକ ବର୍ଷରୁ କମ) କାହାକୁ ଅସୁସ୍ଥ କରିପାରେ? କାହିଁକି ଏବଂ କାହିଁକି ନୁହେଁ?<br><br>○ Do you think a child’s feces (1 to <5 year old) can make someone sick? Why or why not?<br>ଆପଣଙ୍କୁ ଲାଗୁଛି କି ଯେ ଏକ ପିଲାର ଝାଡା (ଏକ ବର୍ଷରୁ ଅଧିକ ଓ ପାଞ୍ଚ ବର୍ଷରୁ କମ) କାହାକୁ ଅସୁସ୍ଥ କରିପାରେ? କାହିଁକି ଏବଂ କାହିଁକି ନୁହେଁ? | **C11. RISKS**<br>○ What concerns do you have when your child uses the latrine for defecation?<br>ଯେତେବେଳେ ଆପଣଙ୍କ ପିଲା ଝାଡା ଯିବା ପାଇଁ ପାଇଖାନା ବ୍ୟବହାର କରେ,ଆପଣଙ୍କର କଣ ଆଶଙ୍କା ଥାଏ?<br><br>○ What concerns do you have when your child goes out for defecation?<br>ଯେତେବେଳେ ଆପଣଙ୍କ ପିଲା ଝାଡା ଯିବା ପାଇଁ ବାହାରକୁ ଯାଏ,ଆପଣଙ୍କର କଣ ଆଶଙ୍କା ଥାଏ? |
| **C12. ATTITUDES**<br>○ What are the **good things** about managing your child’s feces?<br>ଆପଣଙ୍କୁ ନିଜ ପିଲାର ଝାଡା ପରିଚାଳନରେ କଣ ଭଲ ଲାଗେ?<br><br>○ What are the **bad things**?<br>କଣ ଖରାପ ଲାଗେ? | **C12. ATTITUDES**<br>○ What are the **good things** about teaching your child how to use the latrine for defecation?<br>ଆପଣଙ୍କୁ ନିଜ ପିଲାକୁ ପାଇଖାନାକୁ ଝାଡା ଯିବାପାଇଁ ବ୍ୟବହାର କରିବାରେ ଶିକ୍ଷା ଦେବାରେ କଣ ଭଲ ଲାଗେ?<br><br>○ What are the **bad things**?<br>କଣ ଖରାପ ଲାଗେ? |
| **C13. ABILITY**<br>Due to coronavirus or the lockdown, what challenges are you facing with managing your child’s feces? Any challenges with disposal?<br><br>କରୋନାଭାଇରସ କିମ୍ବା ଲକଡ଼ାଉନ ପାଇଁ, ଆପଣ ନିଜ ପିଲାର ଝାଡାକୁ ପରିଚାଳନା କରିବାରେ କେଉଁ କେଉଁ ସମସ୍ୟାର ସାମ୍ନା କରନ୍ତି? ଫୋପାଡିବାକୁ ନେଇ କିଛି ସମସ୍ୟା? | **C13. ABILITY**<br>Due to coronavirus or the lockdown, what challenges are you facing with teaching your child how to use the latrine for defecation?<br><br>କରୋନାଭାଇରସ କିମ୍ବା ଲକଡ଼ାଉନ ପାଇଁ ଆପଣ ନିଜ ପିଲାକୁ ପାଇଖାନାକୁ ଝାଡା ଯିବା ପାଇଁ ବ୍ୟବହାର କରିବାର ଶିକ୍ଷା ଦେବାରେ କେଉଁ ସମସ୍ୟାର ସାମ୍ନା କରନ୍ତି? |

| | |
|---|---|
| ***Probes***:<br>-Less help with caring for child?<br>ପିଲାର ଯତ୍ନରେ କମ ସାହାଯ୍ୟ?<br>-More workload now? ବର୍ତ୍ତମାନ ଅଧିକ କାମ? | ***Probes***:<br>-Less help with teaching child? ପିଲାକୁ ଶିଖେଇବାରେ କମ ସାହାଯ୍ୟ?<br>-Child has become more difficult? ପିଲାଟି ଦୁଷ୍ଟ ହୋଇଯାଇଛି କି?<br>-More workload now? ବର୍ତ୍ତମାନ ଅଧିକ କାମ? |
| **C14a. NORMS**<br>Due to coronavirus or the lockdown, do you think people in this village are disposing of their child's feces into the latrine more, less, or there has been no change?<br><br>କରୋନାଭାଇରସ କିମ୍ବା ଲକଡାଉନ ପାଇଁ, ଆପଣ ଭାବୁଛନ୍ତି କି ଯେ ଏଇ ଗାଁ ର ଲୋକ ମାନେ ନିଜ ପିଲାର ଝାଡାକୁ ଲାଟ୍ରିନରେ ଅଧିକ ଫୋପାଡୁଛନ୍ତି, କମ କିମ୍ବା କିଛି ପରିବର୍ତ୍ତନ ହୋଇନାହିଁ?<br>☐ 1. More ଅଧିକ<br>☐ 2. Less କମ<br>☐ 3. No change କିଛି ପରିବର୍ତ୍ତନ ନୁହେଁ<br>☐ 99. Don't know ଜାଣିନାହିଁ<br><br>**Please explain why: ଦୟାକରି ବର୍ଣ୍ଣନା କରନ୍ତୁ କାହିଁକି** | **C14a. NORMS**<br>Due to coronavirus or the lockdown, do you think people in this village are teaching their children to defecate in the latrine more, less, or there has been no change?<br><br>କରୋନାଭାଇରସ କିମ୍ବା ଲକଡାଉନ ପାଇଁ, ଆପଣ ଭାବୁଛନ୍ତି କି ଯେ ଏଇ ଗାଁ ର ଲୋକ ମାନେ ନିଜ ପିଲାକୁ ପାଇଖାନାରେ ଝାଡା ଯିବା ପାଇଁ ଅଧିକ ଶିଖାଉଛନ୍ତି, କମ କିମ୍ବା କିଛି ପରିବର୍ତ୍ତନ ହୋଇନାହିଁ?<br>☐ 1. More ଅଧିକ<br>☐ 2. Less କମ<br>☐ 3. No change କିଛି ପରିବର୍ତ୍ତନ ନୁହେଁ<br>☐ 99. Don't know ଜାଣିନାହିଁ<br><br>**Please explain why: ଦୟାକରି ବର୍ଣ୍ଣନା କରନ୍ତୁ କାହିଁକି** |
| **C14b. NORMS**<br>Due to coronavirus or the lockdown, do your household members now expect you to dispose of your child's feces into the latrine? Why or why not?<br><br>କରୋନାଭାଇରସ କିମ୍ବା ଲକଡାଉନ ପାଇଁ କଣ ଆପଣଙ୍କ ଘରର ସଦସ୍ୟମାନେ ଆପଣଙ୍କ ଠାରୁ ଏହି ଆଶା କରୁଛନ୍ତି କି ଯେ ବର୍ତ୍ତମାନ ଆପଣ ନିଜ ପିଲାର ଝାଡାକୁ ଲାଟ୍ରିନରେ ଫୋପାଡନ୍ତୁ? କାହିଁକି ଏବଂ କାହିଁକି ନୁହେଁ? | **C14b. NORMS**<br>Due to coronavirus or the lockdown, do your household members expect you to now teach your child how to use the latrine for defecation? Why or why not?<br><br>କରୋନାଭାଇରସ କିମ୍ବା ଲକଡାଉନ ପାଇଁ, କଣ ଆପଣଙ୍କ ଘରର ସଦସ୍ୟମାନେ ଆପଣଙ୍କ ଠାରୁ ଏହି ଆଶା କରୁଛନ୍ତି କି ଯେ ଆପଣ ବର୍ତ୍ତମାନ ନିଜ ପିଲାକୁ ପାଇଖାନାକୁ ଝାଡା ଯିବା ପାଇଁ ବ୍ୟବହାର କରିବାର ଶିକ୍ଷା ଦିଅନ୍ତୁ? କାହିଁକି ଏବଂ କାହିଁକି ନୁହେଁ? |

### COVID-19 Mobile Phone Interview – Primary Caregiver

| | |
|---|---|
| **ENUMERATOR READ:** Thank you for your responses so far. Now I'm going to ask you some questions about how coronavirus has impacted other aspects of your daily life.<br><br>**ଏନୁମରେଟର ପଢନ୍ତୁ**: ବର୍ତ୍ତମାନ ପର୍ଯ୍ୟନ୍ତ ଦେଇଥିବା ଆପଣଙ୍କ ଉତ୍ତର ପାଇଁ ଧନ୍ୟବାଦ। ମୁଁ ବର୍ତ୍ତମାନ କିଛି ପ୍ରଶ୍ନ କରୋନାଭାଇରସ ଆପଣଙ୍କ ଜୀବନର ଅନ୍ୟ ଦିଗଗୁଡିକୁ କିପରି ପ୍ରଭାବିତ କରିଛି, ସେ ବିଷୟରେ ପଚାରିବି। | |
| **C15.** The last time you defecated, did you defecate in the open or use the latrine?<br><br>କଣ ଶେଷ ଥର ଯେବେ ଆପଣ ଝାଡା ଯାଇଥିଲେ **,** ତେବେ ଆପଣ ଖୋଲା ରେ ଝାଡା ଯାଇଥିଲେ ନା ପାଇଖାନା ବ୍ୟବହାର କରିଥିଲେ**?** | ☐ 1. Open ଖୋଲା ରେ<br>☐ 2. Latrine ପାଇଖାନା ରେ<br>☐ 3. Somewhere else (not open field or latrine)<br>ଅଲଗା କେଉଁଠି (ଖୋଲା ପଡିଆ ନୁହେଁ କିମ୍ବା ପାଇଖାନା ନୁହେଁ)<br>☐ 99. Don't know ଜାଣିନାହିଁ |
| **C16.** Has coronavirus or the lockdown changed the **defecation location of you or any family members?** Please explain.<br><br>କଣ କରୋନାଭାଇରସ କିମ୍ବା ଲକଡ଼ାଉନ ଆପଣ କିମ୍ବା ଆପଣଙ୍କ କେହି ପରିବାର ସଦସ୍ୟଙ୍କ ଝାଡା ଯିବା ଜାଗାକୁ ବଦଳେଇଛି କି? ଯଦି ହଁ, ବର୍ଣ୍ଣନା କରନ୍ତୁ କିପରି।<br><br>***Probe:*** Have there been any changes for **elderly adults** in your household?<br><br>ଆପଣଙ୍କ ଘରର ବୃଦ୍ଧ ବ୍ୟକ୍ତିଙ୍କ କ୍ଷେତ୍ରରେ କିଛି ପରିବର୍ତ୍ତନ ଆସିଛି କି**?** | ☐ 1. Yes ହଁ<br>☐ 2. No ନାହିଁ<br><br>**Please explain. ବର୍ଣ୍ଣନା କରନ୍ତୁ** |
| **C17.** Due to coronavirus or the lockdown, do you think people in this village are defecating in the open more, defecating in a latrine more, or there has been no change in where people defecate? Explain why<br><br>ଆପଣ ଭାବୁଛନ୍ତି କି ଯେ, କରୋନାଭାଇରସ କିମ୍ବା ଲକଡ଼ାଉନ ପାଇଁ ଆପଣଙ୍କ ଗ୍ରାମର ଲୋକମାନେ, ଖୋଲାରେ ଅଧିକ ଝାଡା ଯାଉଛନ୍ତି, ଲାଟ୍ରିନର ରେ ଅଧିକ ଝାଡା ଯାଉଛନ୍ତି ନା ଲୋକ ଝାଡା କରୁଥିବା ଜାଗାରେ କୌଣସି ପରିବର୍ତ୍ତନ ଆସିନାହିଁ? ବର୍ଣ୍ଣନା କରନ୍ତୁ କଣ ପାଇଁ। | ☐ 1. More open defecation ଖୋଲରେ ଝାଡା ଯିବା ବଢିଛି<br>☐ 2. More latrine use ଲାଟ୍ରିନ ବ୍ୟବହାର ବଢିଛି<br>☐ 3. No change କିଛି ପରିବର୍ତ୍ତନ ନୁହେଁ<br>☐ 99. Don't know ଜାଣିନାହିଁ<br><br>**Explain why: ବର୍ଣ୍ଣନା କରନ୍ତୁ କାହିଁକି** |
| **C18. Has coronavirus or the lockdown changed your handwashing practices? Please explain.**<br><br>କଣ କରୋନାଭାଇରସ **କିମ୍ବା ଲକଡ଼ାଉନ ଆପଣଙ୍କ** ହାତଧୁଆ ପ୍ରକ୍ରିୟାକୁ ବଦଳେଇଛି**?** ଦୟାକରି ବର୍ଣ୍ଣନା କରନ୍ତୁ।<br><br>***Probes:***<br>**Has there been any change in how often you wash your hands?**<br>କଣ ଆପଣ ପ୍ରାୟତଃ କେତେଥର ହାତ ଧୁଅନ୍ତି**,** ସେଥିରେ କିଛି ପରିବର୍ତ୍ତନ ଆସିଛି କି **?** | ☐ 1. Yes ହଁ<br>☐ 2. No ନାହିଁ<br><br>**Please explain. ଦୟାକରି ବର୍ଣ୍ଣନା କରନ୍ତୁ** |

| | |
|---|---|
| **Has there been any change in how much soap or water you use to wash your hands?**<br>କଣ ଆପଣ ପ୍ରାୟତଃ କେତେ ପରିମାଣର ସାବୁନ କିମ୍ବା ପାଣି ବ୍ୟବହାର କରନ୍ତି, ସେଥିରେ କିଛି ପରିବର୍ତ୍ତନ ଆସିଛି କି? | |
| **C19.** Has coronavirus or the lockdown changed the **handwashing practices for any young children (<5 years) in your household**? Please explain.<br><br>କଣ କରୋନାଭାଇରସ କିମ୍ବା ଲକଡ଼ାଉନ ଆପଣଙ୍କ ଘରର **5 ବର୍ଷରୁ କମ ବୟସର କେହି ପିଲାର ହାତଧୁଆ ପ୍ରକ୍ରିୟାକୁ** ବଦଳେଇଛି କି? ଦୟାକରି ବର୍ଣ୍ଣନା କରନ୍ତୁ। | ☐ 1. Yes ହଁ<br>☐ 2. No ନାହିଁ<br><br>**Please explain. ଦୟାକରି ବର୍ଣ୍ଣନା କରନ୍ତୁ** |
| **C20. In the past 7 days, have you had any difficulties with getting the water that you need for your household? Please explain.**<br><br>କରୋନାଭାଇରସ କିମ୍ବା ଲକଡାଉନ ପାଇଁ ଗତ ସାତ ଦିନ ମଧ୍ୟରେ, କଣ ଆପଣଙ୍କୁ ନିଜ ଘରର ଆବଶ୍ୟକତା ପାଇଁ ଦରକାର ହେଉଥିବା ପାଣି ମିଳିବାରେ କିଛି ଅସୁବିଧା ହେଇଛି କି ? | ☐ 1. Yes ହଁ<br>☐ 2. No ନାହିଁ<br><br>**Please explain. ଦୟାକରି ବର୍ଣ୍ଣନା କରନ୍ତୁ** |
| **C21.** Has coronavirus or the lockdown changed your **drinking water practices** such as where you get your water, how you store it, and whether or not you clean/treat the water? Please explain.<br><br>କଣ କରୋନାଭାଇରସ କିମ୍ବା ଲକଡ଼ାଉନ ଆପଣଙ୍କ **ପାଣି ପିଇବା ସହିତ ଜଡିତ ପ୍ରକ୍ରିୟା ମାନଙ୍କୁ** ବଦଳେଇଛି କି, ଯେପରିକି ଆପଣ ପାଣି କେଉଁଠାରୁ ଆଣନ୍ତି, ତାକୁ କିପରି ରଖନ୍ତି କିମ୍ବା କିପରି ପରିଷ୍କାର କରନ୍ତି? ଦୟାକରି ବର୍ଣ୍ଣନା କରନ୍ତୁ। | Change in **where you get** your drinking water?<br>**ଆପଣ ପିଇବା ପାଣି ଆଣୁଥିବା ସ୍ଥାନରେ ପରିବର୍ତ୍ତନ**?<br>☐ 1. Yes ହଁ<br>☐ 2. No ନାହିଁ<br><br>Change in **how you store** your drinking water?<br>ଆପଣ କିପରି ପିଇବା ପାଣିକୁ ରଖନ୍ତି, ସେଥିରେ ପରିବର୍ତ୍ତନ?<br>☐ 1. Yes ହଁ<br>☐ 2. No ନାହିଁ<br><br>Change in whether or not you **clean/treat** your drinking water?<br>ଆପଣ ନିଜ ପିଇବା ପାଣିକୁ କିପରି ସଫା/ପରିଷ୍କାର କରନ୍ତି, ସେଥିରେ ପରିବର୍ତ୍ତନ?<br>☐ 1. Yes ହଁ<br>☐ 2. No ନାହିଁ<br><br>**Please explain. ଦୟାକରି ବର୍ଣ୍ଣନା କରନ୍ତୁ** |

<table>
<tr><td>C22. Has coronavirus or the lockdown changed your household cleaning practices? Please explain.<br><br>କଣ କରୋନାଭାଇରସ କିମ୍ବା ଲକଡ଼ାଉନ ଆପଣଙ୍କ ଘର ସଫା କରିବା ପ୍ରକ୍ରିୟାକୁ ବଦଳେଇଛି? ଦୟାକରି ବର୍ଣ୍ଣନା କରନ୍ତୁ<br><br>Probes: Has there been any change in how often you clean your home?<br><br>କଣ ଆପଣ ପ୍ରାୟତଃ କେତେଥର ଘର ସଫା କରନ୍ତି, ସେଥିରେ କିଛି ପରିବର୍ତ୍ତନ ଆସିଛି କି ?<br><br>Has there been any change in how much detergent/disinfectant or water you use to clean your home?<br><br>କଣ ଆପଣ ଘର ସଫା କରିବା ପାଇଁ ପ୍ରାୟତଃ ଯେତେ ଲୁଗାଧୁଆ ପାଉଡର/ ଫିନାଇଲ କିମ୍ବା ପାଣି ବ୍ୟବହାର କରନ୍ତି, ସେଥିରେ କିଛି ପରିବର୍ତ୍ତନ ହେଇଛି କି?</td><td>☐ 1. Yes ହଁ<br>☐ 2. No ନାହିଁ<br><br><u>Please explain. ଦୟାକରି ବର୍ଣ୍ଣନା କରନ୍ତୁ</u></td></tr>
<tr><td colspan="2">END OF INTERVIEW: Thank the respondent for their time and give them the COVID-19 information and resources summary provided by Gram Vikas.<br><br>ସାକ୍ଷାତକରର ଶେଷ: ଉତ୍ତରଦାତାଙ୍କୁ ତାଙ୍କ ସମୟ ପାଇଁ ଧନ୍ୟବାଦ ଜଣାନ୍ତୁ ଏବଂ ତାଙ୍କୁ ଗ୍ରାମବିକାଶ ଦ୍ୱାରା ଦିଆଯାଇଥିବା COVID-19 ସୂଚନା ବିଷୟରେ ଅବଗତ କରନ୍ତୁ</td></tr>
<tr><td>End Time<br>ଶେଷ ସମୟ</td><td></td></tr>
</table>
